# Estimated transmissibility and impact of SARS-CoV-2 lineage B.1.1.7 in England

**DOI:** 10.1101/2020.12.24.20248822

**Authors:** Nicholas G. Davies, Sam Abbott, Rosanna C. Barnard, Christopher I. Jarvis, Adam J. Kucharski, James D. Munday, Carl A. B. Pearson, Timothy W. Russell, Damien C. Tully, Alex D. Washburne, Tom Wenseleers, Amy Gimma, William Waites, Kerry L. M. Wong, Kevin van Zandvoort, Justin D. Silverman, CMMID COVID-19 Working Group, The COVID-19 Genomics UK (COG-UK) Consortium, Karla Diaz-Ordaz, Ruth Keogh, Rosalind M. Eggo, Sebastian Funk, Mark Jit, Katherine E. Atkins, W. John Edmunds

## Abstract

A novel SARS-CoV-2 variant, VOC 202012/01 (lineage B.1.1.7), emerged in southeast England in November 2020 and is rapidly spreading towards fixation. Using a variety of statistical and dynamic modelling approaches, we estimate that this variant has a 43–90% (range of 95% credible intervals 38–130%) higher reproduction number than preexisting variants. A fitted two-strain dynamic transmission model shows that VOC 202012/01 will lead to large resurgences of COVID-19 cases. Without stringent control measures, including limited closure of educational institutions and a greatly accelerated vaccine roll-out, COVID-19 hospitalisations and deaths across England in 2021 will exceed those in 2020. Concerningly, VOC 202012/01 has spread globally and exhibits a similar transmission increase (59–74%) in Denmark, Switzerland, and the United States.

---

In December 2020, evidence began to emerge that a novel SARS-CoV-2 variant, Variant of Concern 202012/01 (lineage B.1.1.7, henceforth VOC 202012/01), was rapidly outcompeting preexisting variants in southeast England (*1*). The variant increased in incidence during a national lockdown in November 2020, which was mandated in response to a previous and unrelated surge in COVID-19 cases, and continued to spread following the lockdown despite ongoing restrictions in many of the most affected areas. Concern over this variant led the UK government to enact stronger restrictions in these regions on 20 December 2020, and eventually to impose a third national lockdown on 5 January 2021. As of 15 February 2021, VOC 202012/01 comprises roughly 95% of new SARS-CoV-2 infections in England, and has now been identified in at least 82 countries (*2*). Our current understanding of effective pharmaceutical and non-pharmaceutical control of SARS-CoV-2 does not reflect the epidemiological and clinical characteristics of VOC 202012/01. Estimates of the growth rate, disease severity, and impact of this novel variant are crucial for informing rapid policy responses to this potential threat.

## Characteristics of the new variant

VOC 202012/01 is defined by 17 mutations (14 non-synonymous point mutations and 3 deletions), of which eight are in the spike protein, which mediates SARS-CoV-2 attachment and entry into human cells. At least three mutations potentially affect viral function. Mutation N501Y is a key contact residue in the receptor binding domain and enhances virus binding affinity to human angiotensin converting enzyme 2 (ACE2) (*3, 4*). Mutation P681H is immediately adjacent to the furin cleavage site in spike, a known region of importance for infection and transmission (*5, 6*). Deletion ΔH69/ΔV70 in spike has arisen in multiple independent lineages of SARS-CoV-2, is linked to immune escape in immunocompromised patients, and enhances viral infectivity in vitro (*7, 8*). This deletion is also responsible for certain commercial testing kits failing to detect the spike glycoprotein gene, with genomic data confirming these S gene target failures in England are now overwhelmingly due to the new variant (*1*).

The proportion of COVID-19 cases attributable to VOC 202012/01 is rapidly increasing in all regions of England, following an initial expansion in the South East (**Fig. 1A**), and is spreading at comparable rates among males and females and across age and socioeconomic strata (**Fig. 1B**). One potential explanation for the spread of VOC 202012/01 within England is a founder effect: that is, if certain regions had higher levels of transmission as a result of more social interactions, variants that were more prevalent within these regions could become more common overall. Changes in social contact patterns correlate closely with changes in transmission (*9*) (**Fig 1C, D**) and with COVID-19 burden in England (*10*). However, we did not find substantial differences in social interactions between regions of high and low VOC 202012/01 prevalence, as measured by Google mobility (*11*) and social contact survey data (*12*) from September to December 2020 (**Fig. 1E, F**). Therefore, the apparent decoupling between contact rates and transmission in late 2020 may suggest altered transmission characteristics for VOC 202012/01.

**Fig 1.**
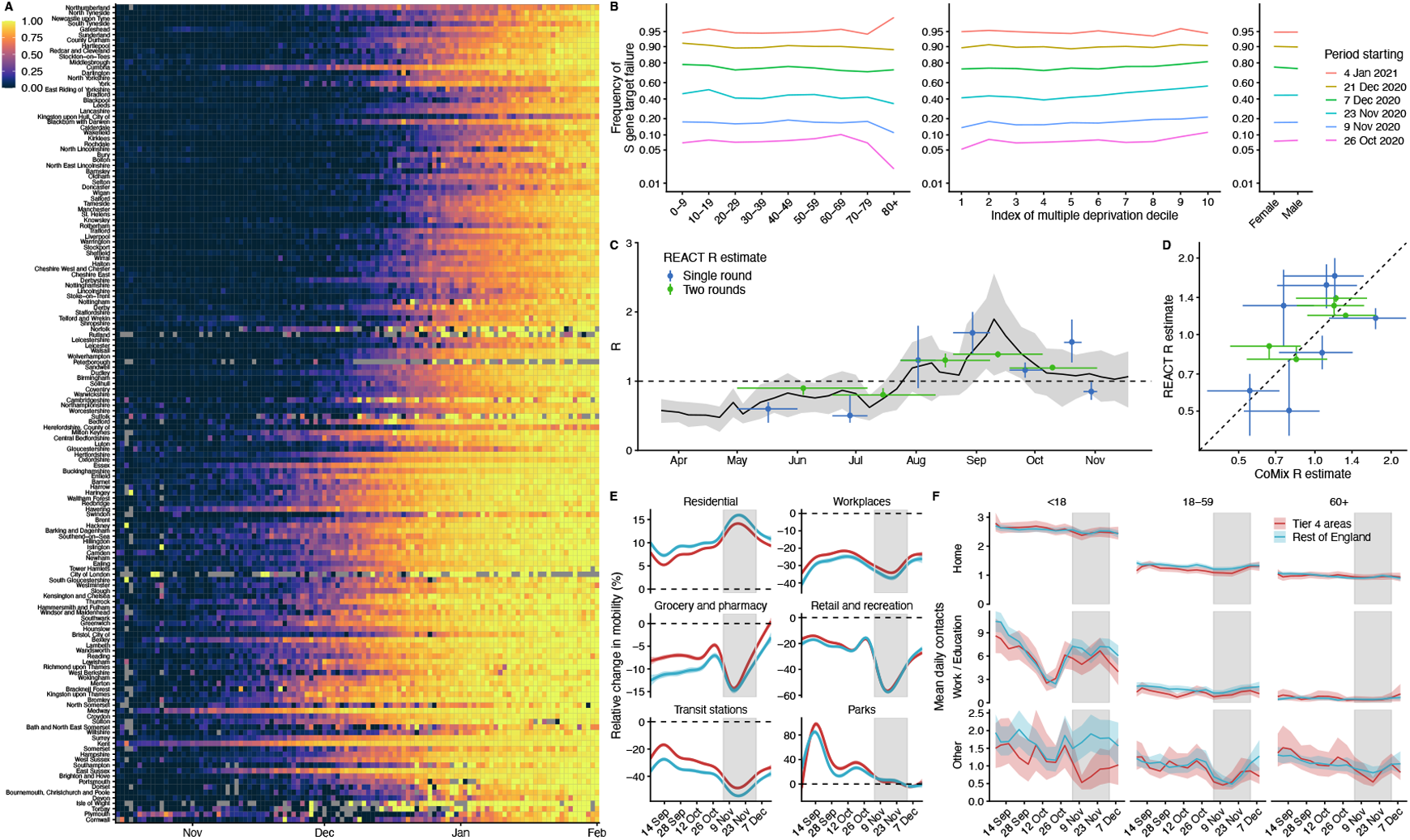
Rapid spread of VOC 202012/01 in England. **(A)** Proportion of S gene target failure among positive Pillar 2 community SARS-CoV-2 tests in upper-tier local authorities of England from 1 October 2020–10 January 2021, sorted by latitude. **(B)** Spread of S gene target failure by age, index of multiple deprivation decile (1 = most deprived), and sex within Greater London. **(C, D)** Estimates of *R*_0_ from CoMix social contact survey (*12*) compared to *R*_t_ estimates from REACT-1 prevalence survey (*9*) for England, with 90% CIs. *R*_*t*_ estimates based on single and aggregated REACT-1 survey rounds are shown. **(E)** Percentage change (95% CI) in Google Mobility indices relative to baseline over time and **(F)** setting-specific mean contacts (95% CI) from the CoMix study (*12*) over time and by age for Tier 4 local authorities compared to the rest of England. Tier 4 local authorities are areas within the South East, East of England, and London regions that were placed under stringent restrictions from 20 December 2020 due to high prevalence of VOC 202012/01 and growing case rates. Grey shaded areas show the second national lockdown in England.

### Measuring the new variant’s growth rate

VOC 202012/01 appears unmatched in its ability to outcompete other SARS-CoV-2 lineages in England. Analysing the COG-UK dataset (*13*), which comprises over 150,000 sequenced SARS-CoV-2 samples from across the UK, we found that the relative population growth rate of VOC 202012/01 in the first 31 days following its initial phylogenetic observation was higher than that of all 307 other lineages with enough observations to obtain reliable growth-rate estimates (**Fig. 2A, S1**). While the relative growth rate of VOC 202012/01 has declined slightly over time, it remains among the highest of any lineage as a function of lineage age (**Fig. 2B**), and the lineage continues to expand.

**Fig 2.**
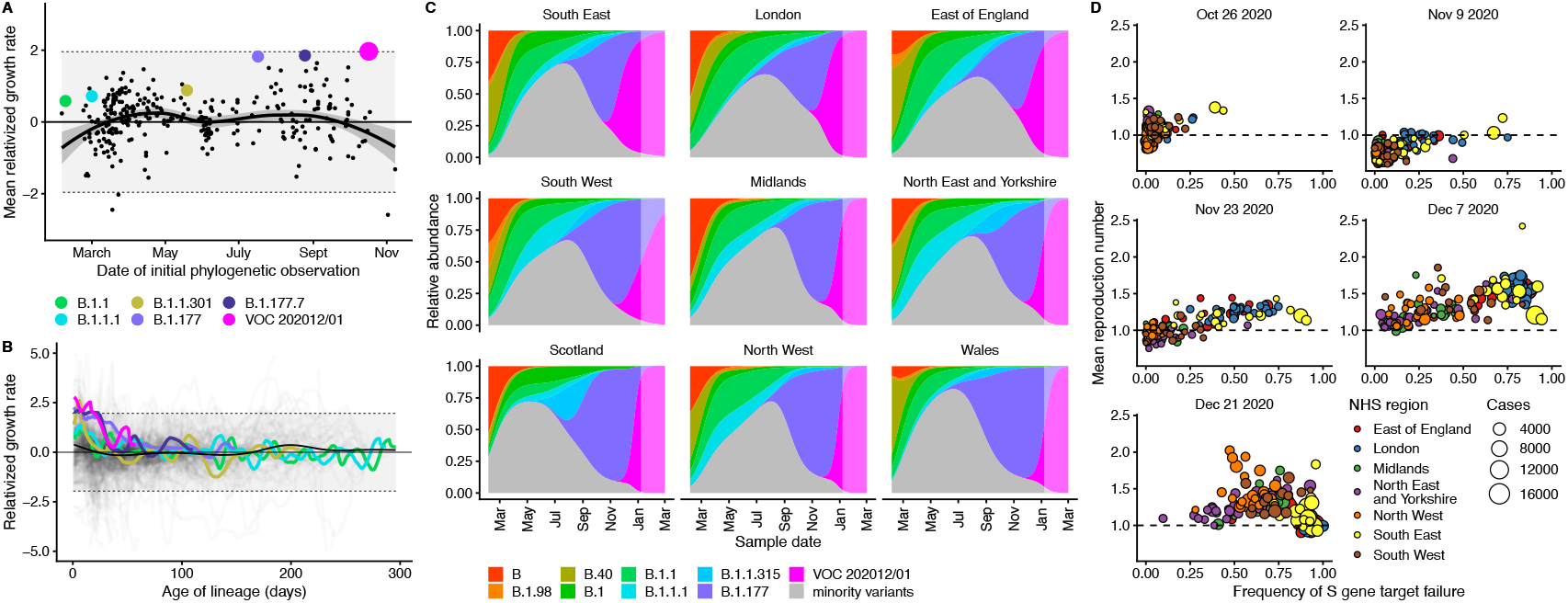
Measuring the growth rate of VOC 202012/01. **(A)** Average relativized growth rate, i.e. a measure of variant fitness relative to other variants present during the 31 days following initial phylogenetic observation of a given variant, for all lineages in the COG-UK dataset, highlighting many lineages that have risen to prominence including B.1.177, the lineage with the highest relative abundance during the IPO of VOC 202012/01. The shaded regions show conservative 95% rejection intervals; VOC 202012/01 is the first strain to exceed this threshold of faster relativized growth. While many lineages exhibit above-average rates of growth, VOC 202012/01 has had the highest average relativized growth of any lineage in the history of COG-UK surveillance of SARS-COV-2. **(B)** Plotting all lineages’ relativized growth rates (*ρ* (*t*)) as a function of lineage age with conservative 95% rejection intervals highlights the significantly faster growth of VOC 202012/01 relative to other lineages at comparable times since their initial observation. Later declines in VOC and B.1.177 correspond to highly uncertain estimates of growth rates for data that are yet to be backfilled, and so these declines in *ρ* (*t*) are sensitive to the processing of future sequences from recent dates (**Fig. S1**). **(C)** Muller plots of the relative abundances of the major SARS-CoV-2 variants in the UK, based on a multinomial spline fit to COG-UK sequence data (separate-slopes multinomial spline model, **Tables 1 & S1**). A model extrapolation until March 1 is shown (shaded area). Minority variants are 440 circulating SARS-CoV-2 variants of low abundance. Specific colours represent the same lineages in panels A–C. **(D)** Mean reproduction number over 7-day periods in 149 upper-tier local authorities of England (coloured by the NHS England region they are within) plotted against the weekly proportion of Pillar 2 community SARS-CoV-2 tests with S gene target failure shows the spread of VOC 202012/01, a corresponding increase in the reproduction number by local authority, and the eventual impact of targeted government restrictions from 20 December 2020. Testing data are shown for the week following the reproduction number estimates to account for delays from infection to test.

To quantify the growth advantage of VOC 202012/01, we performed a series of multinomial and logistic regression analyses on COG-UK data. A time-varying multinomial spline model estimates an increased growth rate for VOC 202012/01 of +0.104 days^-1^ (95% CI 0.100–0.108) relative to the previously dominant lineage, B.1.177 (model 1a, **Table 1**; **Fig. 2C, S2, S3**). Assuming a generation interval of 5.5 days (*14*), this corresponds to a 77% (73–81%) increase in the reproduction number *R*. The growth advantage of VOC 202012/01 persists under more conservative model assumptions (model 1b, **Table 1**; **Fig. S4**), is consistent across all regions of the UK (model 2a, **Table S1**; **Fig. S5**), and is similar when measured from S gene target failures among community COVID-19 tests instead of COG-UK sequence data (model 2h, **Table 1**; **Fig. S6**). Data from other countries yield similar results: we estimate that *R* for VOC 202012/01 relative to other lineages is 55% (45–66%) higher in Denmark, 74% (66–82%) higher in Switzerland, and 59% (56–63%) higher in the United States, with consistent rates of displacement across regions within each country (models 3a–c, **Table 1**; **Figs. S6, S7**).

**Table 1.** Estimates of increased reproduction number for VOC 202012/01. Means and 95% CIs (GLMM) / 95% CrIs (*R*_*t*_ regression, transmission model) shown. GLMM models do not estimate a baseline growth rate or reproduction number. See **Table S1** for full details.

| Model type | | Model assumptions | Data | Geography | Baseline growth rate | Additive increase in growth rate, $\Delta r$ | Baseline reproduction number | Multiplicative increase in reproduction number* |
| --- | --- | --- | --- | --- | --- | --- | --- | --- |
| GLMM | 1a | separate-slopes multinomial spline model† | sequence | regions of UK | — | 0.104 [0.100, 0.108] | — | 77% [73, 81] |
| GLMM | 1b | common-slope multinomial model† | sequence | lower-tier local authorities of UK | — | 0.093 [0.091, 0.095] | — | 67% [65, 69] |
| GLMM | 2h | separate-slope binomial spline model†† | S gene target failure** | regions of England | — | 0.109 [0.107, 0.111] | — | 83% [81, 84] |
| $R_t$ regression | 4a | regional time-varying baseline | S gene target failure | upper-tier local authorities of England | 0.007 [0.002, 0.012] | 0.067 [0.060, 0.073] | 1.04 [1.01, 1.07] | 43% [38, 48] |
| $R_t$ regression | 4b | regional static baseline | S gene target failure | upper-tier local authorities of England | 0.007 [0.002, 0.012] | 0.085 [0.079, 0.091] | 1.04 [1.01, 1.07] | 57% [52, 62] |
| Transmission model | 5a | increased transmissibility | S gene target failure** | regions of England | -0.001 [-0.017, 0.012] | 0.118 [0.067, 0.168] | 1.01 [0.94, 1.09] | 82% [43, 130] |
| GLMM | 3a | common-slope binomial model†† | sequence | regions of Denmark | — | 0.080 [0.067, 0.092] | — | 55% [45, 66] |
| GLMM | 3b | common-slope binomial model†† | sequence + RT-PCR rescreening | regions of Switzerland | — | 0.101 [0.092, 0.109] | — | 74% [66, 82] |
| GLMM | 3c | common-slope binomial model†† | S gene target failure** | states of USA | — | 0.084 [0.080, 0.088] | — | 59% [56, 83] |

† VOC 202012/01 versus B.1.177
†† VOC 202012/01 versus all other variants
\* Increases in the reproduction number assume a generation interval of 5.5 days.
\*\* Binomial counts adjusted for the true positive rate (proportion of S gene target failures that are VOC 202012/01), estimated from misclassification model (for UK) or a binomial GLMM fitted to sequencing data of S gene target failures (for US).

As an alternative approach, we performed a regression analysis of previously-estimated reproduction numbers from case data against the frequency of S gene target failure in English upper-tier local authorities (**Fig. 2D**), using local control policies and mobility data as covariates and including a time-varying spline to capture any unmeasured confounders. This yielded an estimated increase in *R* for VOC 202012/01 of 43% (38–48%), increasing to a 57% (52–62%) increase if the spline was not included (model 4a–b, **Table 1**). The various statistical models we fitted yield slightly different estimates for the growth rate of VOC 202012/01, reflecting different assumptions and model structures, but all identify a substantially increased growth rate (**Table S1**).

### Mechanistic hypotheses for the rapid spread

To understand possible biological mechanisms for why VOC 202012/01 spreads more quickly than preexisting variants, we extended an age- and regionally-structured mathematical model of SARS-CoV-2 transmission (*10, 15*) to consider two co-circulating variants (**Fig. S8; Tables S2, S3**). The model uses Google mobility data (*11*), validated by social contact surveys (*10*), to capture changes in contact patterns over time for each region of England. We created five versions of the model, each including one alternative parameter capturing a potential mechanism.

The hypotheses we tested are as follows. First, observations of lower Ct values (*16*–*18*)—i.e., higher viral load—support that VOC may be more transmissible per contact with an infectious person than preexisting variants (hypothesis 1). Second, longitudinal testing data (*17*) suggest that VOC may be associated with a longer period of viral shedding, and hence a potentially longer infectious period (hypothesis 2). Third, the ΔH69/ΔV70 deletion in spike contributed to immune escape in an immunocompromised patient (*7*), potentially suggesting that immunity to preexisting variants affords reduced protection against infection with VOC (hypothesis 3). Fourth, that VOC initially spread during the November 2020 lockdown in England, during which schools were open, suggests that children may be more susceptible to infection with VOC than with preexisting variants (hypothesis 4). Children are typically less susceptible to SARS-CoV-2 infection than adults (*19, 20*), possibly because of immune cross-protection due to other human coronaviruses (*21*), which could be less protective against VOC. Finally, VOC could have a shorter generation time than preexisting variants (hypothesis 5). A shorter generation time could account for an increased growth rate without requiring a higher reproduction number, which would make control of VOC 202012/01 via social distancing measures relatively easier to achieve.

We fit each model to time series of COVID-19 deaths, hospital admissions, hospital and ICU bed occupancy, PCR prevalence, seroprevalence, and the proportion of community SARS-CoV-2 tests with S gene target failure across the three most heavily affected NHS England regions, over the period of 1 March – 24 December 2020 (**Figs. 3, S9–S14**). We assess models using Deviance Information Criteria (DIC) and by comparing model predictions to observed data for the 14 days following the fitting period (i.e., 25 December 2020 – 7 January 2021). Of the five hypotheses assessed, hypothesis 1 (increased transmissibility) had the lowest (i.e., best) combined DIC and predictive deviance. Hypotheses 2 (longer infectious period) and 4 (increased susceptibility in children) also fitted the data well, although hypothesis 4 is not well supported by household secondary attack rate data (**Fig. S15**) or by age-specific patterns of S gene target failure in the community (**Fig. S16**), neither of which identify a substantial increase in susceptibility among children. Hypotheses 3 (immune escape) and 5 (shorter generation time) fit poorly (**Fig. 3A; Table S4**). In particular, hypothesis 5 predicted that the relative frequency of VOC 202012/01 should have dropped during stringent restrictions in late December 2020, because when two variants have the same *R_t_* < 1 but different generation times, infections decline faster for the variant with the shorter generation time.

**Fig 3.**
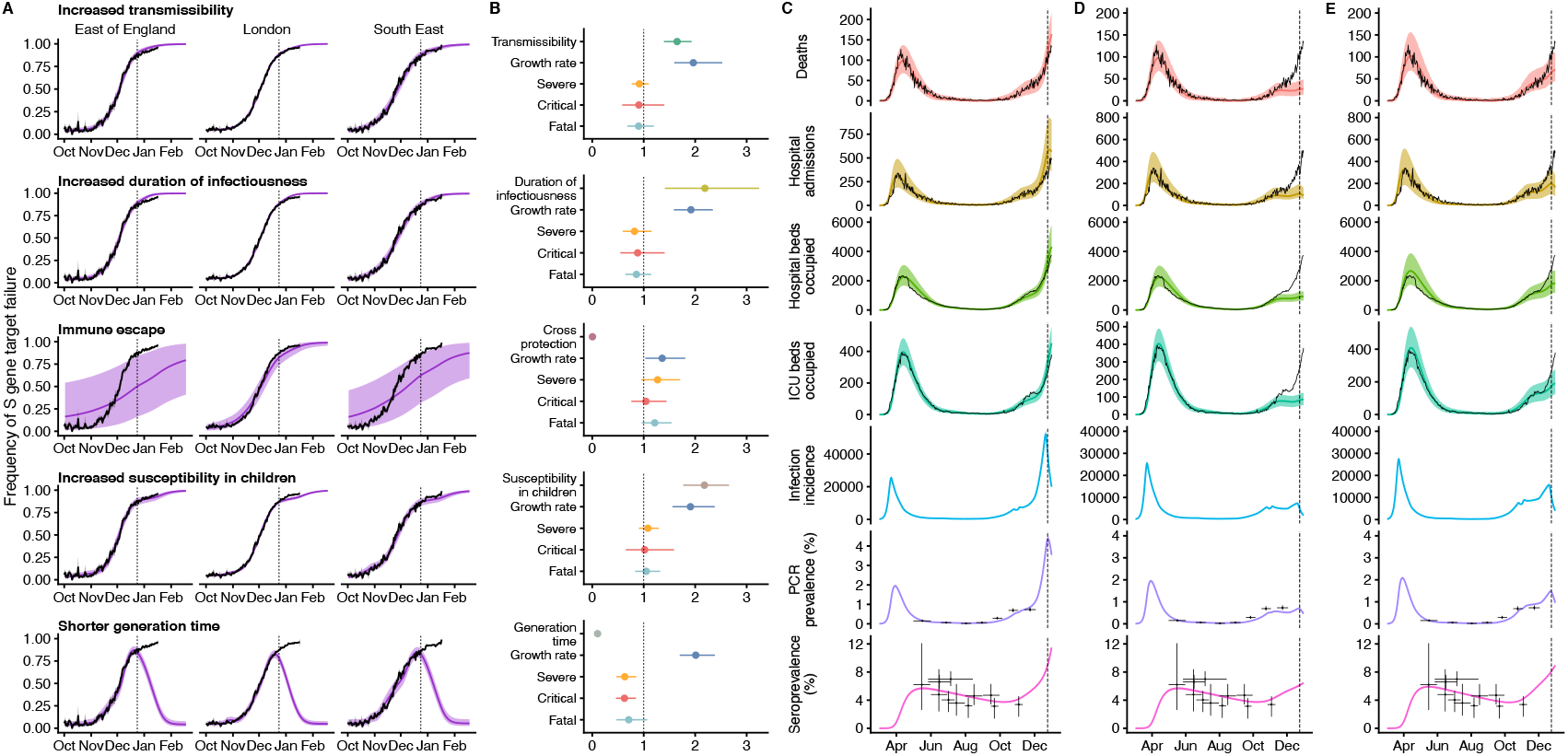
Comparison of possible biological mechanisms underlying the rapid spread of VOC 202012/01. Each row shows a different assumed mechanism. **(A)** Relative frequency of VOC 202012/01 (black line and ribbon shows observed S gene target failure frequency with 95% binomial credible interval; purple line and ribbon shows mean and 95% credible interval from model fit). **(B)** Posterior estimates for relative odds of hospitalisation (severe illness), relative odds of ICU admission (critical illness), relative odds of death (fatal illness), growth rate as a multiplicative factor per week (i.e. exp(7·Δ*r*)), and the parameter that defines the hypothesised mechanism; all parameters are relative to those estimated for preexisting variants. Illustrative model fits for the South East NHS England region: **(C)** fitted two-strain increased transmissibility model with VOC 202012/01 included; **(D)** fitted two-strain increased transmissibility model with VOC 202012/01 removed; **(E)** fitted single-strain model without emergence of VOC 202012/01.

We fitted a combined model incorporating the five hypotheses above, but it was not able to identify a single consistent mechanism across NHS England regions, demonstrating that a wide range of parameter values are compatible with the observed growth rate of VOC 202012/01 (**Fig. S14**). Based on our analysis, we identify increased transmissibility as the most parsimonious model, but emphasize that the five mechanisms explored here are not mutually exclusive and may be operating in concert.

The increased transmissibility model does not identify a clear increase or decrease in the severity of disease associated with VOC 202012/01, finding similar odds of hospitalisation (estimated odds ratio of hospitalisation given infection, 0.92 [95% credible intervals 0.77–1.10]), critical illness (OR 0.90 [0.58–1.40]), and death (OR 0.90 [0.68–1.20]), based upon fitting to the three most heavily affected NHS England regions (**Fig. 3B**). These estimates should be treated with caution, as we would not expect to identify a clear signal of severity when fitting to data up to 24 December 2020, given delays between infection and hospitalization or death. However, the fitted model finds strong evidence of higher relative transmissibility, estimated at 65% (95% CrI: 39– 93%) higher than preexisting variants for the three most heavily affected NHS England regions, or 82% (43–130%) when estimated across all seven NHS England regions (model 5a, **Table 1**). These estimates of increased transmissibility are consistent with our statistical estimates and with a previous estimate of a 70% increased reproduction number for VOC 202012/01 (*16*). This model reproduces observed epidemiological dynamics for VOC 202012/01 (**Figs. 3C, S17**). Without the introduction of a new variant with a higher growth rate, the model is unable to reproduce observed dynamics (**Fig. 3D–E, Fig. S17–S19**), further highlighting that changing contact patterns do not explain the spread of VOC 202012/01.

### Implications for COVID-19 dynamics in England

Using the best-performing transmission model (increased transmissibility) fitted to all seven NHS England regions, we compared projected epidemic dynamics under different assumptions about control measures from mid-December 2020 to the end of June 2021. We compared four scenarios for non-pharmaceutical interventions (NPIs) introduced on 1 January 2021: (i) a moderate-stringency scenario with mobility levels as observed in the first half of October 2020; (ii) a high-stringency scenario with mobility levels as observed during the second national lockdown in England in November 2020, with schools open; (iii) the same high-stringency scenario, but with schools closed until 15 February 2021; and (iv) a very high-stringency scenario with mobility levels as observed during the first national lockdown in early April 2020, with schools closed (**Fig. S20**). In combination with these NPI scenarios, we considered three vaccination scenarios: no vaccinations; 200,000 vaccinations per week; and 2 million vaccinations per week. We assumed that vaccine rollout starts on 1 January 2021 and that vaccinated individuals have a 95% lower probability of disease and a 60% lower probability of infection than unvaccinated individuals. For simplicity, we assumed that vaccine protection was conferred immediately upon receipt of one vaccine dose. Note that these projections serve as indicative scenarios rather than formal predictive forecasts.

Regardless of control measures, all regions of England were projected to experience a new wave of COVID-19 cases and deaths in early 2021, peaking in February 2021 if no substantial control measures are introduced, or in mid-January 2021 if strong control measures succeeded in reducing *R* below 1 (**Fig. 4A**). In the absence of substantial vaccine roll-out, the number of COVID-19 cases, hospitalisations, ICU admissions and deaths in 2021 were predicted to exceed those in 2020, even with stringent NPIs in place (**Table 2**). Implementing more stringent measures in January 2021 (scenarios iii and iv) led to a larger rebound in cases when simulated restrictions were lifted in March 2021, particularly in those regions that had been least affected up to December 2020 (**Fig. S21**). However, these more stringent measures may buy time to reach more widespread population immunity through vaccination. Vaccine roll-out further mitigates transmission, although the impact of vaccinating 200,000 people per week—similar in magnitude to the rates reached in December 2020—was relatively small (**Fig. 4B, Fig. S22**). An accelerated uptake of 2 million people fully vaccinated per week (i.e., 4 million doses for a two-dose vaccine) had a much more substantial impact (**Fig. 4C, Fig. S23**). However, accelerated vaccine roll-out has a relatively limited impact on peak burden, as the peak is largely mediated by the stringency of NPIs enacted in January 2021, before vaccination has much of an impact. The primary benefit of accelerated vaccine roll-out lies in helping to avert a resurgence of cases following the relaxation of NPIs, and in reducing transmission after the peak burden has already been reached.

**Table 2.** Summary of projections for England, 15 December 2020 – 30 June 2021.

**No vaccination**
|  | **Moderate (October 2020)** | **High (November 2020) with schools open** | **High with schools closed** | **Very high (March 2020)** |
| --- | --- | --- | --- | --- |
| **Peak ICU (relative to 1st wave)** | 274% (256 - 292%) | 162% (151 - 172%) | 130% (122 - 136%) | 119% (112 - 124%) |
| **Peak ICU bed requirement** | 9,980 (9,330 - 10,600) | 5,880 (5,490 - 6,280) | 4,720 (4,450 - 4,960) | 4,310 (4,070 - 4,530) |
| **Peak deaths** | 3,960 (3,730 - 4,200) | 2,050 (1,920 - 2,160) | 1,500 (1,440 - 1,570) | 1,830 (1,670 - 2,000) |
| **Total admissions** | 635,000 (604,000 - 659,000) | 454,000 (432,000 - 472,000) | 448,000 (425,000 - 466,000) | 450,000 (425,000 - 472,000) |
| **Total deaths** | 216,000 (205,000 - 227,000) | 146,000 (138,000 - 152,000) | 147,000 (139,000 - 155,000) | 149,000 (140,000 - 157,000) |

**200,000 vaccinations per week**
|  | **Moderate (October 2020)** | **High (November 2020) with schools open** | **High with schools closed** | **Very high (March 2020)** |
| --- | --- | --- | --- | --- |
| **Peak ICU (relative to 1st wave)** | 269% (252 - 287%) | 160% (149 - 170%) | 130% (122 - 136%) | 118% (112 - 124%) |
| **Peak ICU bed requirement** | 9,790 (9,150 - 10,400) | 5,810 (5,430 - 6,200) | 4,710 (4,450 - 4,950) | 4,310 (4,070 - 4,520) |
| **Peak deaths** | 3,700 (3,500 - 3,920) | 1,930 (1,820 - 2,040) | 1,490 (1,430 - 1,550) | 1,320 (1,280 - 1,380) |
| **Total admissions** | 610,000 (580,000 - 634,000) | 438,000 (416,000 - 454,000) | 415,000 (394,000 - 430,000) | 394,000 (373,000 - 413,000) |
| **Total deaths** | 202,000 (192,000 - 213,000) | 137,000 (130,000 - 143,000) | 129,000 (123,000 - 135,000) | 119,000 (112,000 - 125,000) |

**2 million vaccinations per week**
|  | **Moderate (October 2020)** | **High (November 2020) with schools open** | **High with schools closed** | **Very high (March 2020)** |
| --- | --- | --- | --- | --- |
| **Peak ICU (relative to 1st wave)** | 236% (221 - 252%) | 149% (139 - 158%) | 128% (121 - 134%) | 118% (111 - 124%) |
| **Peak ICU bed requirement** | 8,590 (8,050 - 9,170) | 5,400 (5,070 - 5,760) | 4,650 (4,390 - 4,880) | 4,290 (4,060 - 4,500) |
| **Peak deaths** | 2,470 (2,330 - 2,610) | 1,510 (1,450 - 1,580) | 1,390 (1,340 - 1,450) | 1,290 (1,250 - 1,340) |
| **Total admissions** | 483,000 (459,000 - 502,000) | 353,000 (337,000 - 366,000) | 277,000 (265,000 - 287,000) | 190,000 (182,000 - 197,000) |
| **Total deaths** | 140,000 (133,000 - 146,000) | 98,900 (94,600 - 103,000) | 81,000 (77,600 - 84,200) | 58,200 (56,100 - 60,300) |

**Fig 4.**
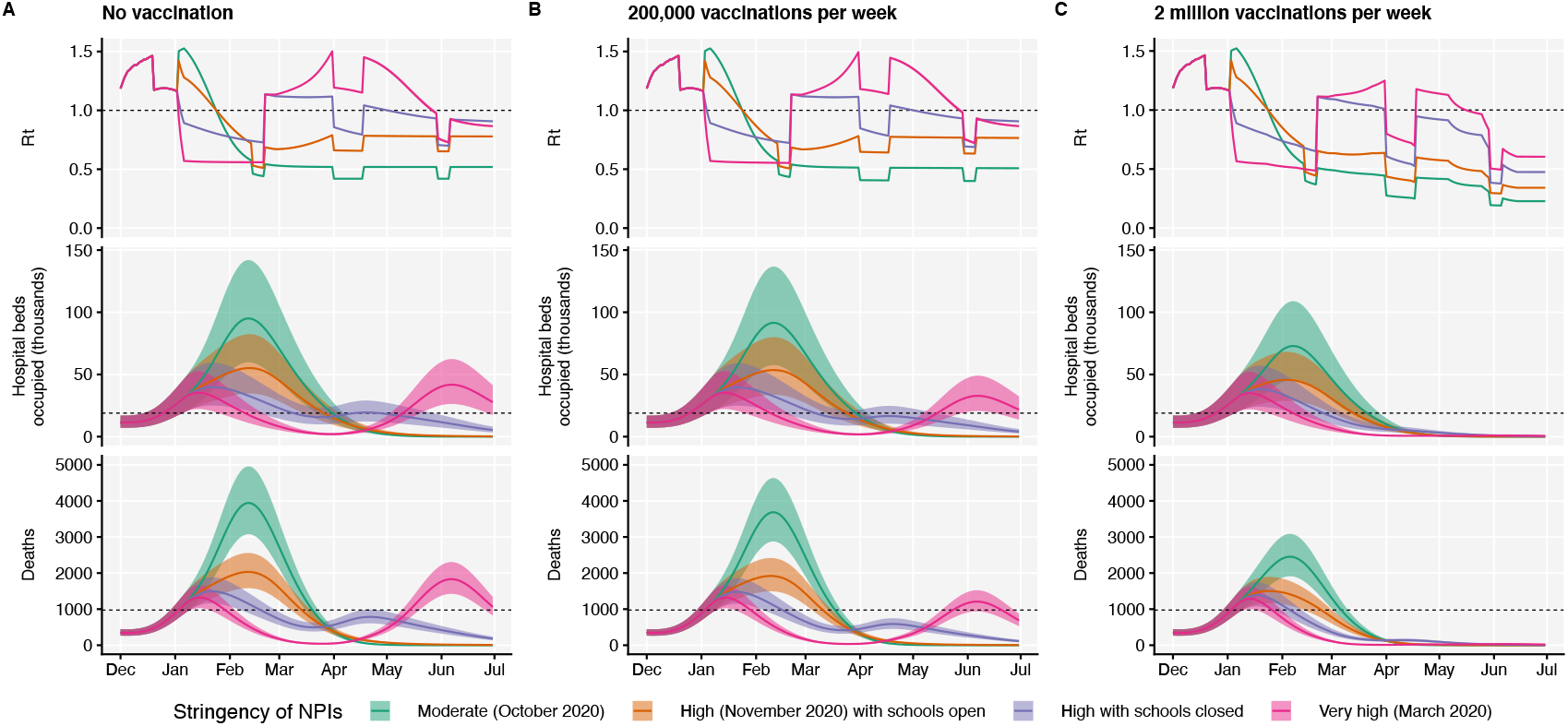
Projections of epidemic dynamics under different control measures. We compare four alternative scenarios for non-pharmaceutical interventions from 1 January 2021: (i) mobility returning to levels observed during relatively moderate restrictions in early October 2020; (ii) mobility as observed during the second lockdown in England in November 2020, then gradually returning to October 2020 levels from 1 March to 1 April 2021, with schools open; (iii) as (ii), but with school closed until 15 February 2021; (iv) as (iii), but with a lockdown of greater stringency as observed in March 2020 (Fig. S20). **(A)** Without vaccination. **(B)** With 200,000 people vaccinated per week. **(C)** With 2 million people vaccinated per week. We assume that vaccination confers 95% vaccine efficacy against disease and 60% vaccine efficacy against infection, and that vaccination starts on 1 January 2021 with vaccine protection starting immediately upon receipt. This is intended to approximate the fact that vaccination started in early December, but that full protection occurs after a time lag and potentially after a second dose. Vaccines are given first to 70+ year olds until 85% coverage is reached in this age group, then to 60+ year olds until 85% coverage is reached in this age group, continuing into younger age groups in 10-year decrements. Resurgences starting in March 2021 are due to the relaxation of non-pharmaceutical interventions, including reopening schools (**Fig. S20**). Median and 95% credible intervals are shown. The dotted lines in rows 2 and 3 show peak hospitalisations and deaths from the first COVID-19 wave in England (April 2020).

As a sensitivity analysis, we also ran model projections with a seasonal component such that transmission is 20% higher in the winter than in the summer (*22*), but this did not qualitatively affect our results (**Fig. S24, Table S5**).

## Discussion

Combining multiple behavioural and epidemiological data sources with statistical and dynamic modelling, we estimated that the novel SARS-CoV-2 variant VOC 202012/01 has a 43–90% (range of 95% credible intervals 38–130%) higher reproduction number than preexisting variants of SARS-CoV-2 in England, assuming no changes to the generation interval. Based on early population-level data, we were unable to identify whether the new variant is associated with higher disease severity. Theoretical considerations suggest that mutations conferring increased transmissibility to pathogens may be inextricably linked to reduced severity of disease (*23*). However, this framework assumes that a long history of adaptive evolution has rendered mutations yielding increased transmissibility inaccessible without a decrease in virulence, which does not obviously hold for a recently emerged human pathogen such as SARS-CoV-2.

Regardless, without strengthened controls, there is a clear risk that future epidemic waves may be larger – and hence associated with greater burden – than previous waves. The UK government initiated a third national lockdown on 5 January 2021 in response to the rapid spread of VOC 202012/01, including school closures. Educational settings are among the largest institutions linked to SARS-CoV-2 clusters that remained open during the November 2020 lockdown (*24*), which means the enacted school and university closures may substantially assist in reducing the burden of COVID-19 in early 2021.

The rise in transmission from VOC 202012/01 has crucial implications for vaccination. First, it means prompt and efficient vaccine delivery and distribution is even more important to reduce the impact of the epidemic in the near future. Increased transmission resulting from VOC 202012/01 will raise the herd immunity threshold, meaning the potential burden of SARS-CoV-2 is larger and higher vaccine coverage will be required to achieve herd immunity. It is therefore extremely concerning that VOC 202012/01 has spread to at least 82 countries globally (*2*). Although VOC 202012/01 was first identified in England, a rapidly spreading variant has also been detected in South Africa (*25, 26*), where there has been a marked increase in transmission in late 2020. Another variant exhibiting immune escape has emerged in Brazil (*27, 28*). Thus, vaccination timelines will also be a crucial determinant of future burden in other countries where similar new variants are present. Second, there is a need to assess how VOC 202012/01 and other emerging lineages affect the efficacy of vaccines (*29, 30*). Vaccine developers may need to consider developing formulations with variant sequences, and powering post-licensure studies to detect differences in efficacy between the preexisting and new variants. Licensing authorities may need to clarify abbreviated pathways to marketing for vaccines that involve altering strain formulation without any other changes to their composition.

There are limitations to our analysis. We have considered a small number of intervention and vaccination scenarios, which should not be regarded as the only available options for policymakers. Our transmission model does not explicitly capture care home or hospital transmission of SARS-CoV-2, and is fit to each region of England separately rather than pooling information across regions and explicitly modelling transmission between regions. There are also uncertainties in the choice of model used to generate these predictions, and the exact choice will yield differences in the measures needed to control the epidemic. We note that even without increased susceptibility of children to VOC 202012/01, the more efficient spread of the variant implies that the difficult societal decision of closing schools will be a key public health question for multiple countries in the months ahead.

We only assess relative support in the data for the mechanistic hypotheses proposed, but there may be other plausible mechanisms driving the resurgence of cases that we did not consider, and we have not identified the specific combination of mechanisms driving the increased transmission of VOC 202012/01. We identify increased transmissibility as the most parsimonious mechanistic explanation for the higher growth rate of VOC 202012/01, but a longer infectious period also fits the data well (**Table S4**) and is supported by longitudinal testing data (*17*). Our conclusions about school closures were based on the assumption that children had reduced susceptibility and infectiousness compared to adults (*19*), but the precise values of these parameters and the impact of school closures remains the subject of scientific debate (*31*). We based our assumptions about the efficacy of NPIs on the measured impact on mobility of previous national lockdowns in England, but the impact of policy options cannot be predicted with certainty.

Despite these limitations, we found strong evidence that VOC 202012/01 is spreading substantially faster than preexisting SARS-CoV-2 variants. Our modelling analysis suggests this difference could be explained by an overall higher infectiousness of VOC 202012/01, but not by a shorter generation time or immune escape alone. Further experimental work will provide insight into the biological mechanisms for our observations, but given our projections of a rapid rise in incidence from VOC 202012/01—and the detection of other novel and highly-transmissible variants (*25*–*28*)—there is an urgent need to consider what new approaches may be required to sufficiently reduce the ongoing transmission of SARS-CoV-2.

## Materials and Methods

### Summary of control measures in England in late 2020

Following a resurgence of cases in September and October 2020, a second national lockdown was implemented in England, from 5 November to 2 December 2020. Restrictions included a stay-at-home order with exemptions for exercise, essential shopping, obtaining or providing medical care, education and work for those unable to work from home. Schools were kept open. Non-essential shops, retail and leisure venues were required to close. Pubs, bars and restaurants were allowed to offer takeaway services only. Following the second national lockdown, regions in England were assigned to tiered local restrictions according to medium, high and very high alert levels (Tiers 1, 2 and 3). In response to rising cases in southeast England and concerns over VOC 202012/01, the UK government announced on 19 December 2020 that a number of regions in southeast England would be placed into a new, more stringent ‘Tier 4’, corresponding to a Stay at Home alert level. Tier 4 restrictions were broadly similar to the second national lockdown restrictions. As cases continued to rise and VOC 202012/01 spread throughout England, on 5 January 2021 a third national lockdown was introduced in England, with schools and universities closed and individuals advised to stay at home, with measures to be kept in place until at least mid-February 2021.

### Data sources

To assess the spread of VOC 202012/01 in the United Kingdom, we used publicly-available sequencing-based data from the COG-UK Consortium (*13*) (5 February 2020 – 6 January 2021) and Pillar 2 SARS-CoV-2 testing data provided by Public Health England (1 October 2020 – 7 January 2021) for estimating the frequency of S gene target failure in England. COG-UK sequencing data for Northern Ireland were excluded due to low sample sizes.

To assess the spread of VOC 202012/01 in Denmark, Switzerland and the USA, we used publicly available sequence data giving the incidence of VOC 202012/01 aggregated by week and region provided by the Danish Covid-19 Genome Consortium & the Statens Serum Institut (*32*) (15 October 2020 – 28 January 2021), sequence and RT-PCR 501Y.V1 rescreening data giving the incidence of VOC 202012/01 in different regions of Switzerland provided by Christian Althaus and Tanja Stadler and the Geneva University Hospitals, the Swiss Viollier Sequencing Consortium from ETH Zürich, the Risch laboratory, the University Hospital Basel, the Institute for Infectious Diseases, University of Bern and the Swiss National Covid-19 Science Task Force (*33, 34*) (2 November 2020 – 11 Feb 2021), and publicly available US nation-wide Helix SARS-CoV-2 Surveillance data, comprising both S-gene target failure data and randomly selected S-negative samples that were sequenced to infer the proportion of S-negative samples that were the VOC (*35, 36*) (6 September – 11 February 2020).

To estimate mobility, we used anonymised mobility data collected from smartphone users by Google Community Mobility (*11*). Percentage change in mobility per day was calculated for each lower-tier local authority in England and a generalised additive model with a spline for time was fitted to these observations to provide a smoothed effect of the change in mobility over time (Fig. 1C).

To estimate social contact rates (Fig. 1D), we used data on reported social contacts from the CoMix survey (*12*), which is a weekly survey of face-to-face contact patterns, taken from a sample of approximately 2500 individuals broadly representative of the UK population with respect to age and geographical location. We calculated the distribution of contacts using 1000 bootstrap samples with replacement from the raw data. Bootstrap samples were calculated at the participant level, then all observations for those participants are included in a sample to respect the correlation structure of the data. We collect data in two panels which alternate weekly, therefore we calculated the mean smoothed over the 2 week intervals to give a larger number of participants per estimate and account for panel effects. We calculated the mean number of contacts (face to face conversational contact or physical contact) in the settings “home”, “work”, “education” (including childcare, nurseries, schools and universities and colleges), and “other” settings. We calculate the mean contacts by age group and area of residence (those areas which were subsequently placed under Tier 4 restrictions on 20 December 2020 as they were experiencing high and rapidly increasing incidence, and those areas of England that were not placed under these restrictions). The mean number of contacts is influenced by a few individuals who report very high numbers of contacts (often in a work context). The means shown here are calculated based on truncating the maximum number of contacts recorded at 200 per individual per day. We compare *R*_*t*_estimates derived from CoMix (*12*) to those derived from the REACT-1 prevalence survey (*9*) for England.

### Statistical methods in brief

#### Growth of VOC 202012/01 following initial phylogenetic observation

For each lineage *i* in the COG-UK dataset, we pool the number of sequences observed within that lineage across the UK for every day, *t*, yielding integer-valued sequence counts *N*(*i, t*). We estimate the time-varying exponential growth rates of cases of each strain, *r*(*i, t*), using a negative binomial state-space model correcting for day-of-week effects whose dispersion parameter was optimized for each strain by marginal likelihood maximization. We defined the relativized growth rate of a lineage *i* at time *t* as 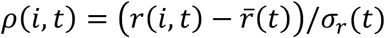, where 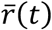 is the average growth rate of all circulating strains at time *t* and *σ*_*r*_(*t*) the standard deviation of growth rates across all lineages at time *t*, such that *ρ*(*i,t*) is analogous to a z-statistic or Wald-type statistic and allows comparison of growth rate differences across time when the average growth rate and scale of growth rate differences varies.

#### Competitive advantage and increased growth rate of VOC-202012/01

To estimate the increase in growth rate of VOC 202012/01, we fitted a set of multinomial and binomial generalized linear mixed models (GLMMs), in which we estimated the rate by which the VOC displaces other resident SARS-CoV-2 variants across different regions in the UK, based on both the COG-UK sequence data as well as the S gene target failure data. In the analysis of the S gene target failure data, binomial counts were adjusted for the true positive rate. For comparison, we also calculated the growth advantage of the VOC in Denmark, Switzerland and the US based on both sequencing and S gene target failure data. All models took into account sample date and region plus, if desired, their interaction, and all mixed models took into account possible overdispersion and for the UK further included local-tier local authority as a random intercept. From these models, we estimated the difference in Malthusian growth rate between other competing variants Δ*r*, as well as the expected multiplicative increase in basic reproduction number *R*_*t*_and infectiousness, assuming unaltered generation time, which can be shown to be equal to exp(Δ*r*.*T*), where *T* is the mean generation interval. The multiplicative increase being equal to exp(Δ*r*.*T*) is an approximation that holds for a delta-distributed generation interval, but we show in the Supplementary Material that this is a good approximation for the gamma-distributed generation interval that we assume. In our calculations, we used estimated SARS-CoV-2 mean generation times *T* of either 5.5 days (*14*) (Table 1) or 3.6 days (*37, 38*) (Table S1).

#### R_t_ analysis

We calculated the weekly proportion of positive tests that were S-gene negative out of all positive tests that tested for the S-gene by English upper-tier local authority. We used reproduction number estimates obtained using the method described in (*37*) and (*39*) and implemented in the EpiNow2 R package (*40*), downloaded from https://github.com/epiforecasts/covid-rt-estimates/blob/master/subnational/united-kingdom-local/cases/summary/rt.csv. We then built a separate model of the expected reproduction number in UTLA *i* during week *t* starting in the week beginning the 5 October 2020 as a function of local restrictions, mobility indicators, residual temporal variation, and proportion of positive tests with S gene target failure. The residual temporal variation is modelled either as a region-specific thin-plate regression spline (“Regional time-varying”) or a static regional parameter (“Regional static”). The key estimand is the relative change in reproduction number in the presence of S gene target failure that is not explained by any of the other variables.

### Transmission dynamic model

We extended a previously developed modelling framework structured by age (in 5-year age bands, with no births, deaths, or aging due to the short timescales modelled) and by geographical region (*10, 15*) to include two variants of SARS-CoV-2 (VOC 202012/01 and non-VOC 202012/01). The model is a discrete-time deterministic compartmental model which allows for arbitrary delay distributions for transitions between compartments. We fitted this model to multiple regionally-stratified data sources across the 7 NHS England regions as previously: deaths, hospital admissions, hospital bed occupancy, ICU bed occupancy, daily incidence of new infections, PCR prevalence of active infection, seroprevalence, and the daily frequency of VOC 202012/01 across each of the regions as measured by S gene target failure frequency corrected for false positives. The model assumes that individuals with clinical symptoms are more infectious than individuals with subclinical infection (*19*). We assume that vaccinated individuals have a lower probability of both clinical and subclinical infection (**Fig. S9**), but that vaccinated individuals who do develop clinical or subclinical infection are as infectious as unvaccinated individuals with clinical or subclinical infection. To model school closure, we removed all school contacts from our contact matrix based upon POLYMOD data and varying over time according to Google Mobility indices, as described previously (*10*). See Supporting Information for details of Bayesian inference including likelihood functions and prior distributions.

Our individual transmission model fits to separate NHS regions of England produce independent estimates of parameters such as relative transmissibility and differences in odds of hospitalisation or death resulting from infection with VOC 202012/01. In order to produce overall estimates for these parameters, we model posterior distributions from individual NHS regions as draws from a mixture distribution, comprising a normally-distributed top-level distribution from which central estimates for each NHS region are drawn. We report the mean and credible intervals of the top-level distribution when reporting model posterior estimates for England.

In model fitting, we assume that our deterministic transmission model approximates the expectation over stochastic epidemic dynamics. This is not exact (*41*) but the error in this approximation is small for the population-level processes we are modelling, as it decays with 1/*N* (*42*). This approach is well developed for state space models of communicable disease dynamics that fit an epidemic process to observed data via a stochastic observation process.

### Apparent growth of VOC 202012/01 not a result of testing artefacts

The apparent frequency of VOC 202012/01 could be inflated relative to reality if this variant leads to increased test-seeking behaviour (e.g. if it leads to a higher rate of symptoms than preexisting variants). However, this would not explain the growth in the relative frequency of VOC 202012/01 over time. Mathematically, if variant 1 has growth rate *r*_1_ and variant 2 has growth rate *r*_2_, the relative frequency over time is *a*_2_exp(*r*_2_*t*) / (*a*_1_exp(*r*_1_*t*) + *a*_2_ exp(*r*_2_*t*)), where *a*_1_ and *a*_2_ are the frequency of variant 1 and 2, respectively, at time *t* = 0. However, if variant 1 has probability *x* of being reported and variant 2 has probability *y*, and both have growth rate *r*, the relative frequency over time is *a*_2_*y* exp(*rt*) / (*a*_1_*x* exp(*rt*) + *a*_2_*y* exp(*rt*)), which is constant.

## Data Availability

All analysis code and data have been made publicly available.

https://www.github.com/nicholasdavies/newcovid

https://zenodo.org/record/4562961

## Acknowledgements

Three anonymous reviewers gave helpful suggestions. We thank Public Health England, COG-UK Consortium volunteers (UK); the Danish Covid-19 Genome Consortium, the Statens Serum Institut (DK); Christian Althaus, Tanja Stadler, Lorenz Risch, the Geneva University Hospitals, the Swiss Viollier Sequencing Consortium at ETH Zürich, the Risch laboratory, the University Hospital Basel, the Institute for Infectious Diseases at University of Bern, the Swiss National Covid-19 Science Task Force (CH); and Helix OpCo, LLC (US) for providing data. Sharon Peacock, Ewan Harrison, Mads Albertsen, Christian Althaus, Tanja Stadler, Lorenz Risch, and Karthik Gangavarapu facilitated data access. Alex Selby suggested improvements to the analysis code. Troy Day gave useful advice for calculating selective benefit and transmission advantage. Stefan Flasche, Rein Houben, Stéphane Hué, Yalda Jafari, Mihály Koltai, Fabienne Krauer, Yang Liu, Rachel Lowe, Billy Quilty, and Julián Villabona Arenas gave input during conception and manuscript drafting.

## Author contributions

NGD, SA, RCB, CIJ, AJK, JDM, CABP, TWR, DCT, ADW, TW, AG, WW, KLMW, KvZ, JDS, KDO, RK, RME, SF, MJ, KEA, and WJE conceived the study, performed analyses, and wrote the manuscript. NGD led the transmission model analysis, CIJ led the mobility analysis, ADW led the relativized growth rate analysis, TW led the GLMM analysis, and SA and SF led the *R*_*t*_ analysis. The CMMID COVID-19 Working Group provided discussion and comments.

## Data and materials availability

All analysis code and data have been archived with Zenodo (*43*). Code and data for the negative binomial state-space model, multinomial and binomial mixed models and transmission dynamic model are maintained at https://www.github.com/nicholasdavies/newcovid, and code and data for the *R*_*t*_ analysis are maintained at https://github.com/epiforecasts/covid19.sgene.utla.rt.

This work is licensed under a Creative Commons Attribution 4.0 International (CC BY 4.0) license, which permits unrestricted use, distribution, and reproduction in any medium, provided the original work is properly cited. To view a copy of this license, visit https://creativecommons.org/licenses/by/4.0/. This license does not apply to figures/photos/artwork or other content included in the article that is credited to a third party; obtain authorization from the rights holder before using such material.

## Supplementary Materials

Materials and Methods

Supplementary Text 1, 2

Figures S1-S24

Tables S1-S6

References 43-75

## Supplementary Materials for

Estimated Transmissibility and Impact of SARS-CoV-2 Variant of Concern 202012/01 in England

### Materials and Methods

#### Growth rate of VOC 202012/01 following its initial phylogenetic observation

It’s possible a strain could get lucky and display faster growth rates than other strains, appearing more transmissible despite not being so. Several confounds can affect the significance of an inference of faster growth in a strain such as VOC 202012/01. For instance, any correlated patterns in people of that network can affect the probability a strain has an impressive run of faster growth rates than other strains - if a new strain discovers a region of a contact network with a higher fraction of susceptible people than that experienced by other strains elsewhere on the contact network, then the lucky strain in a pool of susceptible people may appear to grow faster due to the human population structure and not the virus’ phenotypic traits. Similarly, any changes in NPIs that increase the average risk of transmission across subsets of the contact network (e.g. variation in the tier level across the UK) or any patterns of behavior that increase the variability of the risk of transmission across people in the network (e.g. when some connected groups of people have a higher-than-average risk of transmission due to occupation, less participation in transmission-reducing behaviors, etc.) might affect the probability that a strain exhibits a large run such as that seen in VOC 202012/01.

Furthermore, since defining a “new strain” requires at least 5 genomes of at least 95% coverage co-localized in space (*44*) it’s possible that newly named strains could be more likely to have faster-than-average growth rates as these growing branches of the viral phylogeny may be bioindicators of a spatially (or contact-network) autocorrelated pool of susceptible people with room for further, faster growth.

In this section, we aim to control for time-varying average growth rates, heterogeneity in population structure, and the potential for lineages to be bioindicators of spatially-autocorrelated susceptible populations with an expectation of faster growth after the initial phylogenetic observation (IPO) of the lineage. When accounting for time-varying average growth rates across lineages in circulation, the time varying scale fitness differences across lineages at every point in time, and the time since the initial phylogenetic observation (IPO), the VOC 202012/01 stands out as having the fastest post-IPO relative growth of any lineage in the COG-UK dataset (Fig. 2A&B, main text).

This analysis centered around what we refer to as the “relativized growth rate”. For each lineage *i* in the COG metadata dataset, we pool the number of sequences observed within that lineage across the UK for every day, *t*, yielding integer-valued sequence counts *N*(*i, t*). We estimate the time-varying exponential growth rates of cases of each strain, *r*(*i, t*), using a negative binomial state-space model whose dispersion parameter was optimized for each strain by marginal likelihood maximization. The negative binomial state-space model was implemented using the R package KFAS (*45*) to estimate abundances and growth rates with a second-order polynomial trend to capture time-varying exponential growth/decay and a 7-day seasonal component to correct for day-of-week effects.

To remove the impact of leading zeros on estimates of growth rates, we started estimating growth rates on the first date for which the following week contained at least three observations of the lineage (including the first observation of that week) – we call the first date of this week the “initial phylogenetic observation” or IPO of the lineage. For lagging zeroes, we removed any zeroes after 7 days of consecutive zeros which continued until the final date used in this analysis. As a result of this filtering of leading and lagging zeroes, there was a variable number of lineages each day, but these lineages served as a minimal set of lineages whose growth rates can serve as a reference frame for assessing the significance of the growth and changes in relative abundance of the VOC (*46*).

The final date used in this analysis was determined by an analysis of backfilling patterns of the COG-UK dataset. The COG-UK dataset contains a “sample date” column for every sequence, and samples are not added on the date they are collected but back-filled once samples are shipped, sequenced, and uploaded. As a consequence, the recent dates in the COG-UK dataset exhibit a decline in the total number of counts and lineage richness, a period during which there will be biases in comparing growth rates across lineages with different relative abundances as rare lineages flat-line with zero observations and the observed counts of abundant lineages continue to decline. These biases during the period of backfilling can be further confounded by any differences in the processing times of sequences across surrogate data providers which sample different, non-representative subsets of the UK population. By downloading the COG-UK dataset at multiple dates, we find that over 90% of sequences are accounted for 1 month prior to the download date. Therefore, to avoid biases and confounds due to backfilling, we limit our analysis of growth rates to all but the last 1 month of data in the COG-UK dataset. This results in estimation of growth rates of the VOC up to December 12th, 2020 (**Fig. S1**).

To control for time-varying average growth rates, we defined a statistic we refer to as *relativized growth rates*, denoted *ρ* (*i, t*) for each lineage *i* and time *t*,

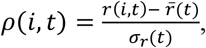

where 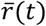 is the average growth rate of all circulating strains at time *t* and σ_*r*_ (*t*)the standard deviation of growth rates across all lineages at time *t*. This statistic is analogous to a z-statistic or Wald-type statistic and allows comparison of growth rate differences across time when the average growth rate and scale of growth rate differences varies. We compute the average relativized fitness of each lineage for the first month after its IPO. This statistic reflects how much faster the lineage grew compared to other lineages circulating for that same month, and allows us to control for potential IPO-effects of lineages whose first observations came at different times in the UK COVID epidemic.

For a lineage to increase in frequency, it mainly needs to increase faster than the lineage with the highest relative abundance, whereas to have an above-average relativized fitness it will need to increase faster than the average lineage (*47*). As such, analyzing relativized growth rates is an additional way to assess not just whether VOC 202012/01 grew faster than the dominant lineage B 1.177—as it’s possible other lineages with similar rarity could have had similar runs of positive growth—but rather test whether or not VOC 202012/01 consistently beat out all other lineages, including the rare ones and recent IPOs, and whether this burst of positive growth post-IPO in the VOC exceeds that of other major lineages’ post-IPO relativized growth.

We plot the relativized fitness as a function of days-since-IPO across all lineages, highlighting a few lineages that have risen to high relative abundance over the course of 2020 (Fig. 2A & B, main text).

#### Competitive advantage and increased growth rate of SARS-CoV2 VOC 202012/01

To infer the competitive advantage of the VOC-202012/01 over other circulating SARS-CoV2 strains (Fig. 2C, main text; Figs. S2–S7) we use the COG-UK sequencing data to calculate the rate by which the strain is displacing other variants and increases in relative abundance *p*. Formally, this is quantified based on the selection (*48*) rate coefficient *s*, which for a newly invading variant is defined as (*49*)

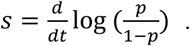

This coefficient measures the rate at which any new variant would displace the resident variant in terms of the increase in the log(odds) to encounter the new variant. A great advantage of the selection rate coefficient is that it can readily be calculated from a logistic regression model as the slope of the proportion of the new variant on a logit (log-odds) link scale in function of time. We can further observe that since the ratio of relative frequencies is equal to the ratio of the absolute representation of the new mutant variant *V* and the wild-type *W* that (*49*)

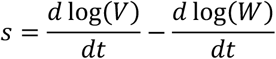

Hence, if selection is density independent and there are no interactions between genotypes, the selection rate is also equal to the difference in Malthusian growth rates between the new variant (*r*_*V*_) and wild-type (*r*_*W*_) (*49*):

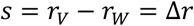

If we further multiply the selection rate by mean generation time *T* then we obtain the dimensionless selection coefficient (*49*)

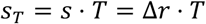

Selection coefficients *s* and *S*_*T*_ represent the most direct measures possible of the fitness advantage enjoyed by any new variant, and are the best possible predictors of whether or not it is expected to increase in frequency during an outbreak (*50*). However, assuming that the generation time of the competing variants remain unaltered (e.g. that the non-infectious period after exposure remains the same), it is also possible to relate the selection coefficient *S*_*T*_ to the expected multiplicative increase in the infectiousness of the virus, as measured by the ratio of the basic reproduction number *R*_*t*_ of the new variant relative to that of the wild type. Specifically, if generation time is gamma distributed with mean *T* and *SD σ*, and if we set *k* = (*σ*/*T*)^2^, it is the case that the basic reproduction number (*51*) *R*_*t*_

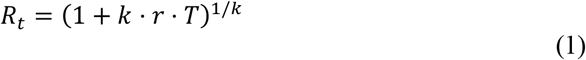

Furthermore, for small *k* (small *SD* of the generation time *σ* relative to the mean *T*), the following approximation (*52*) holds

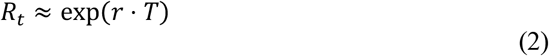

From this, it follows that the ratio of the effective reproduction number of the invading new variant *R*_*V*_ relative to that of the wild type *R*_*W*_, i.e. the expected multiplicative increase in the *R*_*t*_ value *M*, assuming no change in generation time *T* between the variants, equals approximately

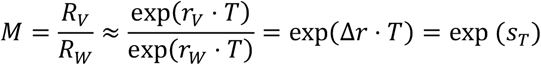

Although this formula is strictly speaking only exact for the limit of *k* → 0 (i.e. with delta-distributed generation times), in practice with our parameter estimates, the error made is extremely small (*52*) even for larger *k*. E.g. with *r*_*v*_ = Δ*r* = 0.11, *r*_*W*_ = 0, *T* = 5.5 days and *σ* = 1.8 (*14*), *k* = 0.33 and application of the exact formula (1) would yield *M* = 1.71, whilst the approximate formula (2) would yield *M* = 1.73, which would amount to an error on *M* of only 1.6%. We should further note that the exp(*Δr* ⋅*T*) formula for the expected multiplicative effect on the *R*_*t*_ values is also approximately correct for exponentially-distributed generation times, where *R*_*t*_ = 1 +*r* ⋅ *T* (*52*) and 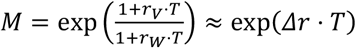 (with small *r*). With gamma-distributed generation times, the exact formula (1) could only be used if we would be able to estimate the variant-specific intrinsic growth rates *r*_*V*_ and *r*_*W*_ (*49*) and corresponding *R*_*t*_ values separately, e.g. using the raw counts, to which one could fit a spline-based Poisson GLM, to yield intrinsic growth rates as the first derivative of the fitted curve on the log link scale. Such a fit, however, would show very large fluctuations due to the implementation of various non-pharmaceutical interventions, and would also require accurate corrections for changes in testing and sequencing intensity over time. Hence, such a calculation would carry a much larger error. Instead, it is much more accurate to estimate the expected multiplicative effect *R*_*t*_ from the rate of change in the log(odds) of the relative abundance of any new variant *p*, Δ*r*. That is, by inferring only the relative growth advantage of the VOC, we can obtain a much more accurate estimate of the competitive advantage than if we would also try to estimate its time-dependent intrinsic absolute growth rate. Furthermore, many superior methods exist to calculate the global, overall *R*_*t*_ value, e.g. ones that also simultaneously take into account hospitalisation data, and given that the *R*_*t*_ value is the average of the *R*_*t*_ value of the individual variants, weighted by their frequency, recalculating expected multiplicative effects on the *R*_*t*_ value to absolute *R*_*t*_ values of the individual variants would still be straightforward.

To estimate pairwise differences in growth rates *Δr* between the VOC variant and other sets of lineages, i.e. pairwise selection rate coefficients, we used both binomial GLMMs (generalized linear mixed models), using data on the representation of pairs of lineages in the COG-UK (*13*) sequencing data (data range: 5 February 2020 to 6 January 2021) or using the Public Health England dataset on the relative frequency of S gene target failure in Pillar 2 SARS-CoV-2 testing data (data range: 1 October 2020 to 17 January 2021) at time of invasion, as well as multinomial spline regression or multinomial mixed models, where we could simultaneously consider the competition for representation among all the major SARS-CoV-2 variants and lineages in different regions across the UK. In both sets of analyses, we considered both the Δ*r* of the VOC 202012/01 (defined as lineage B.1.1.7 and carrying defining mutation N501Y and deletion Δ69/Δ70 in the spike protein) relative to either the earlier dominant lineage B.1.177 (*53*), a set of 410 minority variants, which never reached >13% in the aggregated UK counts in any week or all other circulating variants. For lineage B.1.177, we included any later descendent lineages into the same group.

Binomial GLMMs fit to the COG-UK data included a fixed factor for NHS England region, a continuous covariate for sampling date, the interaction between both if this yielded a more parsimonious fit (based on the Bayesian Information Criterion) or if we were specifically interested to test for differences in rates of spread across regions, as well as random effects for the local-tier local authority (LTLA) and an observation-level random effect to take into account overdispersion (*54*). Binomial GLMMs fit to the UK SGTF data were carried out at the level of NHS England regions, and included NHS region and a natural cubic spline with 3 degrees of freedom in function of sampling date plus the interaction between both as fixed effects and an observation-level random effect to take into account overdispersion (*54*), and binomial counts were adjusted for the true positive rate (i.e. the proportion of S-negative samples that were actually the VOC), which for the UK data were estimated as described in “Misclassification analysis” below. These GLMMs were fit using R’s *glmer* function in the *lme4* package version 1.1.23. For these binomial GLMMs, we used the part of the data where either variant VOC 202012/01 or lineage B.1.177 were initially invading, and for which there was good linearity on a logit scale (Fig. S3). For VOC 202012/01, we therefore used the subset of the data from August 1 2020 onwards, while for lineage B.1.177 we used data for the period between July 1^st^ 2020 and September 30 2020, before it starting to be displaced by VOC 202012/01. From these binomial GLMMs, we subsequently estimated the selection rate Δ*r* from the slope in the log(odds) to encounter the focal variant. Both this marginal slope as well as its 95% confidence intervals were estimated using the *emtrends* function in the *emmeans* R package version 1.5.4-09001. Model predictions or marginal mean model predictions and 95% confidence intervals as well as Tukey posthoc tests to test for differences in slopes (rates of displacement of other strains) across regions were also calculated using this same package. The Tukey posthoc test is used to correct for the familywise error rate. In the calculation of marginal means, we used a bias correction for the presence of the random (*55*) effects. Under the assumption of unaltered generation times, we also made two estimates of the expected multiplicative effect on the *R*_*t*_ value, *M*_*1*_ and *M*_*2*_, based on eqn. (2) above, *M* = *R*_*M*_/*R*_*W*_ ≈ exp(*Δr* ⋅ *T*), using estimated SARS-CoV2 mean generation times *T* of either 3.6 days (*37, 38*) or 5.5 days (*14*). Both the mean and confidence intervals on *Δr* ⋅ *T* were exponentiated, in this way resulting in the estimated geometric mean multiplicative effect on *R*_*t*_. Finally, these effects on reproduction number were expressed as (*M* − 1) × 100, i.e. as the expected percentage increase in *R* for the VOC.

To be able to make a set of other independent baseline estimates of *Δr* outside the UK, we also used binomial GLMMs to estimate the rate of spread by which VOC 202012/01 is displacing other variants in Denmark, Switzerland and the USA, based on openly available data (see *Data sources* in Methods). These analyses included sample date as a continuous covariate, region (or state) as a fixed factor (for Switzerland) or random intercept (for Denmark and the USA), and an observation-level random effect to take into account overdispersion (*54*). Models in which sample date interacted with region (or state) or model incorporating random slopes by region were also fitted, but proved to be less parsimonious based on the BIC criterion. For the US data, binomial counts were adjusted to take into account the true positive rate (the proportion of the S-negative samples that were indeed the VOC), which was estimated using an independent binomial GLMM fitted on sequencing data of S-negative samples, whereby sample date was included as a continuous covariate and state was coded as a random intercept.

Finally, we also fitted two multinomial models in which we considered the multinomial spline model to the COG-UK sequence data using the *multinom* function of the *nnet* R package (*56*) considering the frequencies of 9 major SARS-CoV2 lineages (all reaching at least 13% in some week) as separate variant outcome levels, and subsuming the remaining 410 variants in a category of “minor variants”, thereby allowing us to simultaneously model the competition for representation among all the major variants. This model included a fixed factor region plus a natural cubic spline in function of sample date to allow for slight variation in the selection rate in function of time, plus the interaction between both to allow for different selection rates across regions. A two-degree of freedom natural cubic spline was chosen, as this model both resulted in a visually realistic fit and in a stable and realistic extrapolation (which was no longer the case for natural cubic splines with more knots). In this multinomial model, pairwise *Δr* values between variants VOC 202012/01, B.1.177 and the category of minority variants were calculated using the *emmeans emtrends* function as contrasts in the above-average growth rates of each variant (using argument mode=”latent”(*57*)). Since the growth differences (*Δr*) in this model were time-dependent, we calculated the average growth difference for the VOC vs. minority variants and for the VOC vs. B.1.177 variant contrasts for the period from November 1 2020 onwards and from July 1st 2020 until the 30th of September 2020, respectively, when each of these variants were actively invading in the population. Second, we also fit a multinomial mixed model in which we included a random intercept for the local-tier local authority (LTLA) and also jointly estimated overdispersion. To allow us to estimate the average growth advantage of the VOC, this model was fit under the assumption of identical and non-time varying selection coefficients across regions, and included NHS region and sample date as additive main effects. This model was fit using the *mblogit* function of the *mclogit* R package. The difference in growth rate relative to a particular chosen reference variant was in this model directly inferred from the model coefficients. Finally, the predictions of both models were used to produce Muller plots, to display the change in relative frequencies of the major SARS-CoV2 lineages over time in the UK (Fig. 2C, main text, and Fig. S4).

#### Rt analysis

For the Rt analysis, we used 4 main sources of data: test positive Covid-19 notifications by UTLA (*58*), S-gene status from PCR tests by local authority provided by Public Health England (PHE), Google mobility data stratified by context (*11*), and two publicly available databases of of non-pharmaceutical interventions by UTLA (*59, 60*). We aggregated the data at the weekly level and restricted the analysis to the period beginning Monday, 5 October.

We calculated the weekly proportion of positive tests that were S-gene negative over time by local authority. We estimated reproduction numbers using the method described in (*37*) and (*39*) and implemented in the *EpiNow2* R package (*61*). Daily updated estimates can be downloaded at https://github.com/epiforecasts/covid-rt-estimates/blob/master/subnational/united-kingdom-local/cases/summary/rt.csv. We used two sets of estimates, obtained using uncertain, gamma distributed, generation interval distributions with a mean of 3.6 days (standard deviation (SD): 0.7), and SD of 3.1 days (SD: 0.8) (*38*) or with a mean of 5.5 days (SD: 0.5 days), and SD of 2.1 days (SD: 0.25 days) (*14*), respectively.

We then built a separate model of the expected reproduction number in UTLA *i* during week *t* starting in the week beginning 14 September 2020 as a function of local restrictions, mobility indicators, residual temporal variation, and proportion of positive tests S-gene negative:

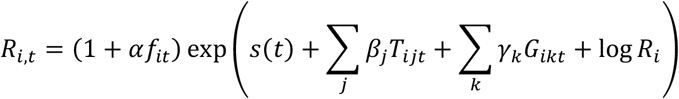

where *R*_*i*_ is an UTLA-level intercept corresponding to Rt during national lockdown in November, *T*_*ijt*_ is 1 if intervention *j* (out of: no tiers, tier 1/2/3) is in place and 0 otherwise, *G*_*ikt;*_ is the relative mobility in context *k* (home, parks, workplace, etc.) at time *t* in UTLA *i* as measured by Google, and *s*(*t*) is a time-varying component, modelled either as a region-specific thin-plate regression spline (“Regional time-varying”) or a static regional parameter (“Regional static”). The key parameter is *α*, the relative change in reproduction number in the presence of the SGTF that is not explained by any of the other variables, where *f*_*it*_ is the proportion out of all positive tests for SARS-CoV-2 where the S-gene was tested with SGTF, and the reproduction number in any given UTLA is

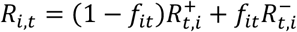

where 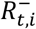 is the S-gene negative reproduction number, 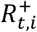 is the S-gene positive reproduction number, and it is assumed that 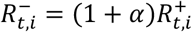.

We used a Student’s t-distribution observation model with a single variance parameter and a single degrees of freedom parameter. All models were implemented using the *brms* (*62*) package in R. All code required to reproduce this analysis is available from https://github.com/epiforecasts/covid19.sgene.utla.rt/.

#### Analysis of differential age susceptibility for VOC 202012/01 based on secondary attack rates

To determine if there was any difference across age cohorts in susceptibility for the new VOC-202012/01, we analysed the age-stratified aggregated data of secondary attack rates reported by Public Health England (*63*) using a binomial GLM (**Fig. S8**). These data comprise secondary attack rates among contact tracing data (from NHS Test and Trace) for the variant of concern (VOC 202012/01), with the identity of strain carried by the index patients (VOC or not) called based on either genomic sequence or S-gene target failure (SGTF) data, and with data split by age bracket of the person that was infected. In total, the dataset contains 17,701 and 456,086 secondary contact records of known age with index patients for which either sequence or SGTF data were available, for the period between 30 November 2020 and 20 December 2020. Out of these secondary contacts, 2,455 and 64,325 became cases, which translates into overall secondary attack rates of 13.87% and 14.10%. To determine the odds ratios for people to be infected by index patients carrying the VOC vs. by those carrying other variants, we fitted a binomial GLM with factors data type (sequence data or SGTF data), age group, variant (VOC or other strains) plus all first order interaction effects. Overdispersion was tested for by fitting an equivalent quasibinomial GLM, but was found to be absent. The R package *emmeans* was used to make effect plots of marginal and predicted means and carry out Sidak posthoc tests to test if the odds for people to be infected by index patients carrying the VOC was higher than that for those carrying other strains (*64*) across the different age categories as well as overall. Possible differential age susceptibility was tested for by comparing the log(odds ratios) for people of different age to be infected by the VOC against the average log(odds ratio) for people to be infected by the VOC overall. These age group × variant interaction contrasts were again calculated using the *emmeans* package, employing a Sidak *p* value correction for multiple testing. Type III Anova tests were carried out using the *Anova* function in R’s *MASS* package.

This analysis identified a small, but non-significant increase in secondary attack rate among children aged 0-9 (binomial GLM, Sidak age group × variant interaction contrast, P = 0.72) and a small, but nonsignificant decrease in secondary attack rate among 10-19 year olds (P = 0.32; **Fig. S8**).

#### Misclassification analysis

We estimated regionally-varying background rates of S gene target failure associated with non-VOC 202012/01 variants using testing data provided by Public Health England as follows:

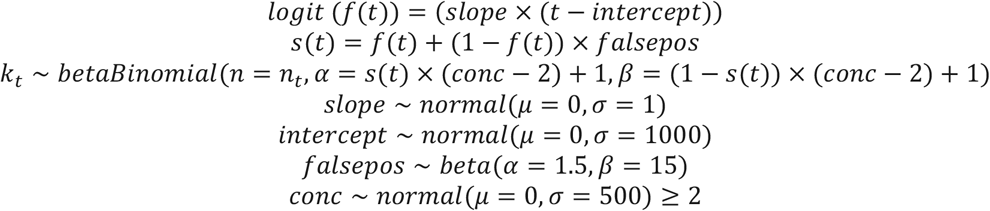

Here, *f*(*t*) is the predicted frequency of VOC 202012/01 among positive tests on day *t* (since 1 September 2020) based on the terms *slope* and *intercept*; *s*(*t*) is the predicted frequency of S gene target failure on day *t* due to the combination of VOC 202012/01 and a background rate of S gene target failure *falsepos, conc* is the “concentration” parameter (= α + β) of a beta distribution with mode *s*(*t*); *k*_*t*_ is the number of S gene target failures detected on day *t*; and *n*_*t*_ is the total number of tests on day *t*. All priors above are chosen to be vague, and the truncation of *conc* to values greater than 2 ensures a unimodal distribution for the proportion of tests with S gene target failure. This model is fitted separately for each NHS England region. Then, the probability that an S gene target failure on day *t* is VOC 202012/01 is estimated as *f* (*t*)/*s* (*t*).

#### Details of Bayesian inference

To fit the dynamic transmission model to data on deaths, hospital admissions, hospital bed and ICU bed occupancy, PCR positivity, and seroprevalence for each of the 7 NHS England regions, we performed Bayesian inference using Markov chain Monte Carlo, employing the Differential Evolution MCMC algorithm (*65*). For each posterior sample, we simulated epidemics from 1 January to 24 December 2020, using data that were current as of 8 January 2021. We used Google Community Mobility data up to 24 December 2020 to capture how interpersonal contact rates changed over the course of the epidemic.

When fitting deaths, hospital admissions, hospital bed occupancy and ICU bed occupancy, we used a negative binomial likelihood with a fitted size parameter for each series and region. For seroprevalence and PCR prevalence, we used a skew-normal likelihood for each data point fitted to produce the same mean and 95% confidence interval as was reported for the data, and took the expected value of the model prediction over the date range during which the prevalence was measured. For fitting to VOC 202012/01 relative frequency over time in the three heavily affected NHS England regions, we used a beta-binomial likelihood with the daily proportion of detected samples that were VOC 202012/01 and a fitted dispersion parameter.

As part of model estimation, we separately fit for each region: the start time of community transmission; the basic reproduction number *R*_*0*_ prior to any changes in mobility or closure of schools; the delay from infection to hospital admission, to ICU admission, and to death; a region-specific relative probability of hospital admission and of ICU admission given infection; the relative infection fatality ratio at the start and at the end of the simulation period, as fatality due to COVID-19 has dropped substantially over time in the UK; a decreasing rate of effective contact between individuals over time, representing better practices of self-isolation and precautions against infection taken by individuals over the course of the year; and coefficients determining the relative mobility of younger people, around age 20, relative to the rest of the population, for the months of July, August, and September onwards. Full details of all fitted parameters, along with prior distributions assumed for each parameter, are in Table S2. Gelman-Rubin convergence diagnostics were all ≤ 1.1 (Table S6).

We use two parametric functions extensively in parameterising the model. The first,

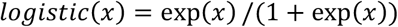

is the standard logistic curve. The second,

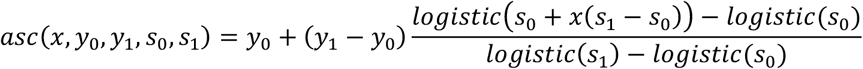

is a logistic-shaped curve parameterised to be a smooth S-shaped function of x from 0 to 1, which goes from y_0_ at x = 0 to y_1_ at x = 1, with an inflection point at x = −s_0_/(−s_0_ + s_1_) if s_0_ < 0 and s_1_ > 0.

Basic epidemiological parameters were broadly informed from the literature and previously reported (*10*). All parameters that we adopted as assumptions are given in Table S3.

## Supplementary Text 1: CMMID COVID-19 Working Group members and funding

The CMMID COVID-19 Working Group is (randomized order): Sophie R Meakin, James D Munday, Amy Gimma, Rosanna C Barnard, Timothy W Russell, Billy J Quilty, Yang Liu, Stefan Flasche, Jiayao Lei, Adam J Kucharski, William Waites, Sebastian Funk, Fiona Yueqian Sun, Fabienne Krauer, Rachel Lowe, Nikos I Bosse, Damien C Tully, Emily S Nightingale, Katharine Sherratt, Rosalind M Eggo, Kaja Abbas, Kathleen O’Reilly, Hamish P Gibbs, C Julian Villabona-Arenas, Naomi R Waterlow, W John Edmunds, Graham Medley, Oliver Brady, Jack Williams, Alicia Rosello, Christopher I Jarvis, Petra Klepac, Mihaly Koltai, Nicholas G. Davies, Frank G Sandmann, Anna M Foss, Sam Abbott, Yalda Jafari, Kiesha Prem, Yung-Wai Desmond Chan, Katherine E. Atkins, Carl A B Pearson, Joel Hellewell, Kevin van Zandvoort, Simon R Procter, Thibaut Jombart, Gwenan M Knight, Akira Endo, Matthew Quaife, Mark Jit, Alicia Showering, Samuel Clifford.

Funding for the CMMID COVID-19 Working Group is as follows. SRM: Wellcome Trust (grant: 210758/Z/18/Z). JDM: Wellcome Trust (grant: 210758/Z/18/Z). AG: European Commission (EpiPose 101003688). RCB: European Commission (EpiPose 101003688). TWR: Wellcome Trust (grant: 206250/Z/17/Z). BJQ: This research was partly funded by the National Institute for Health Research (NIHR) (16/137/109 & 16/136/46) using UK aid from the UK Government to support global health research. The views expressed in this publication are those of the author(s) and not necessarily those of the NIHR or the UK Department of Health and Social Care. BJQ is supported in part by a grant from the Bill and Melinda Gates Foundation (OPP1139859). YL: Bill & Melinda Gates Foundation (INV-003174), NIHR (16/137/109), European Commission (101003688). SFlasche: Wellcome Trust (grant: 208812/Z/17/Z). JYL: Bill & Melinda Gates Foundation (INV-003174). AJK: Wellcome Trust (grant: 206250/Z/17/Z), NIHR (NIHR200908). WW: UK Medical Research Council (MRC) (grant MR/V027956/1). S. Funk: Wellcome Trust (grant: 210758/Z/18/Z), NIHR (NIHR200908). FYS: NIHR EPIC grant (16/137/109). FK: Innovation Fund of the Joint Federal Committee (Grant number 01VSF18015), Wellcome Trust (UNS110424). RL: Royal Society Dorothy Hodgkin Fellowship. NIB: Health Protection Research Unit (grant code NIHR200908). DCT: No funding declared. ESN: Bill & Melinda Gates Foundation (OPP1183986). KS: Wellcome Trust (grant: 210758/Z/18/Z). RME: HDR UK (grant: MR/S003975/1), MRC (grant: MC_PC 19065), NIHR (grant: NIHR200908). KA: Bill & Melinda Gates Foundation (OPP1157270, INV-016832). KO’R: Bill and Melinda Gates Foundation (OPP1191821). HPG: This research was produced by CSIGN which is part of the EDCTP2 programme supported by the European Union (grant number RIA2020EF-2983-CSIGN). The views and opinions of authors expressed herein do not necessarily state or reflect those of EDCTP. This research is funded by the Department of Health and Social Care using UK Aid funding and is managed by the NIHR. The views expressed in this publication are those of the author(s) and not necessarily those of the Department of Health and Social Care (PR-OD-1017-20001). CJVA: European Research Council Starting Grant (Action number 757688). NRW: Medical Research Council (grant number MR/N013638/1). WJE: European Commission (EpiPose 101003688), NIHR (NIHR200908). GFM: NTD Modelling Consortium by the Bill and Melinda Gates Foundation (OPP1184344). OJB: Wellcome Trust (grant: 206471/Z/17/Z). JW: NIHR Health Protection Research Unit and NIHR HTA. AR: NIHR (grant: PR-OD-1017-20002). CIJ: Global Challenges Research Fund (GCRF) project ‘RECAP’ managed through RCUK and ESRC (ES/P010873/1). PK: This research was partly funded by the Royal Society under award RP\EA\180004, European Commission (101003688), Bill & Melinda Gates Foundation (INV-003174). MK: Foreign, Commonwealth and Development Office / Wellcome Trust. NGD: UKRI Research England; NIHR Health Protection Research Unit in Immunisation (NIHR200929); UK MRC (MC_PC_19065). FGS: NIHR Health Protection Research Unit in Modelling & Health Economics, and in Immunisation. AMF: No funding declared. SA: Wellcome Trust (grant: 210758/Z/18/Z). YJ: LSHTM, DHSC/UKRI COVID-19 Rapid Response Initiative. KP: Bill & Melinda Gates Foundation (INV-003174), European Commission (101003688). YWDC: No funding declared. KEA: European Research Council Starting Grant (Action number 757688). CABP: CABP is supported by the Bill & Melinda Gates Foundation (OPP1184344) and the UK Foreign, Commonwealth and Development Office (FCDO)/Wellcome Trust Epidemic Preparedness Coronavirus research programme (ref. 221303/Z/20/Z). JH: Wellcome Trust (grant: 210758/Z/18/Z). KvZ: KvZ is supported by the UK Foreign, Commonwealth and Development Office (FCDO)/Wellcome Trust Epidemic Preparedness Coronavirus research programme (ref. 221303/Z/20/Z), and Elrha’s Research for Health in Humanitarian Crises (R2HC) Programme, which aims to improve health outcomes by strengthening the evidence base for public health interventions in humanitarian crises. The R2HC programme is funded by the UK Government (FCDO), the Wellcome Trust, and the UK National Institute for Health Research (NIHR). SRP: Bill and Melinda Gates Foundation (INV-016832). TJ: RCUK/ESRC (grant: ES/P010873/1); UK PH RST; NIHR HPRU Modelling & Health Economics (NIHR200908). GMK: UK Medical Research Council (grant: MR/P014658/1). AE: The Nakajima Foundation. MQ: European Research Council Starting Grant (Action Number #757699); Bill and Melinda Gates Foundation (INV-001754). MJ: Bill & Melinda Gates Foundation (INV-003174, INV-016832), NIHR (16/137/109, NIHR200929, NIHR200908), European Commission (EpiPose 101003688). AS: No funding declared. SC: Wellcome Trust (grant: 208812/Z/17/Z).

## Supplementary Text 2: COG-UK Consortium members and affiliations

*Funding acquisition, Leadership and supervision, Metadata curation, Project administration, Samples and logistics, Sequencing and analysis, Software and analysis tools, and Visualisation:* Samuel C Robson ^13^.

*Funding acquisition, Leadership and supervision, Metadata curation, Project administration, Samples and logistics, Sequencing and analysis, and Software and analysis tools:* Nicholas J Loman ^41^ and Thomas R Connor ^10, 69^.

*Leadership and supervision, Metadata curation, Project administration, Samples and logistics, Sequencing and analysis, Software and analysis tools, and Visualisation:* Tanya Golubchik ^5^.

*Funding acquisition, Metadata curation, Samples and logistics, Sequencing and analysis, Software and analysis tools, and Visualisation:* Rocio T Martinez Nunez ^42^.

*Funding acquisition, Leadership and supervision, Metadata curation, Project administration, and Samples and logistics:* Catherine Ludden ^88^.

*Funding acquisition, Leadership and supervision, Metadata curation, Samples and logistics, and Sequencing and analysis:* Sally Corden ^69^.

*Funding acquisition, Leadership and supervision, Project administration, Samples and logistics, and Sequencing and analysis:* Ian Johnston ^99^ and David Bonsall ^5^.

*Funding acquisition, Leadership and supervision, Sequencing and analysis, Software and analysis tools, and Visualisation:* Colin P Smith ^87^ and Ali R Awan ^28^.

*Funding acquisition, Samples and logistics, Sequencing and analysis, Software and analysis tools, and Visualisation:* Giselda Bucca ^87^.

*Leadership and supervision, Metadata curation, Project administration, Samples and logistics, and Sequencing and analysis:* M. Estee Torok ^22, 101^.

*Leadership and supervision, Metadata curation, Project administration, Samples and logistics, and Visualisation:* Kordo Saeed ^81, 110^ and Jacqui A Prieto ^83, 109^.

*Leadership and supervision, Metadata curation, Project administration, Sequencing and analysis, and Software and analysis tools:* David K Jackson ^99^.

*Metadata curation, Project administration, Samples and logistics, Sequencing and analysis, and Software and analysis tools:* William L Hamilton ^22^.

*Metadata curation, Project administration, Samples and logistics, Sequencing and analysis, and Visualisation:* Luke B Snell ^11^.

*Funding acquisition, Leadership and supervision, Metadata curation, and Samples and logistics:* Catherine Moore ^69^.

*Funding acquisition, Leadership and supervision, Project administration, and Samples and logistics:* Ewan M Harrison ^99, 88^.

*Leadership and supervision, Metadata curation, Project administration, and Samples and logistics:* Sonia Goncalves ^99^.

*Leadership and supervision, Metadata curation, Samples and logistics, and Sequencing and analysis:* Ian G Goodfellow ^24^, Derek J Fairley ^3, 72^, Matthew W Loose ^18^ and Joanne Watkins ^69^.

*Leadership and supervision, Metadata curation, Samples and logistics, and Software and analysis tools:* Rich Livett ^99^.

*Leadership and supervision, Metadata curation, Samples and logistics, and Visualisation:* Samuel Moses ^25, 106^.

*Leadership and supervision, Metadata curation, Sequencing and analysis, and Software and analysis tools:* Roberto Amato ^99^, Sam Nicholls ^41^ and Matthew Bull ^69^.

*Leadership and supervision, Project administration, Samples and logistics, and Sequencing and analysis:* Darren L Smith ^37, 58, 105^.

*Leadership and supervision, Sequencing and analysis, Software and analysis tools, and Visualisation:* Jeff Barrett ^99^, David M Aanensen ^14, 114^.

*Metadata curation, Project administration, Samples and logistics, and Sequencing and analysis:* Martin D Curran ^65^, Surendra Parmar ^65^, Dinesh Aggarwal ^95, 99, 64^ and James G Shepherd ^48^.

*Metadata curation, Project administration, Sequencing and analysis, and Software and analysis tools:* Matthew D Parker ^93^.

*Metadata curation, Samples and logistics, Sequencing and analysis, and Visualisation:* Sharon Glaysher ^61^.

*Metadata curation, Sequencing and analysis, Software and analysis tools, and Visualisation:* Matthew Bashton ^37, 58^, Anthony P Underwood ^14, 114^, Nicole Pacchiarini ^69^ and Katie F Loveson ^77^.

*Project administration, Sequencing and analysis, Software and analysis tools, and Visualisation:* Alessandro M Carabelli ^88^.

*Funding acquisition, Leadership and supervision, and Metadata curation:* Kate E Templeton ^53, 90^.

*Funding acquisition, Leadership and supervision, and Project administration:* Cordelia F Langford ^99^, John Sillitoe ^99^, Thushan I de Silva ^93^ and Dennis Wang ^93^.

*Funding acquisition, Leadership and supervision, and Sequencing and analysis:* Dominic Kwiatkowski ^99, 107^, Andrew Rambaut ^90^, Justin O’Grady ^70, 89^ and Simon Cottrell ^69^.

*Leadership and supervision, Metadata curation, and Sequencing and analysis:* Matthew T.G. Holden ^68^ and Emma C Thomson ^48^.

*Leadership and supervision, Project administration, and Samples and logistics:* Husam Osman ^64, 36^, Monique Andersson ^59^, Anoop J Chauhan ^61^ and Mohammed O Hassan-Ibrahim ^6^.

*Leadership and supervision, Project administration, and Sequencing and analysis:* Mara Lawniczak ^99^.

*Leadership and supervision, Samples and logistics, and Sequencing and analysis:* Ravi Kumar Gupta ^88, 113^, Alex Alderton ^99^, Meera Chand ^66^, Chrystala Constantinidou ^94^, Meera Unnikrishnan ^94^, Alistair C Darby ^92^, Julian A Hiscox ^92^ and Steve Paterson ^92^.

*Leadership and supervision, Sequencing and analysis, and Software and analysis tools:* Inigo Martincorena ^99^, David L Robertson ^48^, Erik M Volz ^39^, Andrew J Page ^70^ and Oliver G Pybus ^23^.

*Leadership and supervision, Sequencing and analysis, and Visualisation:* Andrew R Bassett^99^.

*Metadata curation, Project administration, and Samples and logistics:* Cristina V Ariani ^99^, Michael H Spencer Chapman ^99, 88^, Kathy K Li ^48^, Rajiv N Shah ^48^, Natasha G Jesudason ^48^ and Yusri Taha ^50^.

*Metadata curation, Project administration, and Sequencing and analysis:* Martin P McHugh ^53^ and Rebecca Dewar ^53^.

*Metadata curation, Samples and logistics, and Sequencing and analysis:* Aminu S Jahun ^24^, Claire McMurray ^41^, Sarojini Pandey ^84^, James P McKenna ^3^, Andrew Nelson ^58, 105^, Gregory R Young ^37, 58^, Clare M McCann ^58, 105^ and Scott Elliott ^61^.

*Metadata curation, Samples and logistics, and Visualisation:* Hannah Lowe ^25^.

*Metadata curation, Sequencing and analysis, and Software and analysis tools:* Ben Temperton ^91^, Sunando Roy ^82^, Anna Price ^10^, Sara Rey ^69^ and Matthew Wyles ^93^.

*Metadata curation, Sequencing and analysis, and Visualisation:* Stefan Rooke ^90^ and Sharif Shaaban ^68^.

*Project administration, Samples and logistics, Sequencing and analysis:* Mariateresa de Cesare ^98^.

*Project administration, Samples and logistics, and Software and analysis tools:* Laura Letchford ^99^.

*Project administration, Samples and logistics, and Visualisation:* Siona Silveira ^81^, Emanuela Pelosi ^81^ and Eleri Wilson-Davies ^81^.

*Samples and logistics, Sequencing and analysis, and Software and analysis tools:* Myra Hosmillo ^24^.

*Sequencing and analysis, Software and analysis tools, and Visualisation:* Áine O’Toole ^90^, Andrew R Hesketh ^87^, Richard Stark ^94^, Louis du Plessis ^23^, Chris Ruis ^88^, Helen Adams ^4^ and Yann Bourgeois ^76^.

*Funding acquisition, and Leadership and supervision:* Stephen L Michell ^91^, Dimitris Grammatopoulos^84, 112^, Jonathan Edgeworth ^12^, Judith Breuer ^30, 82^, John A Todd ^98^ and Christophe Fraser ^5^.

*Funding acquisition, and Project administration:* David Buck ^98^ and Michaela John ^9^.

*Leadership and supervision, and Metadata curation:* Gemma L Kay ^70^.

*Leadership and supervision, and Project administration:* Steve Palmer ^99^, Sharon J Peacock ^88, 64^ and David Heyburn ^69^.

*Leadership and supervision, and Samples and logistics:* Danni Weldon ^99^, Esther Robinson ^64, 36^, Alan McNally ^41, 86^, Peter Muir ^64^, Ian B Vipond ^64^, John BoYes ^29^, Venkat Sivaprakasam ^46^, Tranprit Salluja ^75^, Samir Dervisevic ^54^ and Emma J Meader ^54^.

*Leadership and supervision, and Sequencing and analysis:* Naomi R Park ^99^, Karen Oliver ^99^, Aaron R Jeffries ^91^, Sascha Ott ^94^, Ana da Silva Filipe ^48^, David A Simpson ^72^ and Chris Williams ^69^.

*Leadership and supervision, and Visualisation:* Jane AH Masoli ^73, 91^.

*Metadata curation, and Samples and logistics:* Bridget A Knight ^73, 91^, Christopher R Jones ^73, 91^, Cherian Koshy ^1^, Amy Ash ^1^, Anna Casey ^71^, Andrew Bosworth ^64, 36^, Liz Ratcliffe ^71^, Li Xu-McCrae ^36^, Hannah M Pymont ^64^, Stephanie Hutchings ^64^, Lisa Berry ^84^, Katie Jones ^84^, Fenella Halstead ^46^, Thomas Davis ^21^, Christopher Holmes ^16^, Miren Iturriza-Gomara ^92^, Anita O Lucaci ^92^, Paul Anthony Randell ^38, 104^, Alison Cox ^38, 104^, Pinglawathee Madona ^38, 104^, Kathryn Ann Harris ^30^, Julianne Rose Brown ^30^, Tabitha W Mahungu ^74^, Dianne Irish-Tavares ^74^, Tanzina Haque ^74^, Jennifer Hart ^74^, Eric Witele ^74^, Melisa Louise Fenton ^75^, Steven Liggett ^79^, Clive Graham ^56^, Emma Swindells ^57^, Jennifer Collins ^50^, Gary Eltringham ^50^, Sharon Campbell ^17^, Patrick C McClure ^97^, Gemma Clark ^15^, Tim J Sloan ^60^, Carl Jones ^15^ and Jessica Lynch ^2, 111^.

*Metadata curation, and Sequencing and analysis:* Ben Warne ^8^, Steven Leonard ^99^, Jillian Durham ^99^, Thomas Williams ^90^, Sam T Haldenby ^92^, Nathaniel Storey ^30^, Nabil-Fareed Alikhan ^70^, Nadine Holmes ^18^, Christopher Moore ^18^, Matthew Carlile ^18^, Malorie Perry ^69^, Noel Craine ^69^, Ronan A Lyons ^80^, Angela H Beckett ^13^, Salman Goudarzi ^77^, Christopher Fearn ^77^, Kate Cook ^77^, Hannah Dent ^77^ and Hannah Paul ^77^.

*Metadata curation, and Software and analysis tools:* Robert Davies ^99^.

*Project administration, and Samples and logistics:* Beth Blane ^88^, Sophia T Girgis ^88^, Mathew A Beale ^99^, Katherine L Bellis ^99, 88^, Matthew J Dorman ^99^, Eleanor Drury ^99^, Leanne Kane ^99^, Sally Kay ^99^, Samantha McGuigan ^99^, Rachel Nelson ^99^, Liam Prestwood ^99^, Shavanthi Rajatileka ^99^, Rahul Batra ^12^, Rachel J Williams ^82^, Mark Kristiansen ^82^, Angie Green ^98^, Anita Justice ^59^, Adhyana I.K Mahanama ^81, 102^ and Buddhini Samaraweera ^81, 102^.

*Project administration, and Sequencing and analysis:* Nazreen F Hadjirin ^88^ and Joshua Quick ^41^.

*Project administration, and Software and analysis tools:* Radoslaw Poplawski ^41^.

*Samples and logistics, and Sequencing and analysis:* Leanne M Kermack ^88^, Nicola Reynolds ^7^, Grant Hall ^24^, Yasmin Chaudhry ^24^, Malte L Pinckert ^24^, Iliana Georgana ^24^, Robin J Moll ^99^, Alicia Thornton ^66^, Richard Myers ^66^, Joanne Stockton ^41^, Charlotte A Williams ^82^, Wen C Yew ^58^, Alexander J Trotter ^70^, Amy Trebes ^98^, George MacIntyre-Cockett ^98^, Alec Birchley ^69^, Alexander Adams ^69^, Amy Plimmer ^69^, Bree Gatica-Wilcox ^69^, Caoimhe McKerr ^69^, Ember Hilvers ^69^, Hannah Jones ^69^, Hibo Asad ^69^, Jason Coombes ^69^, Johnathan M Evans ^69^, Laia Fina ^69^, Lauren Gilbert ^69^, Lee Graham ^69^, Michelle Cronin ^69^, Sara Kumziene-SummerhaYes ^69^, Sarah Taylor ^69^, Sophie Jones ^69^, Danielle C Groves ^93^, Peijun Zhang ^93^, Marta Gallis ^93^ and Stavroula F Louka ^93^.

*Samples and logistics, and Software and analysis tools:* Igor Starinskij ^48^.

*Sequencing and analysis, and Software and analysis tools:* Chris J Illingworth ^47^, Chris Jackson ^47^, Marina Gourtovaia ^99^, Gerry Tonkin-Hill ^99^, Kevin Lewis ^99^, Jaime M Tovar-Corona ^99^, Keith James ^99^, Laura Baxter ^94^, Mohammad T. Alam ^94^, Richard J Orton ^48^, Joseph Hughes ^48^, Sreenu Vattipally ^48^, Manon Ragonnet-Cronin ^39^, Fabricia F. Nascimento ^39^, David Jorgensen ^39^, Olivia Boyd ^39^, Lily Geidelberg ^39^, Alex E Zarebski ^23^, Jayna Raghwani ^23^, Moritz UG Kraemer ^23^, Joel Southgate ^10, 69^, Benjamin B Lindsey ^93^ and Timothy M Freeman ^93^.

*Software and analysis tools, and Visualisation:* Jon-Paul Keatley ^99^, Joshua B Singer ^48^, Leonardo de Oliveira Martins ^70^, Corin A Yeats ^14^, Khalil Abudahab ^14, 114^, Ben EW Taylor ^14, 114^ and Mirko Menegazzo ^14^.

*Leadership and supervision:* John Danesh ^99^, Wendy Hogsden ^46^, Sahar Eldirdiri ^21^, Anita Kenyon ^21^, Jenifer Mason ^43^, Trevor I Robinson ^43^, Alison Holmes ^38, 103^, James Price ^38, 103^, John A Hartley ^82^, Tanya Curran ^3^, Alison E Mather ^70^, Giri Shankar ^69^, Rachel Jones ^69^, Robin Howe ^69^ and Sian Morgan ^9^.

*Metadata curation:* Elizabeth Wastenge ^53^, Michael R Chapman ^34, 88, 99^, Siddharth Mookerjee ^38, 103^, Rachael Stanley ^54^, Wendy Smith ^15^, Timothy Peto ^59^, David Eyre ^59^, Derrick Crook ^59^, Gabrielle Vernet ^33^, Christine Kitchen ^10^, Huw Gulliver ^10^, Ian Merrick ^10^, Martyn Guest ^10^, Robert Munn ^10^, Declan T Bradley ^63, 72^ and Tim Wyatt ^63^.

*Project administration:* Charlotte Beaver ^99^, Luke Foulser ^99^, Sophie Palmer ^88^, Carol M Churcher ^88^, Ellena Brooks ^88^, Kim S Smith ^88^, Katerina Galai ^88^, Georgina M McManus ^88^, Frances Bolt ^38, 103^, Francesc Coll ^19^, Lizzie Meadows ^70^, Stephen W Attwood ^23^, Alisha Davies ^69^, Elen De Lacy ^69^, Fatima Downing ^69^, Sue Edwards ^69^, Garry P Scarlett ^76^, Sarah Jeremiah ^83^ and Nikki Smith ^93^.

*Samples and logistics:* Danielle Leek ^88^, Sushmita Sridhar ^88, 99^, Sally Forrest ^88^, Claire Cormie ^88^, Harmeet K Gill ^88^, Joana Dias ^88^, Ellen E Higginson ^88^, Mailis Maes ^88^, Jamie Young ^88^, Michelle Wantoch ^7^, Sanger Covid Team (www.sanger.ac.uk/covid-team) ^99^, Dorota Jamrozy ^99^, Stephanie Lo ^99^, Minal Patel ^99^, Verity Hill ^90^, Claire M Bewshea ^91^, Sian Ellard ^73, 91^, Cressida Auckland ^73^, Ian Harrison ^66^, Chloe Bishop ^66^, Vicki Chalker ^66^, Alex Richter ^85^, Andrew Beggs ^85^, Angus Best ^86^, Benita Percival ^86^, Jeremy Mirza ^86^, Oliver Megram ^86^, Megan Mayhew ^86^, Liam Crawford ^86^, Fiona Ashcroft ^86^, Emma Moles-Garcia ^86^, Nicola Cumley ^86^, Richard Hopes ^64^, Patawee Asamaphan ^48^, Marc O Niebel ^48^, Rory N Gunson ^100^, Amanda Bradley ^52^, Alasdair Maclean ^52^, Guy Mollett ^52^, Rachel Blacow ^52^, Paul Bird ^16^, Thomas Helmer ^16^, Karlie Fallon ^16^, Julian Tang ^16^, Antony D Hale ^49^, Louissa R Macfarlane-Smith ^49^, Katherine L Harper ^49^, Holli Carden ^49^, Nicholas W Machin ^45, 64^, Kathryn A Jackson ^92^, Shazaad S Y Ahmad ^45, 64^, Ryan P George ^45^, Lance Turtle ^92^, Elaine O’Toole ^43^, Joanne Watts ^43^, Cassie Breen ^43^, Angela Cowell ^43^, Adela Alcolea-Medina ^32, 96^, Themoula Charalampous ^12, 42^, Amita Patel ^11^, Lisa J Levett ^35^, Judith Heaney ^35^, Aileen Rowan ^39^, Graham P Taylor ^39^, Divya Shah ^30^, Laura Atkinson ^30^, Jack CD Lee ^30^, Adam P Westhorpe ^82^, Riaz Jannoo ^82^, Helen L Lowe ^82^, Angeliki Karamani ^82^, Leah Ensell ^82^, Wendy Chatterton ^35^, Monika Pusok ^35^, Ashok Dadrah ^75^, Amanda Symmonds ^75^, Graciela Sluga ^44^, Zoltan Molnar ^72^, Paul Baker ^79^, Stephen Bonner ^79^, Sarah Essex ^79^, Edward Barton ^56^, Debra Padgett ^56^, Garren Scott ^56^, Jane Greenaway ^57^, Brendan AI Payne ^50^, Shirelle Burton-Fanning ^50^, Sheila Waugh ^50^, Veena Raviprakash ^17^, Nicola Sheriff ^17^, Victoria Blakey ^17^, Lesley-Anne Williams ^17^, Jonathan Moore ^27^, Susanne Stonehouse ^27^, Louise Smith ^55^, Rose K Davidson ^89^, Luke Bedford ^26^, Lindsay Coupland ^54^, Victoria Wright ^18^, Joseph G Chappell ^97^, Theocharis Tsoleridis ^97^, Jonathan Ball ^97^, Manjinder Khakh ^15^, Vicki M Fleming ^15^, Michelle M Lister ^15^, Hannah C Howson-Wells ^15^, Louise Berry ^15^, Tim Boswell ^15^, Amelia Joseph ^15^, Iona Willingham ^15^, Nichola Duckworth ^60^, Sarah Walsh ^60^, Emma Wise ^2, 111^, Nathan Moore ^2, 111^, Matilde Mori ^2, 108, 111^, Nick Cortes ^2, 111^, Stephen Kidd ^2, 111^, Rebecca Williams ^33^, Laura Gifford ^69^, Kelly Bicknell ^61^, Sarah Wyllie ^61^, Allyson Lloyd ^61^, Robert Impey ^61^, Cassandra S Malone ^6^, Benjamin J Cogger ^6^, Nick Levene ^62^, Lynn Monaghan ^62^, Alexander J Keeley ^93^, David G Partridge ^78, 93^, Mohammad Raza ^78, 93^, Cariad Evans ^78, 93^ and Kate Johnson ^78, 93^.

*Sequencing and analysis:* Emma Betteridge ^99^, Ben W Farr ^99^, Scott Goodwin ^99^, Michael A Quail ^99^, Carol Scott ^99^, Lesley Shirley ^99^, Scott AJ Thurston ^99^, Diana Rajan ^99^, Iraad F Bronner ^99^, Louise Aigrain ^99^, Nicholas M Redshaw ^99^, Stefanie V Lensing ^99^, Shane McCarthy ^99^, Alex Makunin ^99^, Carlos E Balcazar ^90^, Michael D Gallagher ^90^, Kathleen A Williamson ^90^, Thomas D Stanton ^90^, Michelle L Michelsen ^91^, Joanna Warwick-Dugdale ^91^, Robin Manley ^91^, Audrey Farbos ^91^, James W Harrison ^91^, Christine M Sambles ^91^, David J Studholme ^91^, Angie Lackenby ^66^, Tamyo Mbisa ^66^, Steven Platt ^66^, Shahjahan Miah ^66^, David Bibby ^66^, Carmen Manso ^66^, Jonathan Hubb ^66^, Gavin Dabrera ^66^, Mary Ramsay ^66^, Daniel Bradshaw ^66^, Ulf Schaefer ^66^, Natalie Groves ^66^, Eileen Gallagher ^66^, David Lee ^66^, David Williams ^66^, Nicholas Ellaby ^66^, Hassan Hartman ^66^, Nikos Manesis ^66^, Vineet Patel ^66^, Juan Ledesma ^67^, Katherine A Twohig ^67^, Elias Allara ^64, 88^, Clare Pearson ^64, 88^, Jeffrey K. J. Cheng ^94^, Hannah E. Bridgewater ^94^, Lucy R. Frost ^94^, Grace Taylor-Joyce ^94^, Paul E Brown ^94^, Lily Tong ^48^, Alice Broos ^48^, Daniel Mair ^48^, Jenna Nichols ^48^, Stephen N Carmichael ^48^, Katherine L Smollett ^40^, Kyriaki Nomikou ^48^, Elihu Aranday-Cortes ^48^, Natasha Johnson ^48^, Seema Nickbakhsh ^48, 68^, Edith E Vamos ^92^, Margaret Hughes ^92^, Lucille Rainbow ^92^, Richard Eccles ^92^, Charlotte Nelson ^92^, Mark Whitehead ^92^, Richard Gregory ^92^, Matthew Gemmell ^92^, Claudia Wierzbicki ^92^, Hermione J Webster ^92^, Chloe L Fisher ^28^, Adrian W Signell ^20^, Gilberto Betancor ^20^, Harry D Wilson ^20^, Gaia Nebbia ^12^, Flavia Flaviani ^31^, Alberto C Cerda ^96^, Tammy V Merrill ^96^, Rebekah E Wilson ^96^, Marius Cotic ^82^, Nadua Bayzid ^82^, Thomas Thompson ^72^, Erwan Acheson ^72^, Steven Rushton ^51^, Sarah O’Brien ^51^, David J Baker ^70^, Steven Rudder ^70^, Alp Aydin ^70^, Fei Sang ^18^, Johnny Debebe ^18^, Sarah Francois ^23^, Tetyana I Vasylyeva ^23^, Marina Escalera Zamudio ^23^, Bernardo Gutierrez ^23^, Angela Marchbank ^10^, Joshua Maksimovic ^9^, Karla Spellman ^9^, Kathryn McCluggage ^9^, Mari Morgan ^69^, Robert Beer ^9^, Safiah Afifi ^9^, Trudy Workman ^10^, William Fuller ^10^, Catherine Bresner ^10^, Adrienn Angyal ^93^, Luke R Green ^93^, Paul J Parsons ^93^, Rachel M Tucker ^93^, Rebecca Brown ^93^ and Max Whiteley ^93^.

*Software and analysis tools:* James Bonfield ^99^, Christoph Puethe ^99^, Andrew Whitwham ^99^, Jennifier Liddle ^99^, Will Rowe ^41^, Igor Siveroni ^39^, Thanh Le-Viet ^70^ and Amy Gaskin ^69^.

*Visualisation:* Rob Johnson ^39^.

## Affiliations

**1** Barking, Havering and Redbridge University Hospitals NHS Trust, **2** Basingstoke Hospital, **3** Belfast Health & Social Care Trust, **4** Betsi Cadwaladr University Health Board, **5** Big Data Institute, Nuffield Department of Medicine, University of Oxford, **6** Brighton and Sussex University Hospitals NHS Trust, **7** Cambridge Stem Cell Institute, University of Cambridge, **8** Cambridge University Hospitals NHS Foundation Trust, **9** Cardiff and Vale University Health Board, **10** Cardiff University, **11** Centre for Clinical Infection & Diagnostics Research, St. Thomas’ Hospital and Kings College London, **12** Centre for Clinical Infection and Diagnostics Research, Department of Infectious Diseases, Guy’s and St Thomas’ NHS Foundation Trust, **13** Centre for Enzyme Innovation, University of Portsmouth (PORT), **14** Centre for Genomic Pathogen Surveillance, University of Oxford, **15** Clinical Microbiology Department, Queens Medical Centre, **16** Clinical Microbiology, University Hospitals of Leicester NHS Trust, **17** County Durham and Darlington NHS Foundation Trust, **18** Deep Seq, School of Life Sciences, Queens Medical Centre, University of Nottingham, **19** Department of Infection Biology, Faculty of Infectious & Tropical Diseases, London School of Hygiene & Tropical Medicine, **20** Department of Infectious Diseases, King’s College London, **21** Department of Microbiology, Kettering General Hospital, **22** Departments of Infectious Diseases and Microbiology, Cambridge University Hospitals NHS Foundation Trust; Cambridge, UK, **23** Department of Zoology, University of Oxford, **24** Division of Virology, Department of Pathology, University of Cambridge, **25** East Kent Hospitals University NHS Foundation Trust, **26** East Suffolk and North Essex NHS Foundation Trust, **27** Gateshead Health NHS Foundation Trust, **28** Genomics Innovation Unit, Guy’s and St. Thomas’ NHS Foundation Trust, **29** Gloucestershire Hospitals NHS Foundation Trust, **30** Great Ormond Street Hospital for Children NHS Foundation Trust, **31** Guy’s and St. Thomas’ BRC, **32** Guy’s and St. Thomas’ Hospitals, **33** Hampshire Hospitals NHS Foundation Trust, **34** Health Data Research UK Cambridge, **35** Health Services Laboratories, **36** Heartlands Hospital, Birmingham, **37** Hub for Biotechnology in the Built Environment, Northumbria University, **38** Imperial College Hospitals NHS Trust, **39** Imperial College London, **40** Institute of Biodiversity, Animal Health & Comparative Medicine, **41** Institute of Microbiology and Infection, University of Birmingham, **42** King’s College London, **43** Liverpool Clinical Laboratories, **44** Maidstone and Tunbridge Wells NHS Trust, **45** Manchester University NHS Foundation Trust, **46** Microbiology Department, Wye Valley NHS Trust, Hereford, **47** MRC Biostatistics Unit, University of Cambridge, **48** MRC-University of Glasgow Centre for Virus Research, **49** National Infection Service, PHE and Leeds Teaching Hospitals Trust, **50** Newcastle Hospitals NHS Foundation Trust, **51** Newcastle University, **52** NHS Greater Glasgow and Clyde, **53** NHS Lothian, **54** Norfolk and Norwich University Hospital, **55** Norfolk County Council, **56** North Cumbria Integrated Care NHS Foundation Trust, **57** North Tees and Hartlepool NHS Foundation Trust, **58** Northumbria University, **59** Oxford University Hospitals NHS Foundation Trust, **60** PathLinks, Northern Lincolnshire & Goole NHS Foundation Trust, **61** Portsmouth Hospitals University NHS Trust, **62** Princess Alexandra Hospital Microbiology Dept., **63** Public Health Agency, **64** Public Health England, **65** Public Health England, Clinical Microbiology and Public Health Laboratory, Cambridge, UK, **66** Public Health England, Colindale, **67** Public Health England, Colindale, **68** Public Health Scotland, **69** Public Health Wales NHS Trust, **70** Quadram Institute Bioscience, **71** Queen Elizabeth Hospital, **72** Queen’s University Belfast, **73** Royal Devon and Exeter NHS Foundation Trust, **74** Royal Free NHS Trust, **75** Sandwell and West Birmingham NHS Trust, **76** School of Biological Sciences, University of Portsmouth (PORT), **77** School of Pharmacy and Biomedical Sciences, University of Portsmouth (PORT), **78** Sheffield Teaching Hospitals, **79** South Tees Hospitals NHS Foundation Trust, **80** Swansea University, **81** University Hospitals Southampton NHS Foundation Trust, **82** University College London, **83** University Hospital Southampton NHS Foundation Trust, **84** University Hospitals Coventry and Warwickshire, **85** University of Birmingham, **86** University of Birmingham Turnkey Laboratory, **87** University of Brighton, **88** University of Cambridge, **89** University of East Anglia, **90** University of Edinburgh, **91** University of Exeter, **92** University of Liverpool, **93** University of Sheffield, **94** University of Warwick, **95** University of Cambridge, **96** Viapath, Guy’s and St Thomas’ NHS Foundation Trust, and King’s College Hospital NHS Foundation Trust, **97** Virology, School of Life Sciences, Queens Medical Centre, University of Nottingham, **98** Wellcome Centre for Human Genetics, Nuffield Department of Medicine, University of Oxford, **99** Wellcome Sanger Institute, **100** West of Scotland Specialist Virology Centre, NHS Greater Glasgow and Clyde, **101** Department of Medicine, University of Cambridge, **102** Ministry of Health, Sri Lanka, **103** NIHR Health Protection Research Unit in HCAI and AMR, Imperial College London, **104** North West London Pathology, **105** NU-OMICS, Northumbria University, **106** University of Kent, **107** University of Oxford, **108** University of Southampton, **109** University of Southampton School of Health Sciences, **110** University of Southampton School of Medicine, **111** University of Surrey, **112** Warwick Medical School and Institute of Precision Diagnostics, Pathology, UHCW NHS Trust, **113** Wellcome Africa Health Research Institute Durban and **114** Wellcome Genome Campus.

## Supplementary Figures

**Fig. S1.**
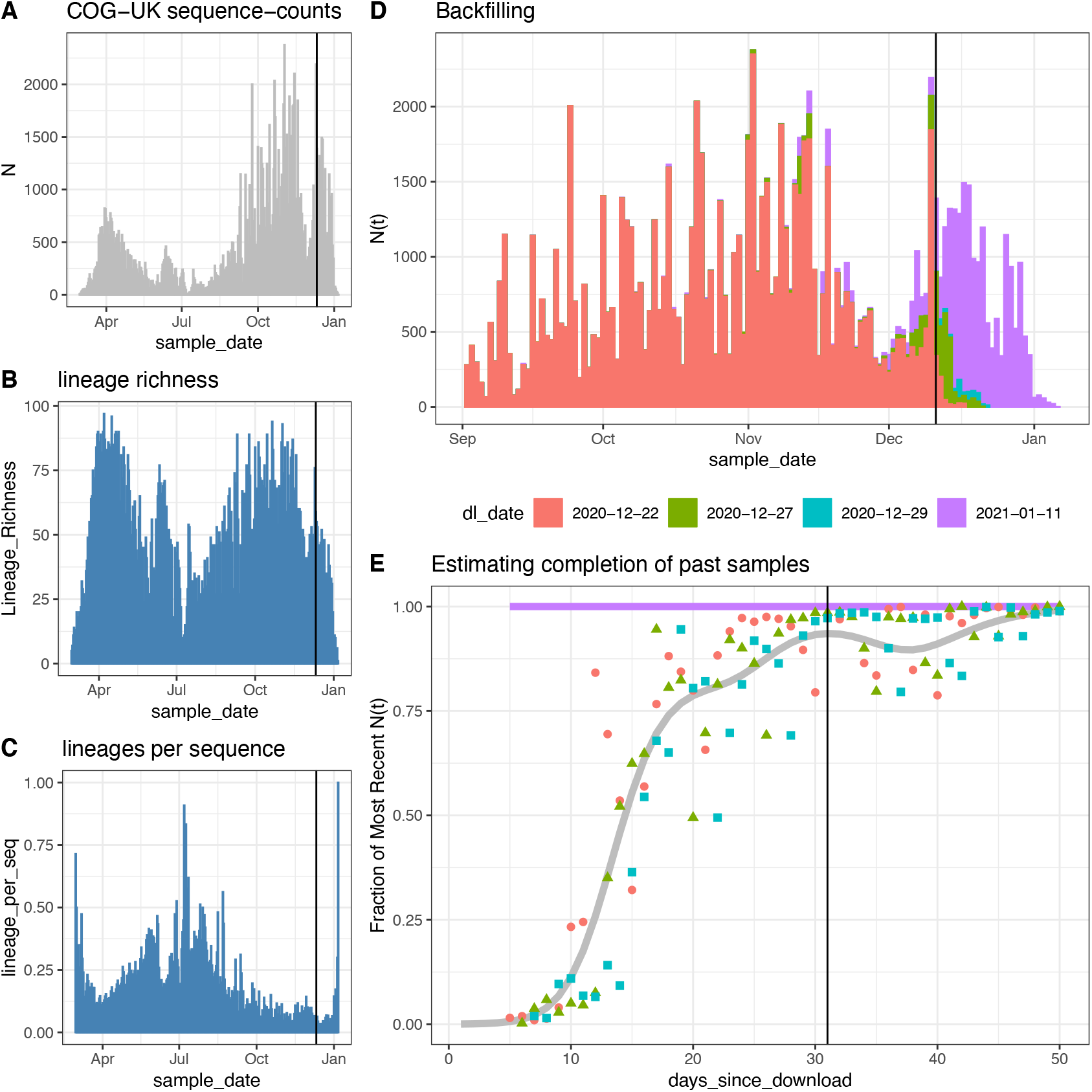
Analysis of COG-UK backfilling. This plot shows the trends in COG-UK sequence counts (A), lineage richness (B), and lineages per-sequence (C) for data downloaded on 2021-01-11. Comparing January 11th download to previous downloads reveals the backfilling of samples from previous sample dates but (D) by 1-month or 31 days prior to a download most of the samples are processed and uploaded to COG-UK. We use data up to 31 days prior to our final download to avoid potential backfilling biases when comparing growth rates across lineages.

**Fig. S2.**
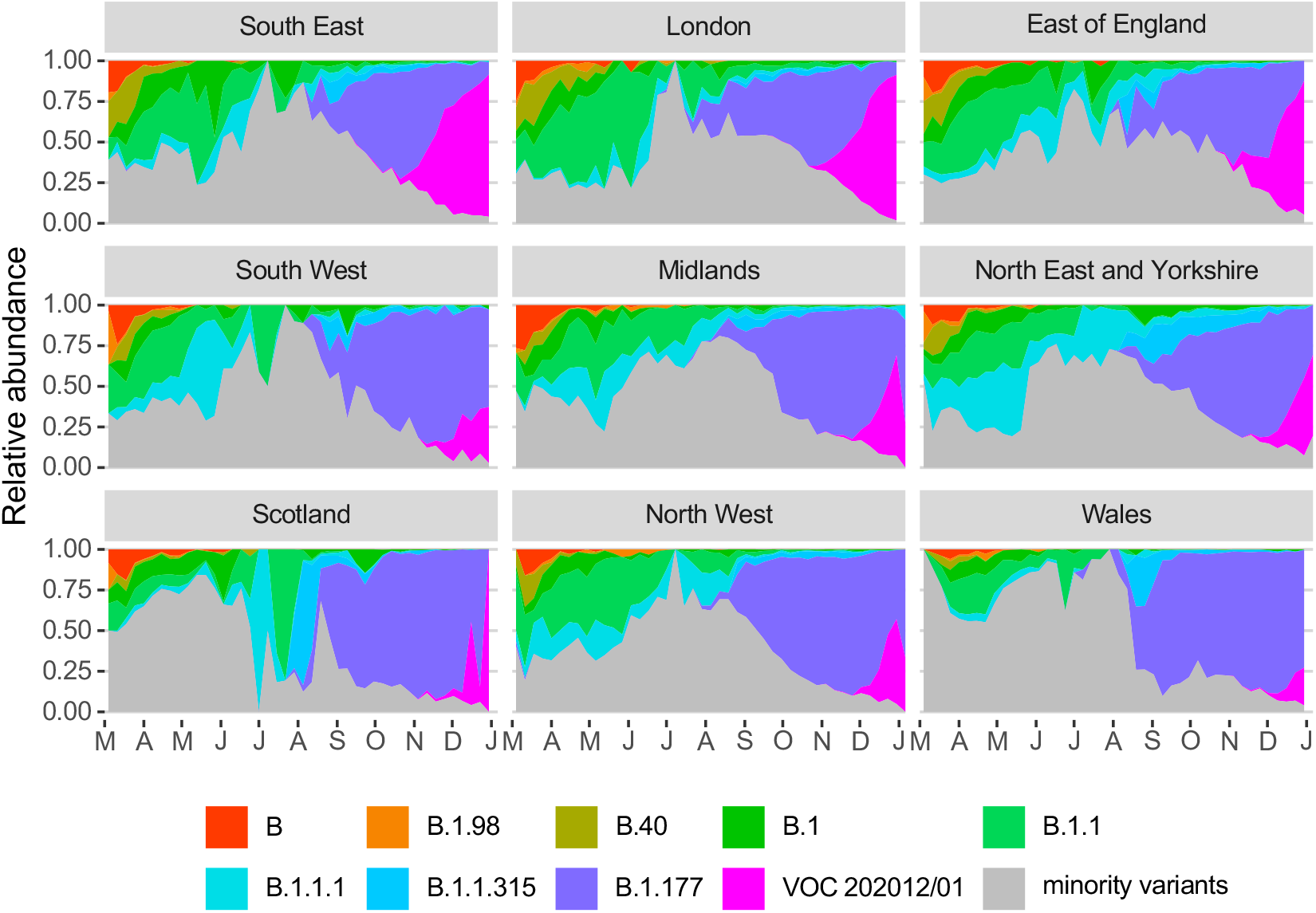
Muller plots of the relative abundance of the 9 major SARS-CoV2 lineages (reaching at least 13% in any week overall) in different NHS regions across the UK, based on the raw COG-UK sequencing data, aggregated by week. The remaining minority variants comprise a collection of a total of 410 lineages. Note that the large fluctuations seen in July & August in some regions such as Scotland are caused by low sample size.

**Fig. S3.**
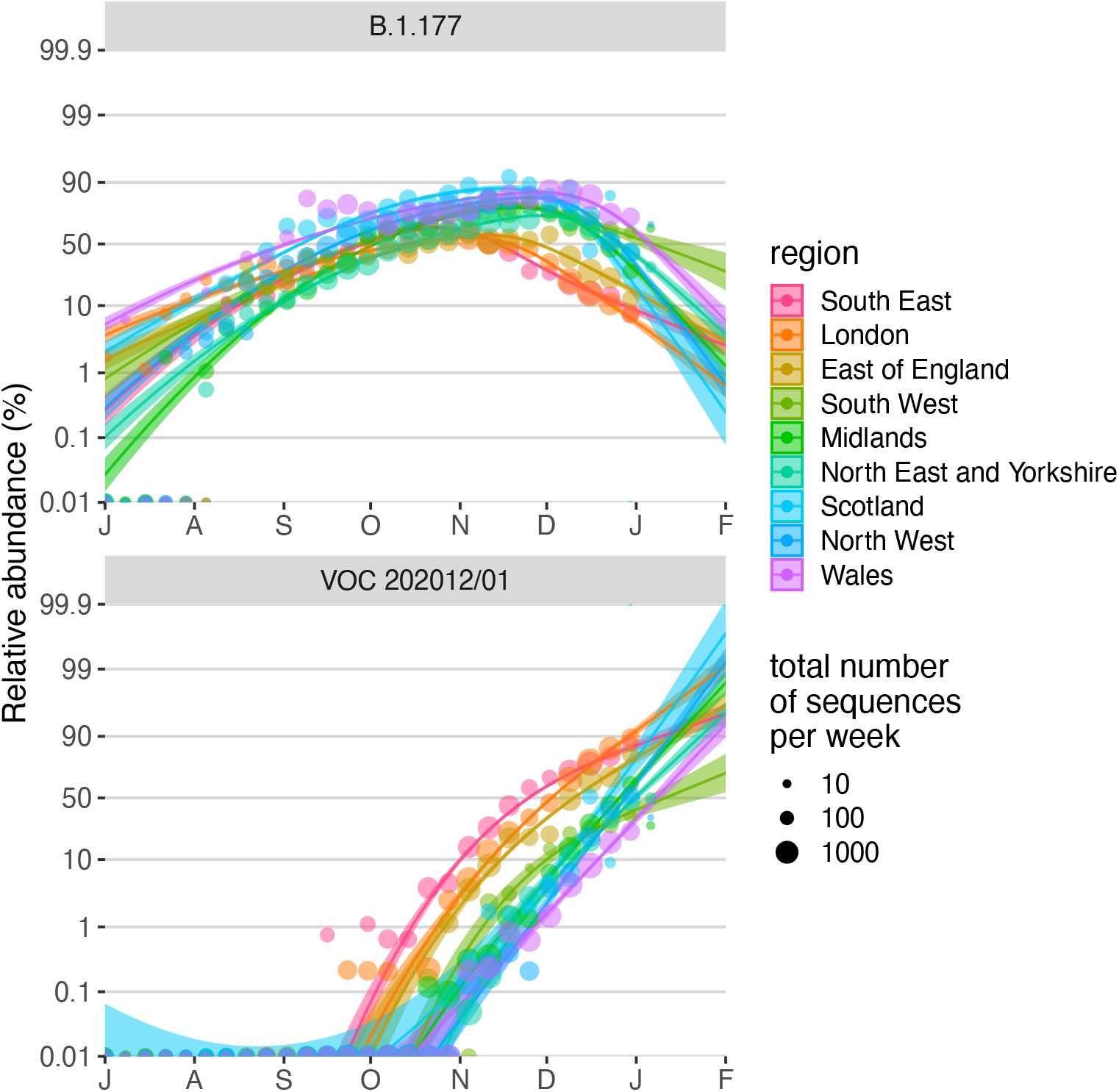
Fitted spread of variants to B.1.177 and VOC 202012/01 estimated from a multinomial spline model by NHS region fit on the COG-UK data (model 1a in Table S1 and Fig. 2C) with 95% confidence intervals and per-week aggregated raw proportions, shown on a logit (log(odds)) scale. The much faster rate of spread of VOC 202012/01 compared to B.1.177 is apparent (cf. Δr values in Table S1). The excellent linearity on a logit scale for VOC 202012/01 allows us to realistically model the spread of this variant using spatially more fine-grained binomial GLMMs (carried out the level of LTLAs), using a subset of the data from August 1 2020 onwards. Likewise, a binomial GLMM was used to model the spread of variant B.1.177 for the period between July 1 2020 and September 30 2020, before it starting to be displaced by VOC 202012/01.

**Fig. S4.**
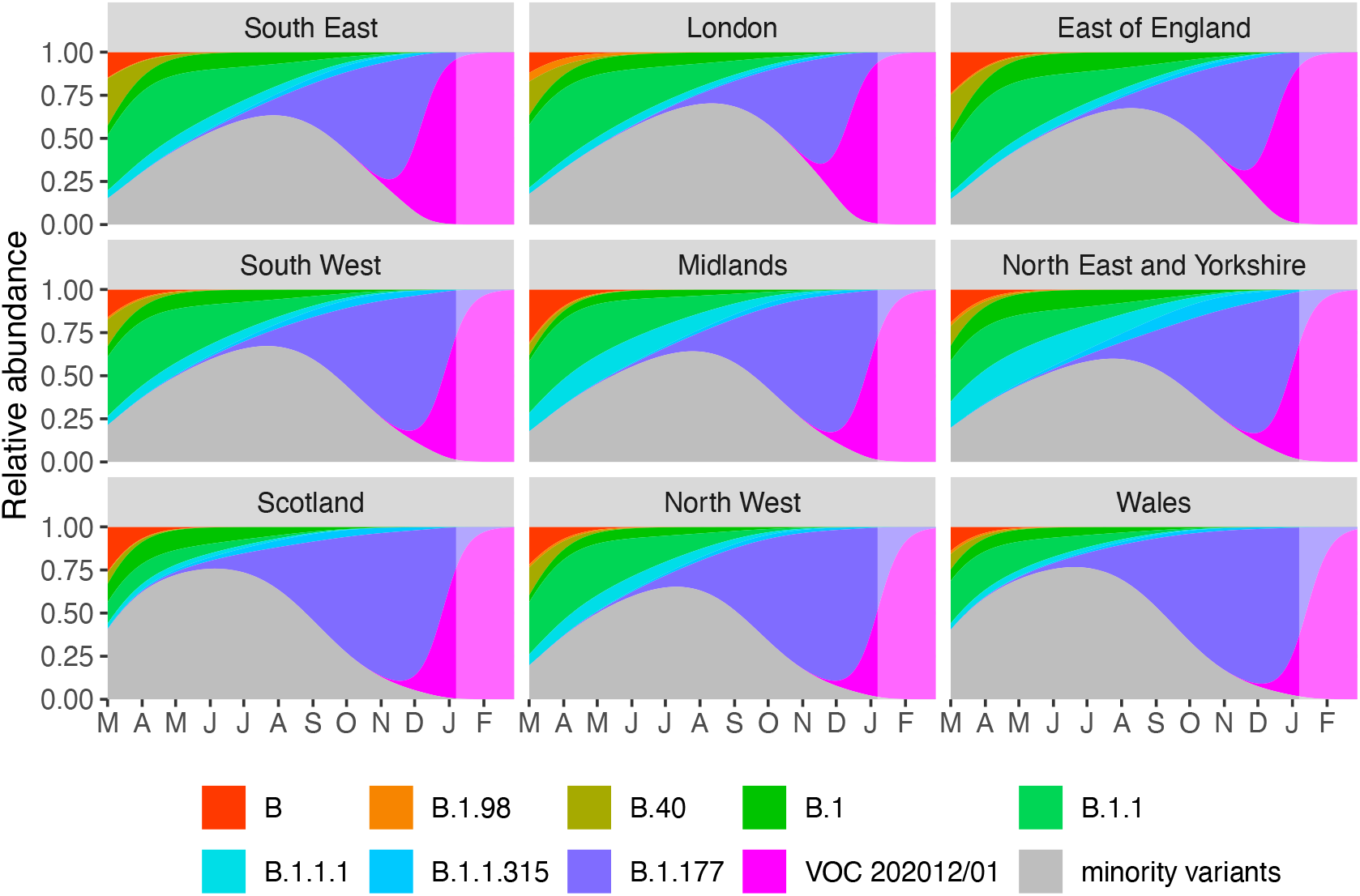
Muller plots of the relative abundances of the major SARS-CoV-2 variants in the UK, based on a multinomial mixed model fit to COG-UK sequence data, incorporating lower-tier local authority as a random intercept as well as overdispersion (common-slopes multinomial mixed model 1b in Table 1). A model extrapolation until the end of February is shown (shaded area). Minority variants are 410 circulating SARS-CoV-2 strains that never reached >13% in any week overall.

**Fig. S5.**
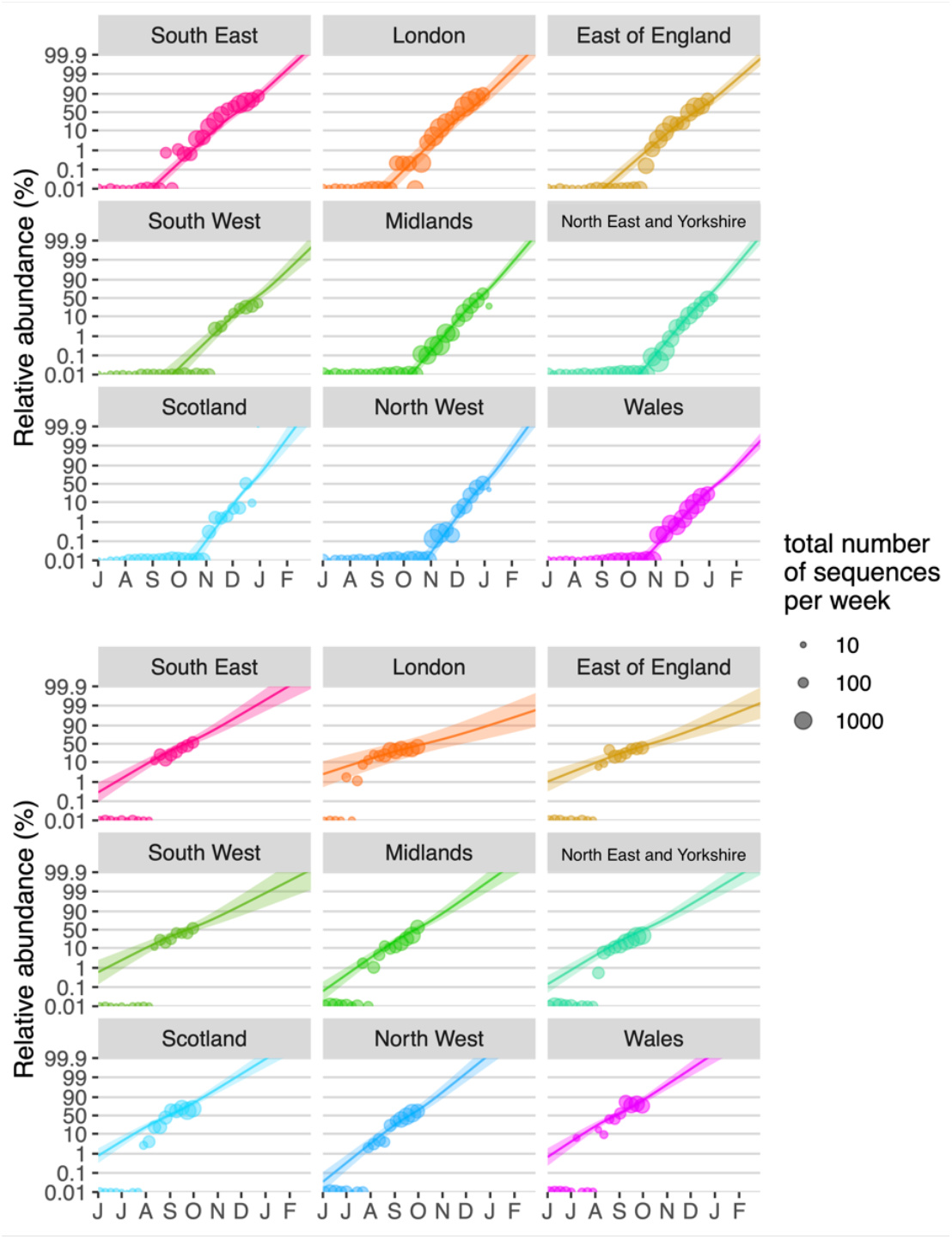
Binomial GLMMs fitted on the COG-UK sequence data with separate-slopes by region (models 2a and 2g in Table S1) show that VOC 202012/01 has been displacing all other SARS-CoV2 variants at a consistently high rate across different regions in the UK (A, Table S1), with pairwise Tukey posthoc tests for differences in slopes across regions mainly demonstrating a slightly slower rate of displacement in the East of England. By contrast, variant B.1.177, which in the UK became the major strain at the end of September, had a much lower competitive advantage in comparison with the minority variants that it displaced, evident from the much lower slope on a log(odds) scale (Table S1). In addition, pairwise Tukey posthoc tests for differences in slopes across regions demonstrate marked and significant cross-regional variation in the rate of spread of this variant (10 out of 36 pairwise comparisons with *P* < 0.05). This supports the idea that VOC 202012/01 enjoys a consistent competitive advantage, whilst the small competitive advantage enjoyed by variant B.1.177 may have been largely, though perhaps not exclusively, the result of stochastic introduction events, e.g. linked with travel to Spain (*53*), where it was first observed.

**Fig. S6.**
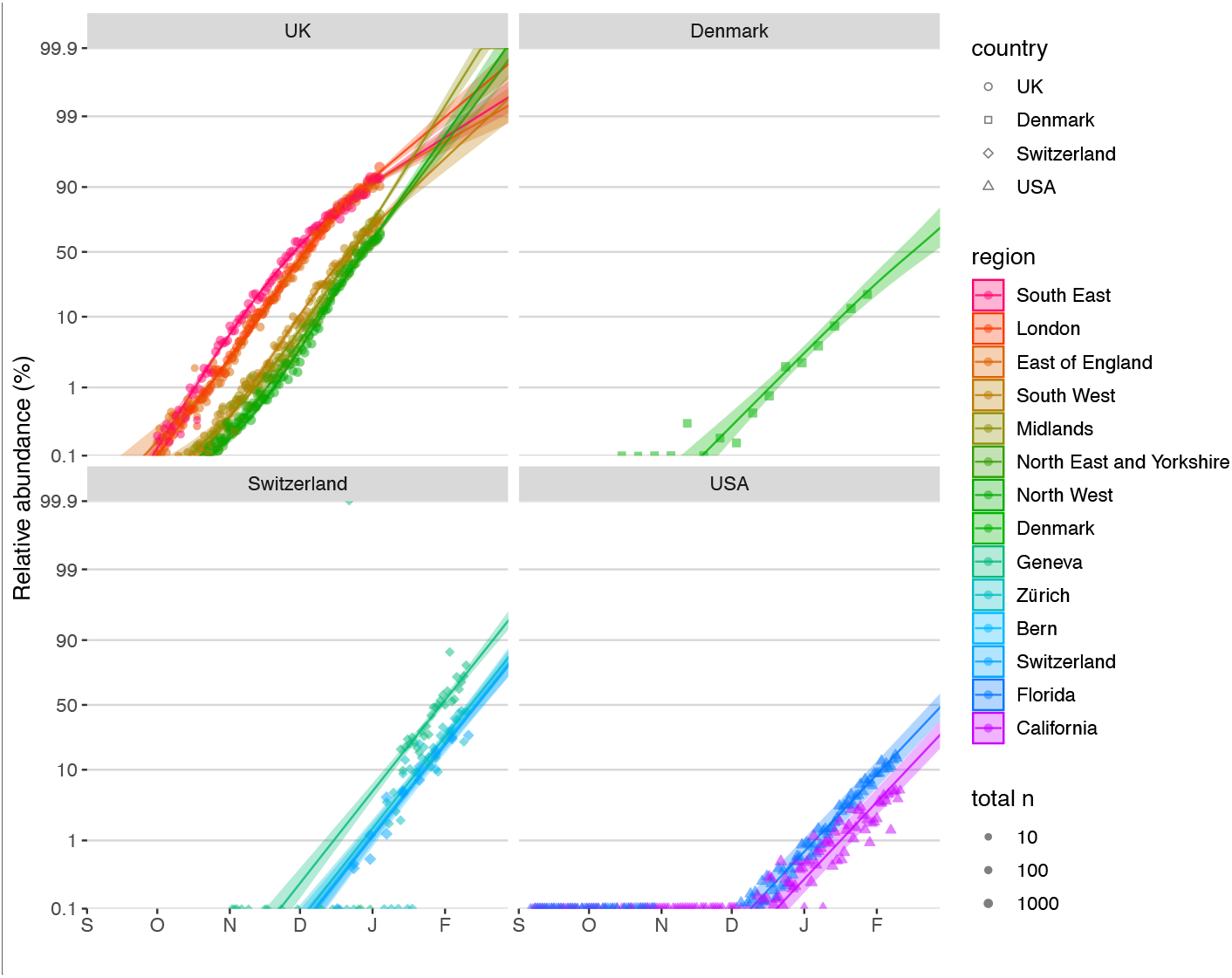
Estimates of the rate at which VOC 202012/01 is displacing other variants in the UK, Denmark, Switzerland and the USA based on S-gene target failure data (UK & USA) and sequencing of SARS-CoV2 strains (Denmark and Switzerland) or RT-PCR 501Y.V1 re-screening (Switzerland) (*32*–*36*) (binomial GLMMs, with adjustment for the true positive rate in the analyses of SGTF data). For the UK data, a binomial spline GLMM with NHS region and a natural cubic spline with 3 degrees of freedom in function of sampling date plus the interaction between both as fixed effects and an observation-level random effect to take into account overdispersion provided the best fit based on the BIC criterion. The different intercepts for the different regions reflect differences in the dates of introduction of the VOC, which in terms of relative timing match those inferred from the COG-UK data (Fig. S3 and S5). By contrast, for the data from Denmark, Switzerland and the USA, models with sample date included as an additive fixed effect and region (or state) coded as either a fixed factor (for Switzerland) or as a random intercept (for Denmark and the USA), and an observation-level random effect included to take into account overdispersion (*54*), provided the best fit. This indicates a near-constant rate of displacement in the different regions or states within each of those countries. Based on these data, the overall average growth advantage in the UK, Denmark, Switzerland and the USA are estimated at 10.9% (10.7-11.1%), 8.0% (6.7-9.2%), 10.1% (9.2-10.9%) and 8.4% (8.0-8.8%) per day, which with a generation time of 5.5 days would translate to mean transmission advantages of 83% (81-84%), 55% (45-66%), 74% (66-82%) and 59% (56-63%) (cf. models 2h, 3a, 3b and 3c in Table S1). For Switzerland and the USA, slightly earlier introduction in Geneva and Florida are apparent.

**Fig. S7.**
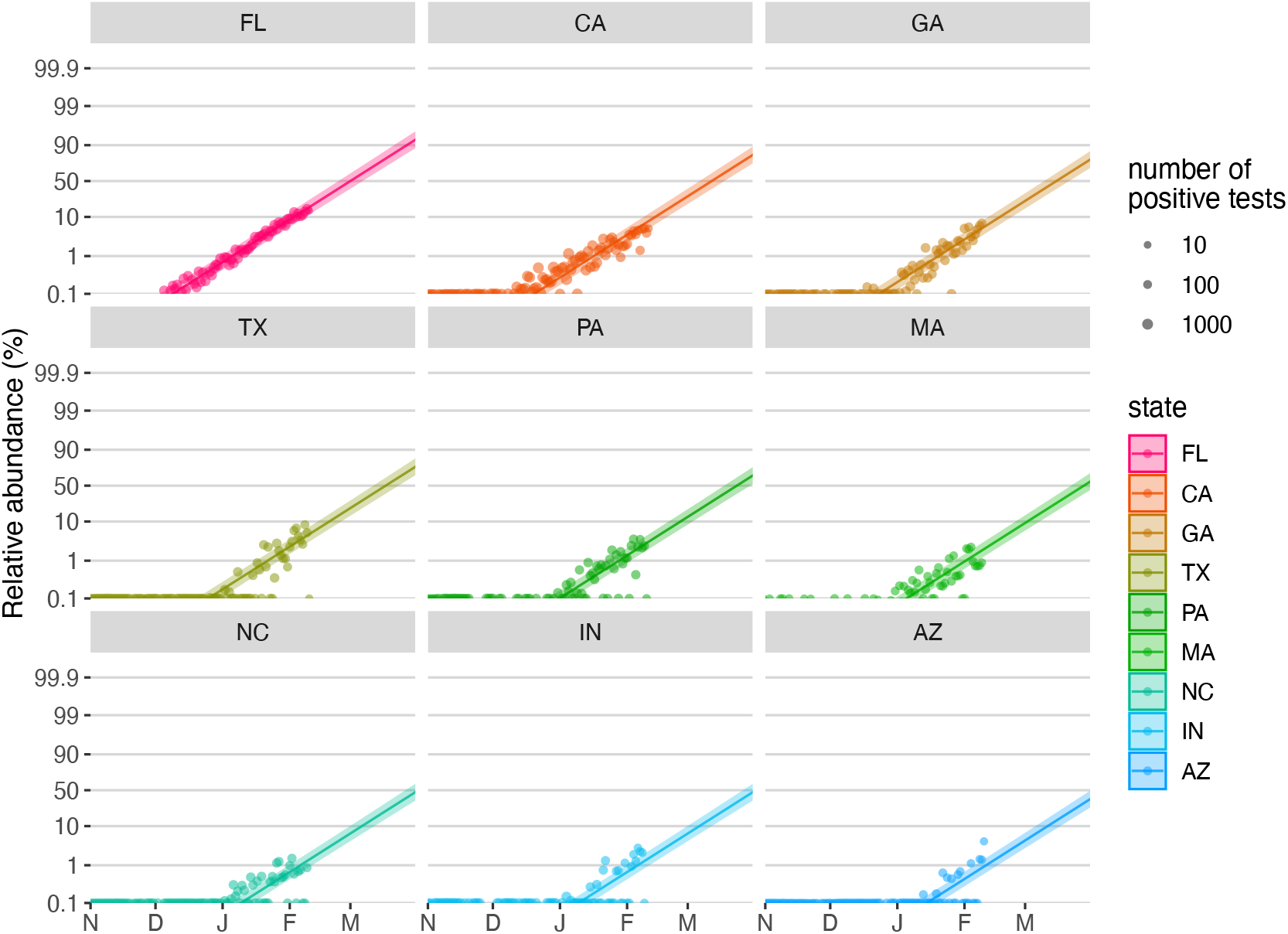
Estimates of the rate at which VOC 202012/01 is displacing other variants in the USA shown by state (displaying the 9 states with the most data), based on S-gene target failure data (*36*). A binomial GLMM with sample date included as an additive fixed effect, state coded as a random intercept and an observation-level random effect included to take into account overdispersion (*54*), provided the best fit based on the BIC criterion. In this model, binomial counts were adjusted to take into account the true positive rate (the proportion of the S-negative samples that were indeed the VOC), which was estimated using an independent binomial GLMM fitted on sequencing data of S-negative samples, whereby sample date was included as a continuous covariate and state was coded as a random intercept. A random-slope model, whereby the regression slope could vary by state, was also fitted but provided a worse model fit based on the BIC criterion. This indicates that with the current evidence, the VOC is displacing the other strains at a comparable rate across the different states of the US. The different intercepts reflect different dates of introduction.

**Fig. S8.**
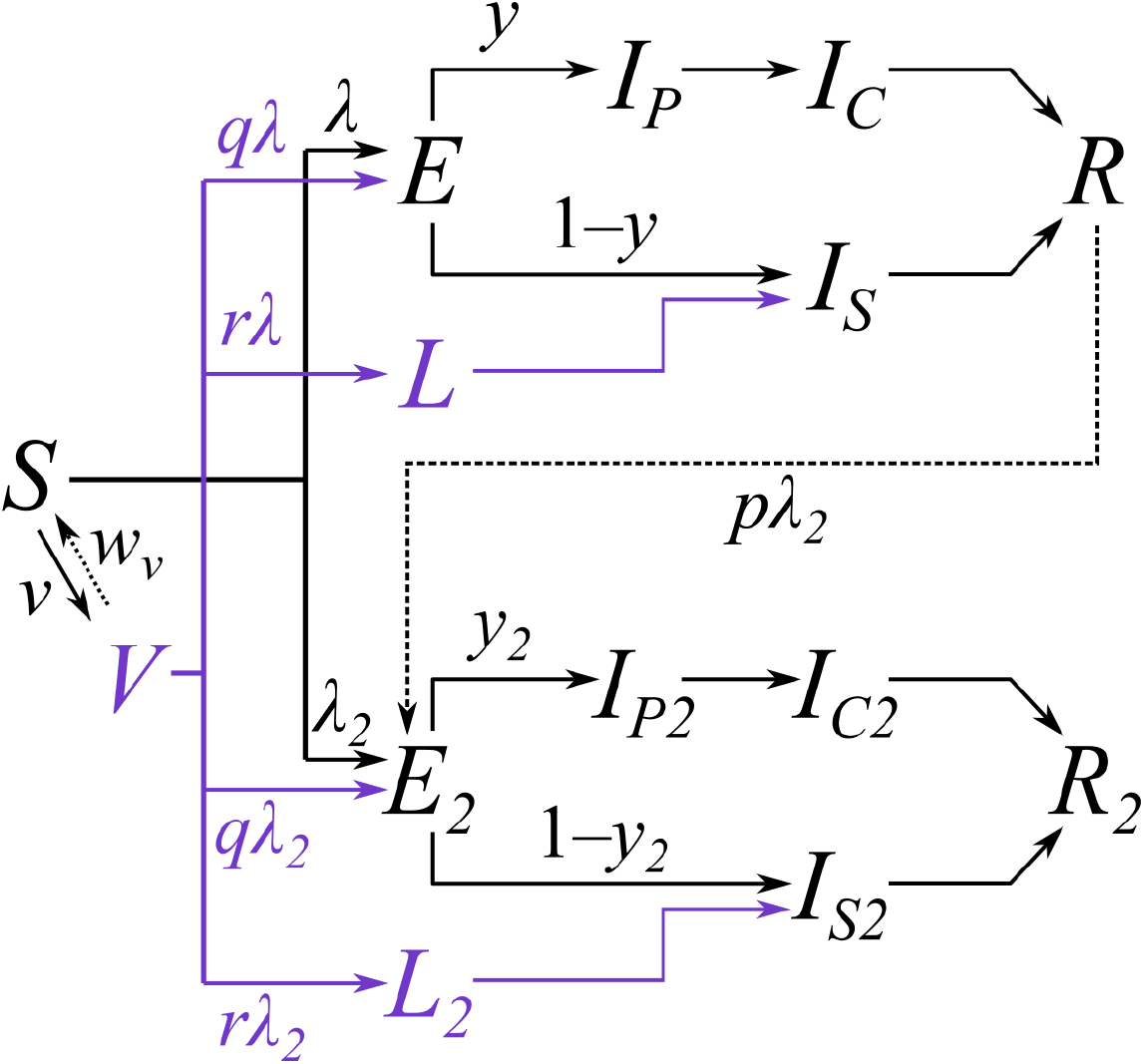
Diagram of the two-strain model with vaccination. Subscripts for age group and region are omitted from this diagram and only certain key parameters are shown. Compartments and processes in purple apply to the vaccine model only. S, susceptible; E, exposed; L, latent (see below); I_P_, preclinically infectious; I_C_, clinically infectious; I_S_, subclinically infectious; R, recovered; V, vaccinated. Subscript 2 represents compartments and parameters for VOC 202012/01. Above, *λ* and *λ*_2_ are the force of infection for preexisting variants versus VOC 202012/01; *y* and *y*_2_ are the fraction of cases that develop clinical symptoms for preexisting variants versus VOC 202012/01; *v* is the rate of vaccination; *w*_*v*_ is the waning rate of vaccination (assumed to be zero for this manuscript); *p* captures cross-protection against VOC 202012/01 conferred by immunity to preexisting variants; *q* captures vaccine protection against disease; and *r* captures vaccine protection against infection. *L* and *L*_2_ are additional compartments for a latent period prior to subclinical infection only (i.e. with zero probability of clinical infection). For a vaccine with efficacy against disease *e*_*d*_ (e.g. *e*_*d*_ = 0.95 for this manuscript) and efficacy against infection *e*_*i*_ (e.g. *e*_*i*_ = 0.6 for this manuscript), we assume *r* = (1 – *e*_*i*_) *e*_*d*_ and *q* = (1 – *e*_*i*_) (1 – *e*_*d*_).

**Fig. S9.**
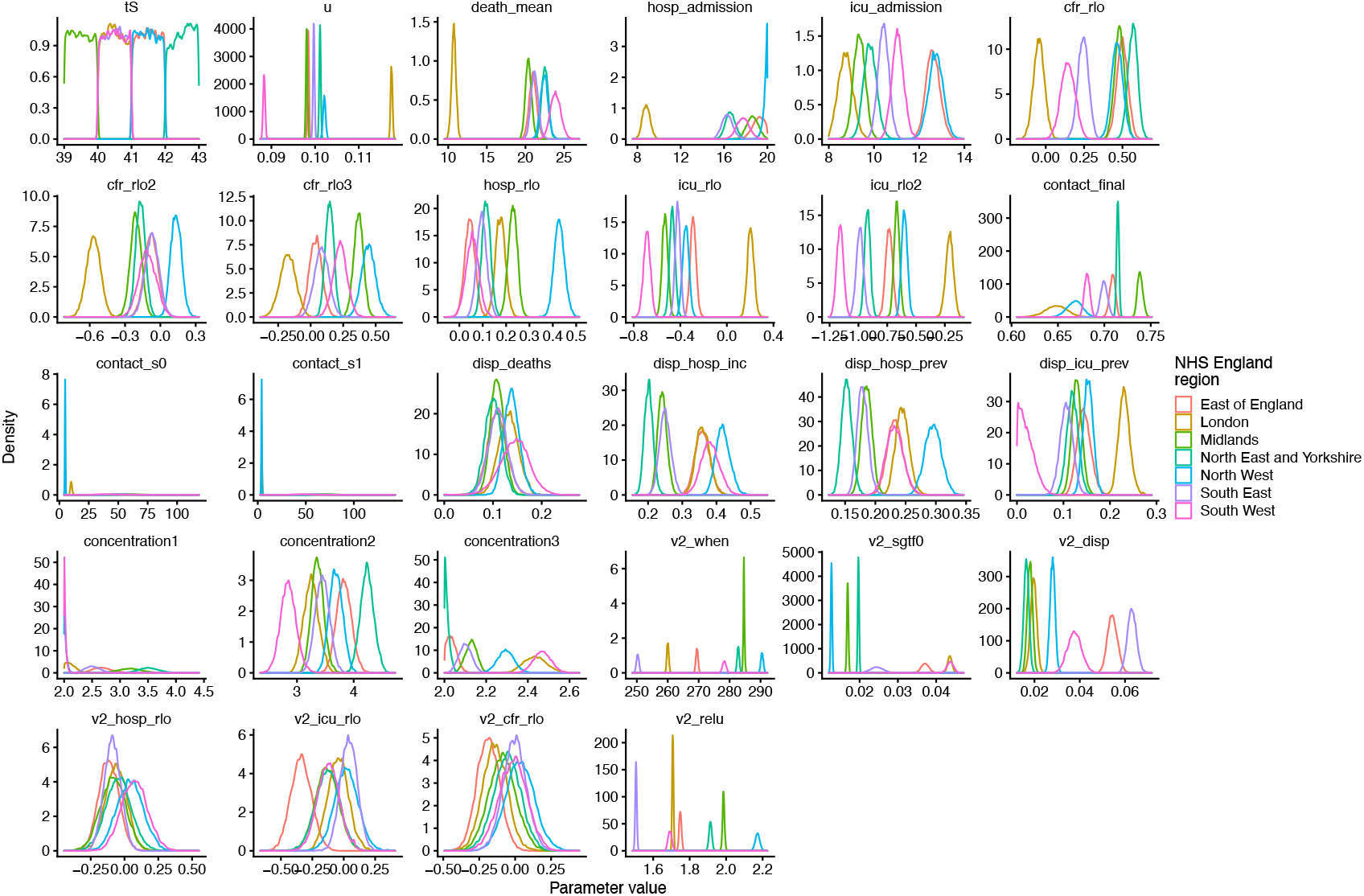
Model posterior densities for the “increased transmissibility” model for seven NHS England regions. See Table S2 for parameter definitions.

**Fig. S10.**
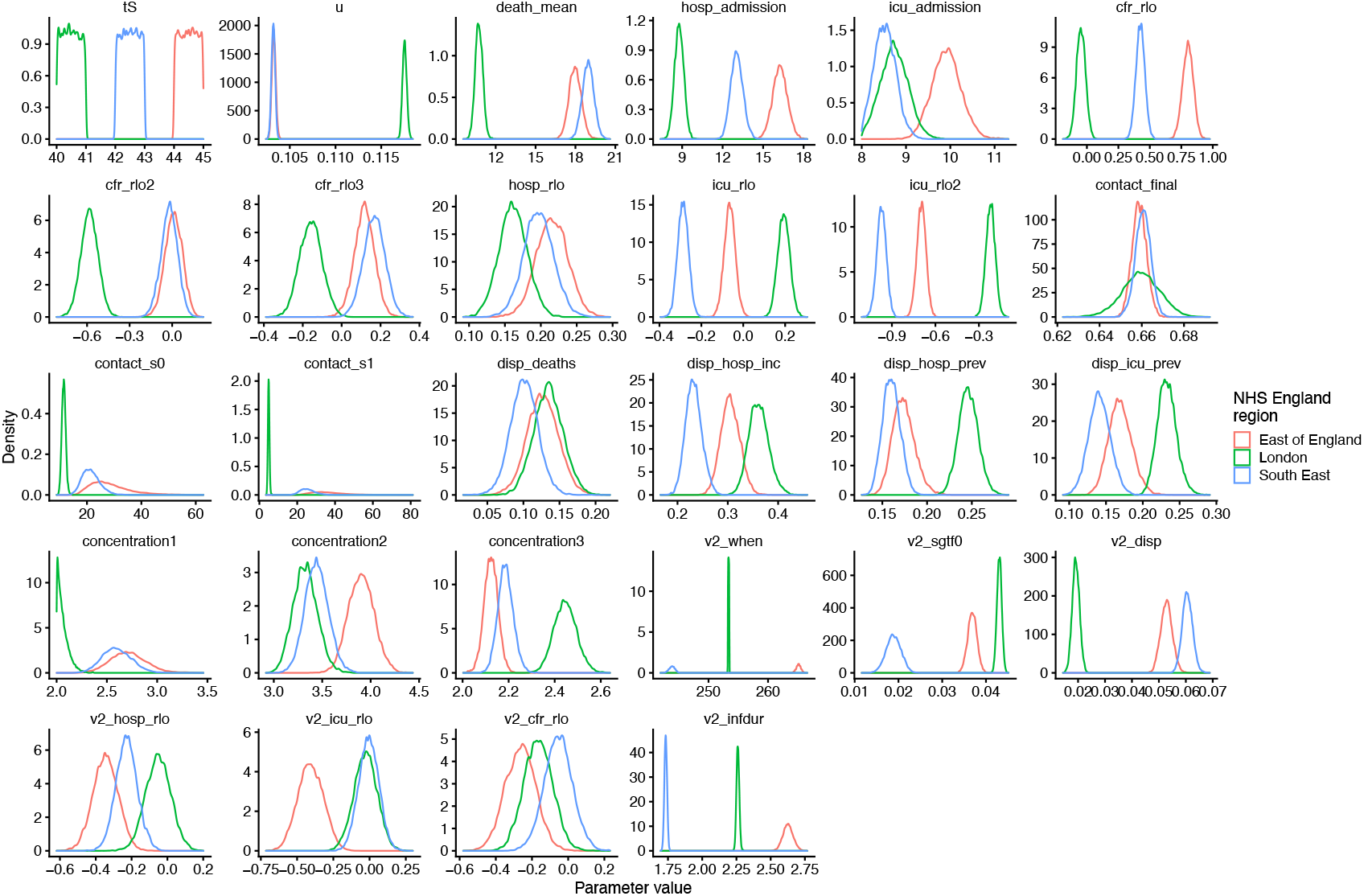
Model posterior densities for “longer infectious period” for three NHS England regions. See Table S2 for parameter definitions.

**Fig. S11.**
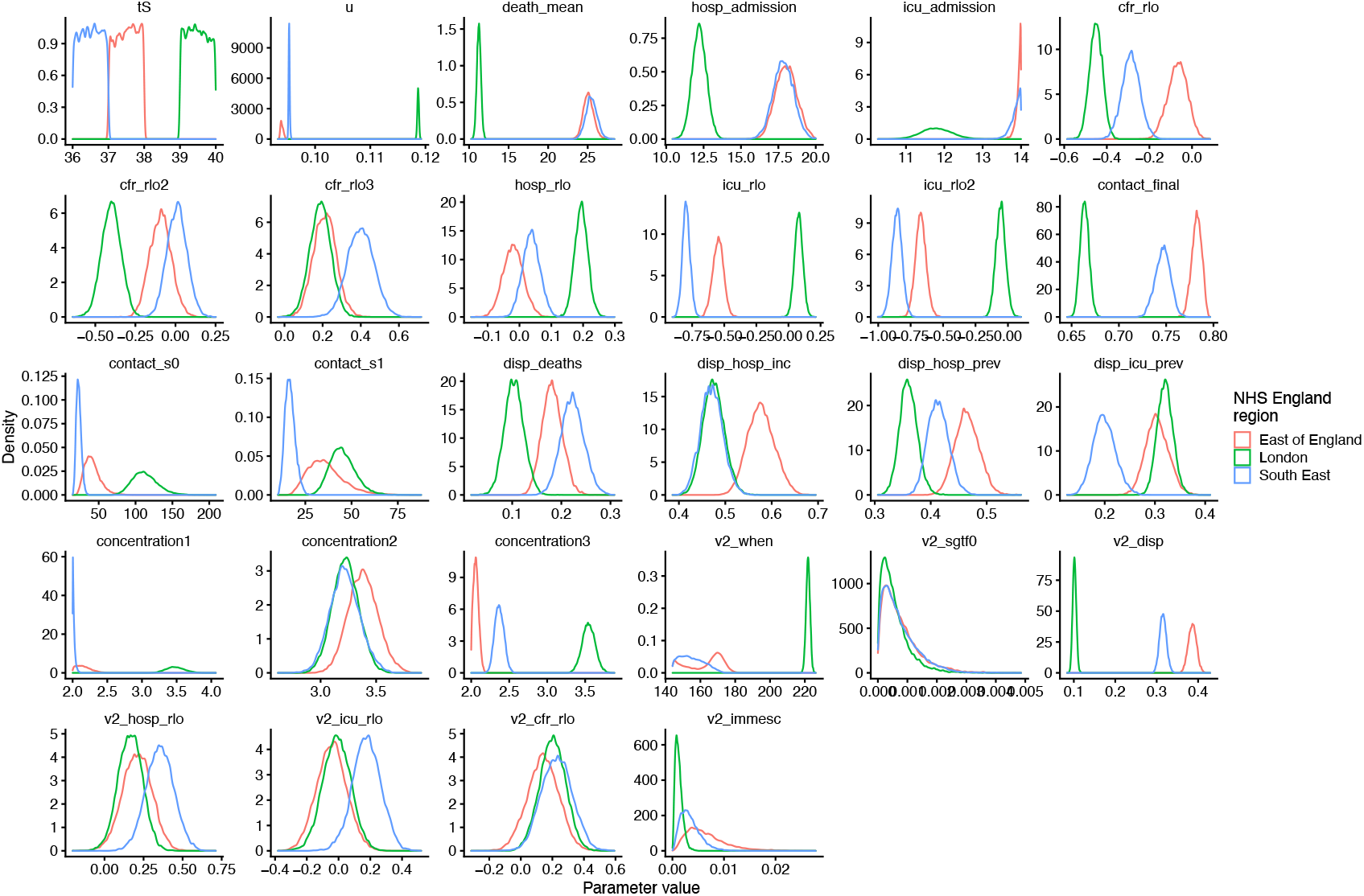
Model posterior densities for “immune escape” model for three NHS England regions. See Table S2 for parameter definitions.

**Fig. S12.**
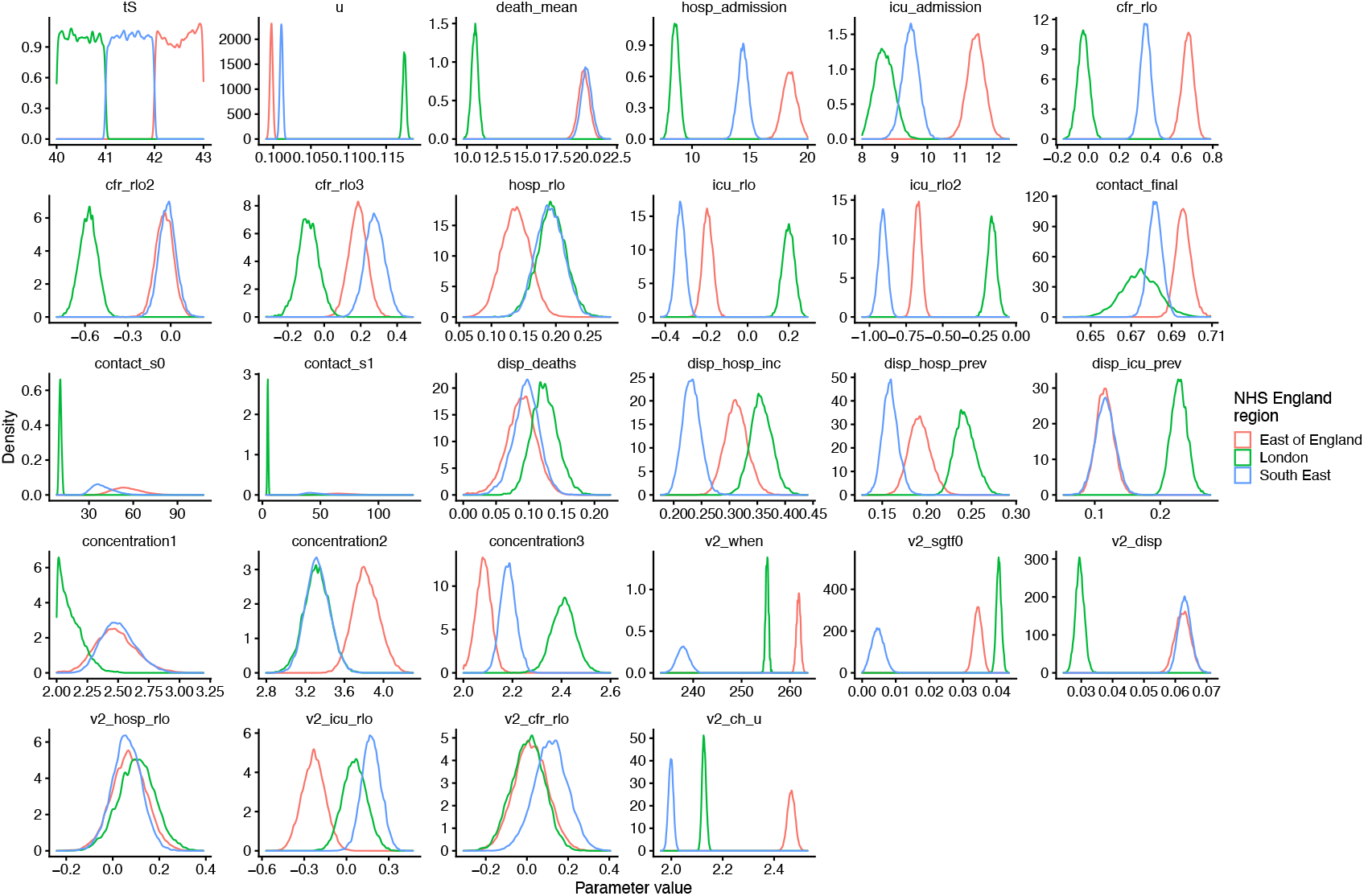
Model posterior densities for “increased susceptibility in children” model for three NHS England regions. See Table S2 for parameter definitions.

**Fig. S13.**
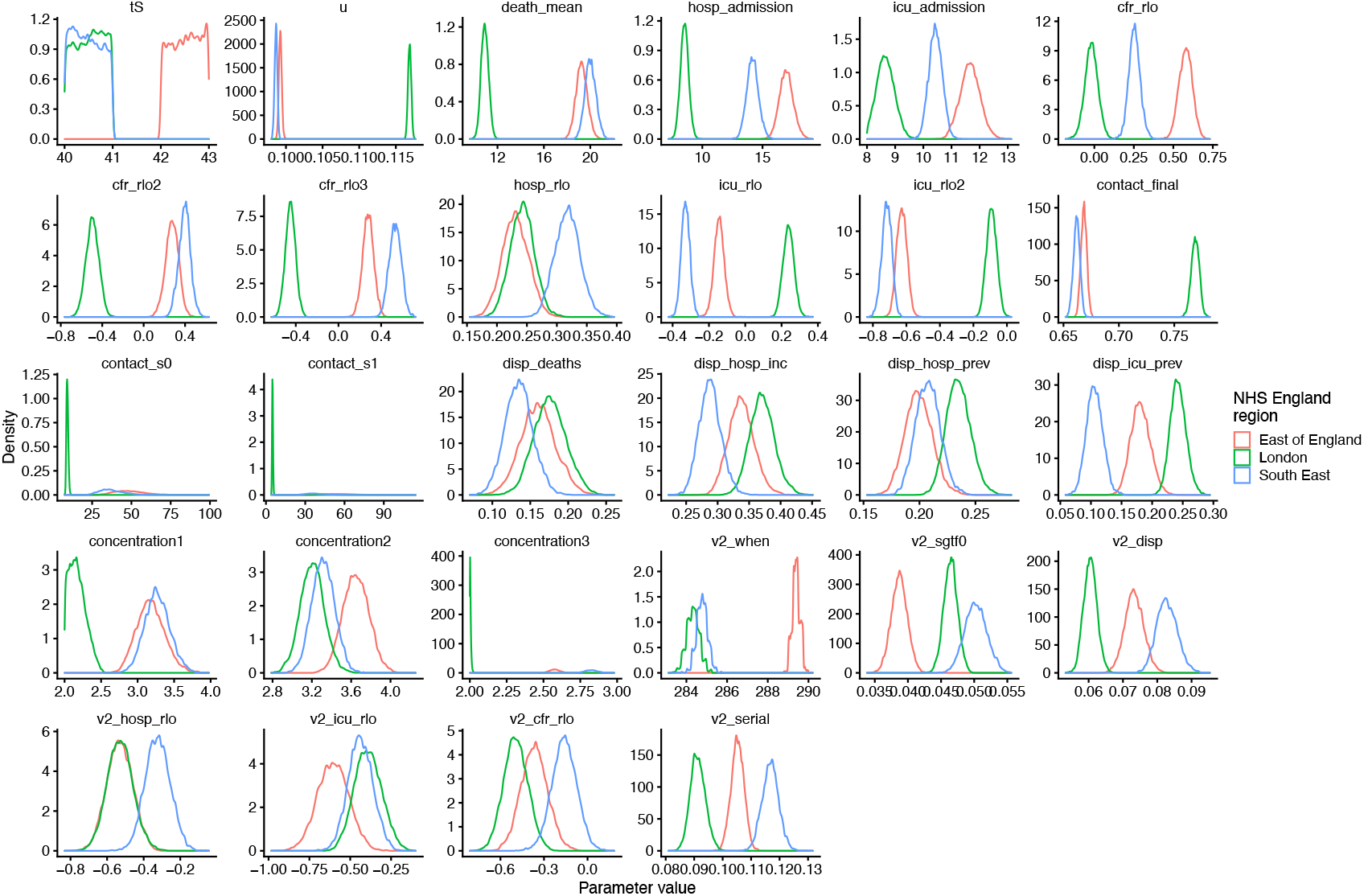
Model posterior densities for “shorter generation time” model for three NHS England regions. See Table S2 for parameter definitions.

**Fig. S14.**
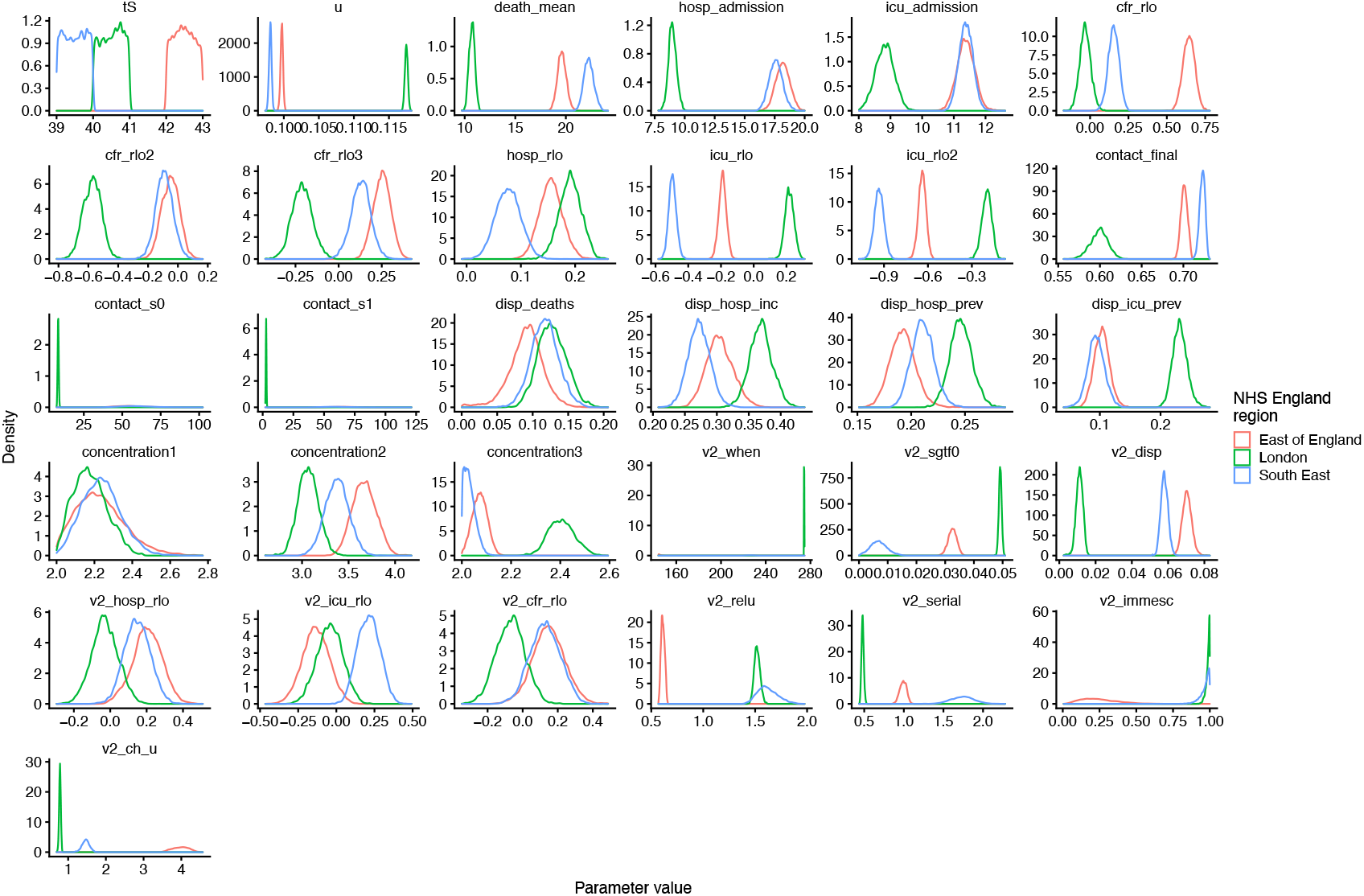
Model posterior densities for a “combined” model with increased transmissibility, altered serial interval, immune escape, and altered susceptibility in children. See Table S2 for parameter definitions.

**Fig. S15.**
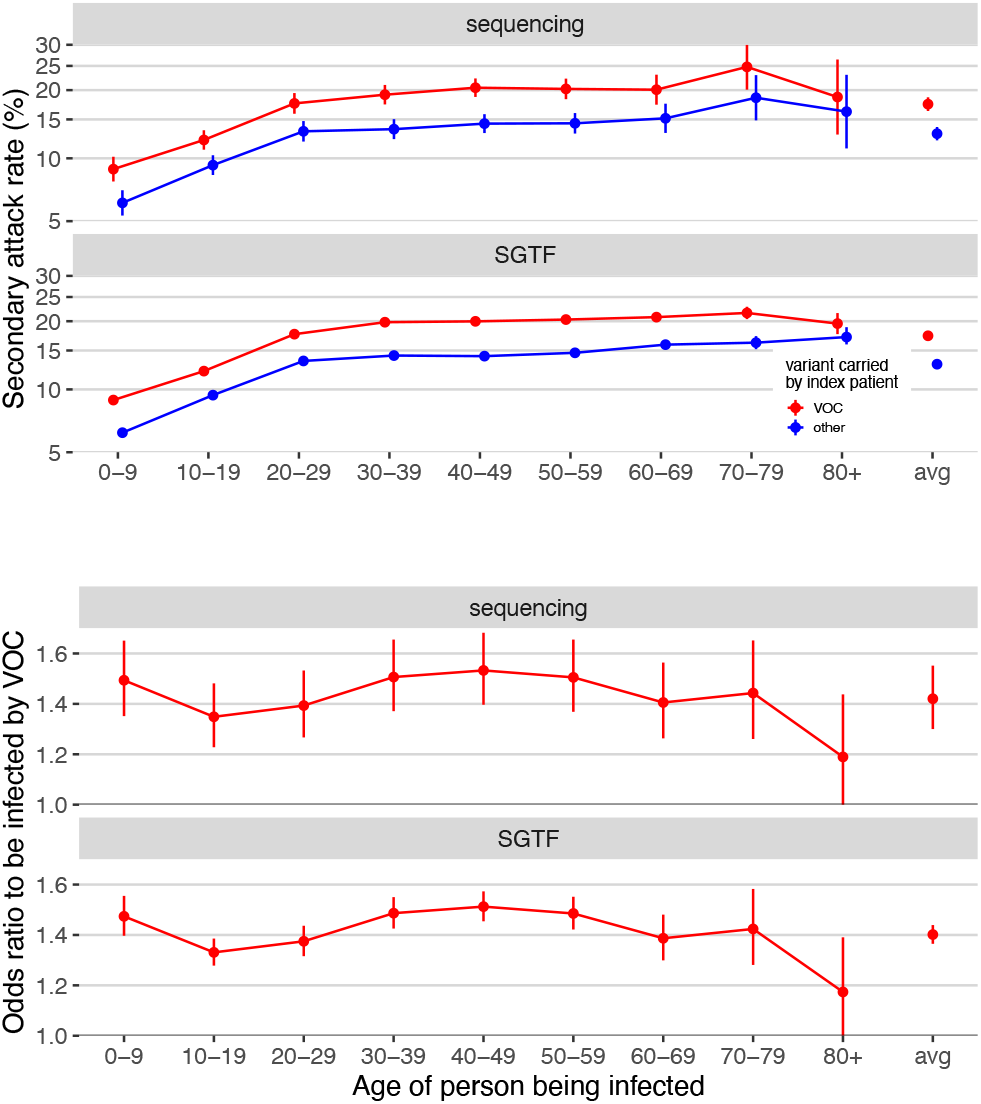
Analysis of age-stratified secondary attack rates, based on data reported by Public Health England (*63*) (data derived from the COG-UK dataset, the PHE Second Generation Surveillance System and NHS Test and Trace). A binomial GLM with data type (sequence data or S-gene target failure), age group of the person being infected, and variant (VOC 202012/01 or not) plus all first order interaction effects shows that the odds to be infected by an index patient carrying the VOC is consistently higher than by those carrying other variants (A). Sidak posthoc tests show the odds to be infected by the VOC to be significantly greater than by a non-VOC variant for nearly all age groups (for all age groups and both data types 2-sided P < 1E-7, except for 80+ where P = 0.07 and 0.06 for sequencing and SGTF data, respectively). The mean probability for secondary contacts to become infected in function of age was not significantly different across both types of data (no significant data type by age interaction effect, Type III test, 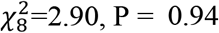) and there was also no difference in the estimated increased odds to be infected by a VOC vs. a non-VOC index patient (no significant data type by variant interaction effect, Type III test, 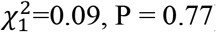). The mean odds ratio to be infected by an index patient carrying the VOC vs. a non-VOC variant across all age groups and both data types was 1.41 [1.34, 1.48] 95% CIs. The relative susceptibility to be infected by the VOC showed little variation in function of the age of the person being infected, with only the 40-49 category being slightly more susceptible to be infected by a VOC vs a non-VOC carrying index patient than average (measured in terms of difference in log odds ratios, Sidak age group x variant interaction contrasts, z ratio = 3.45, P = 0.01, all other P > 0.05).

**Fig. S16.**
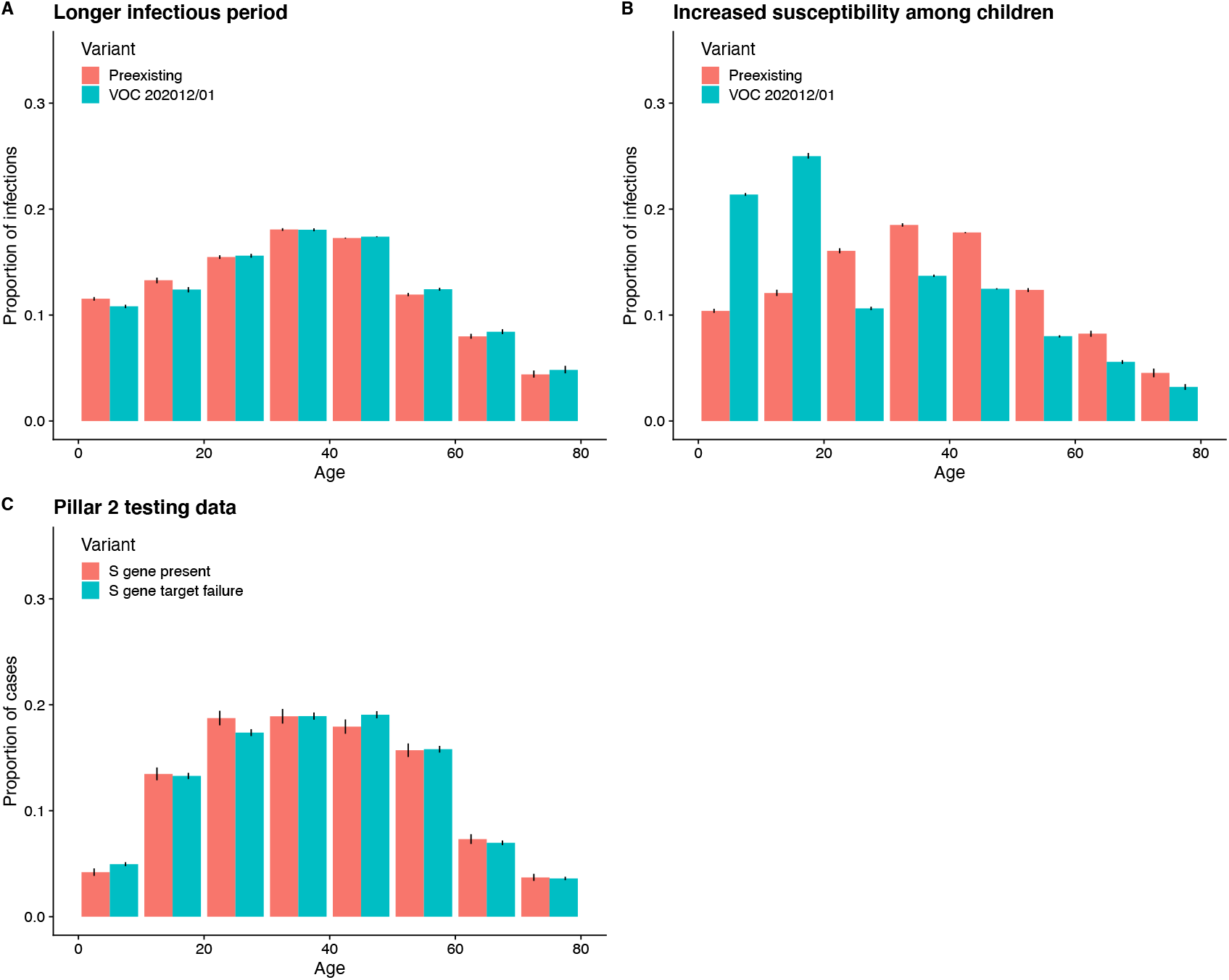
Comparison of age distribution of infections in the two-strain model, depending upon biological mechanism of VOC 202012/01 growth rate, and age distribution of Pillar 2 (community) SARS-CoV-2 tests with and without S gene target failure. Contrasting (A) “longer infectious period” and (B) “increased susceptibility among children” models with (C) pillar 2 testing data. Measured in the fitted model and empirically for the South East NHS England region in December 2020. Empirical data (C) does not show any marked increase of S gene target failure comparable to the increase in VOC 202012/01 infections in the “increased susceptibility among children” model (B). The lower overall proportion of Pillar 2 tests among 0–9-year-olds in the testing data may be due to lower propensity for testing in this age group.

**Fig. S17.**
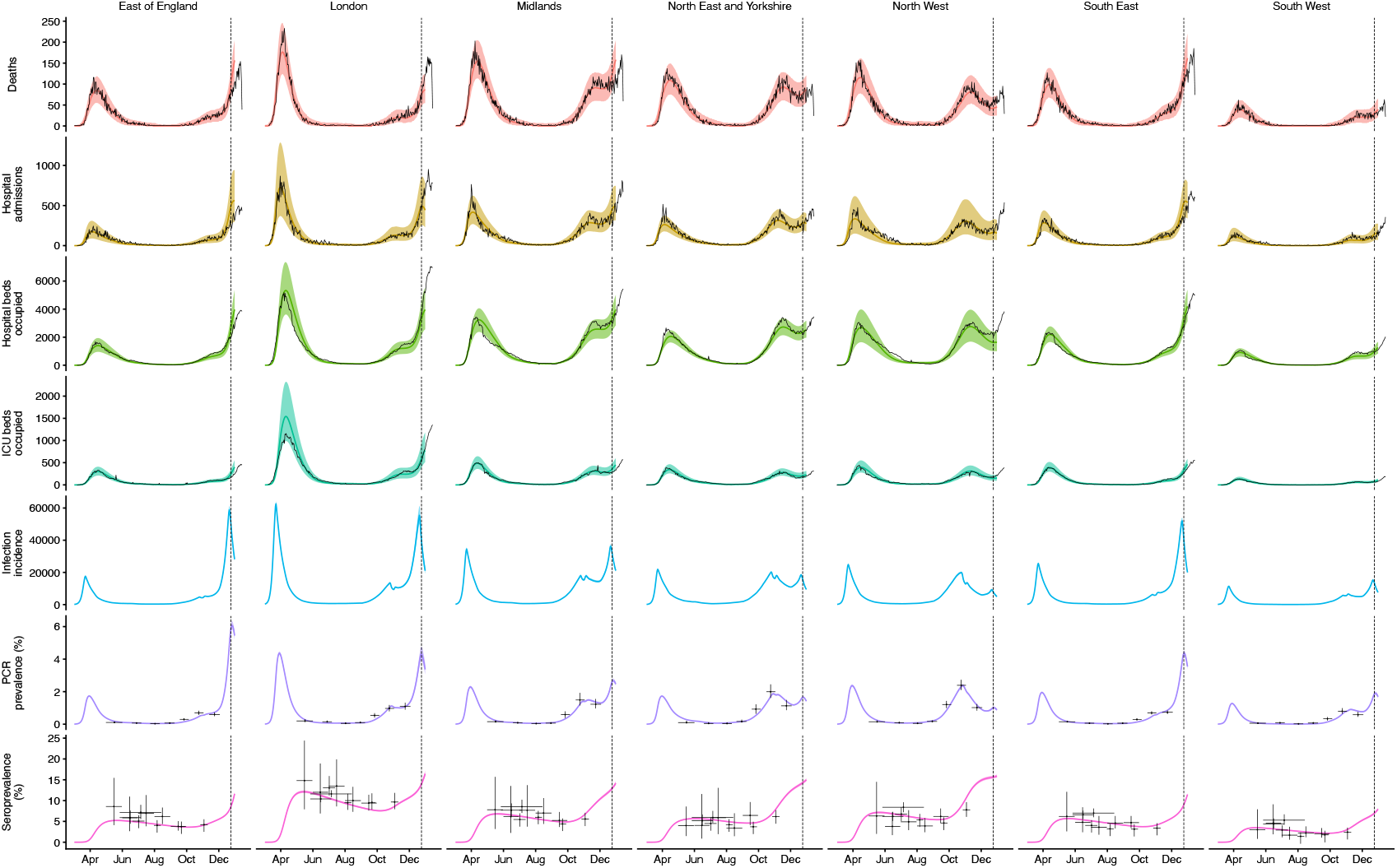
Fit of “increased transmissibility” model to data up to 24 December 2020. Black lines show observed data, while coloured lines and shaded regions show median and 95% credible intervals from the fitted model.

**Fig. S18.**
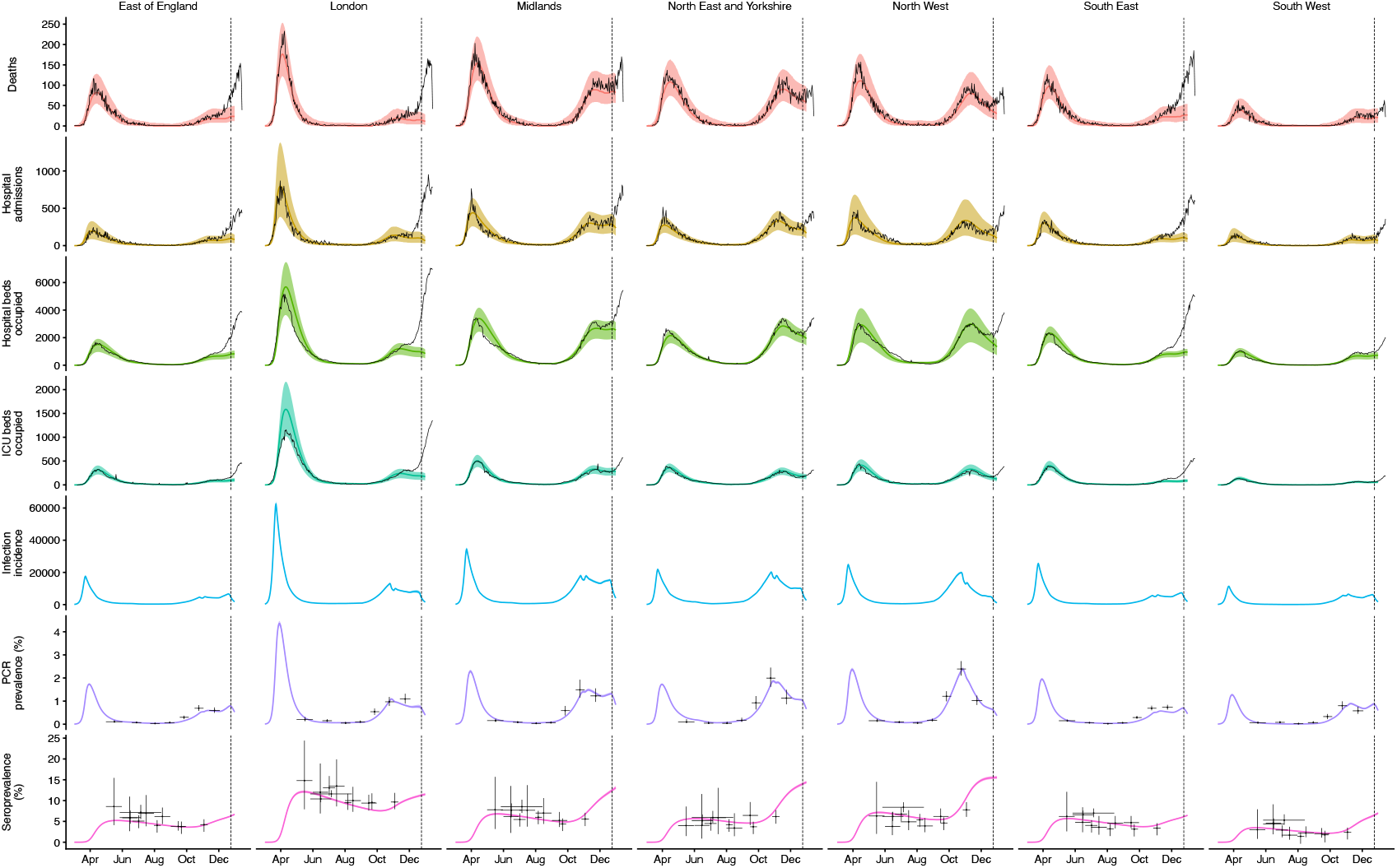
Fit of “increased transmissibility” model to data up to 24 December 2020, with the emergence of the second strain (VOC 202012/01) disabled. Surges in the East of England, London, and the South East are no longer captured. Black lines show observed data, while coloured lines and shaded regions show median and 95% credible intervals from the fitted model.

**Fig. S19.**
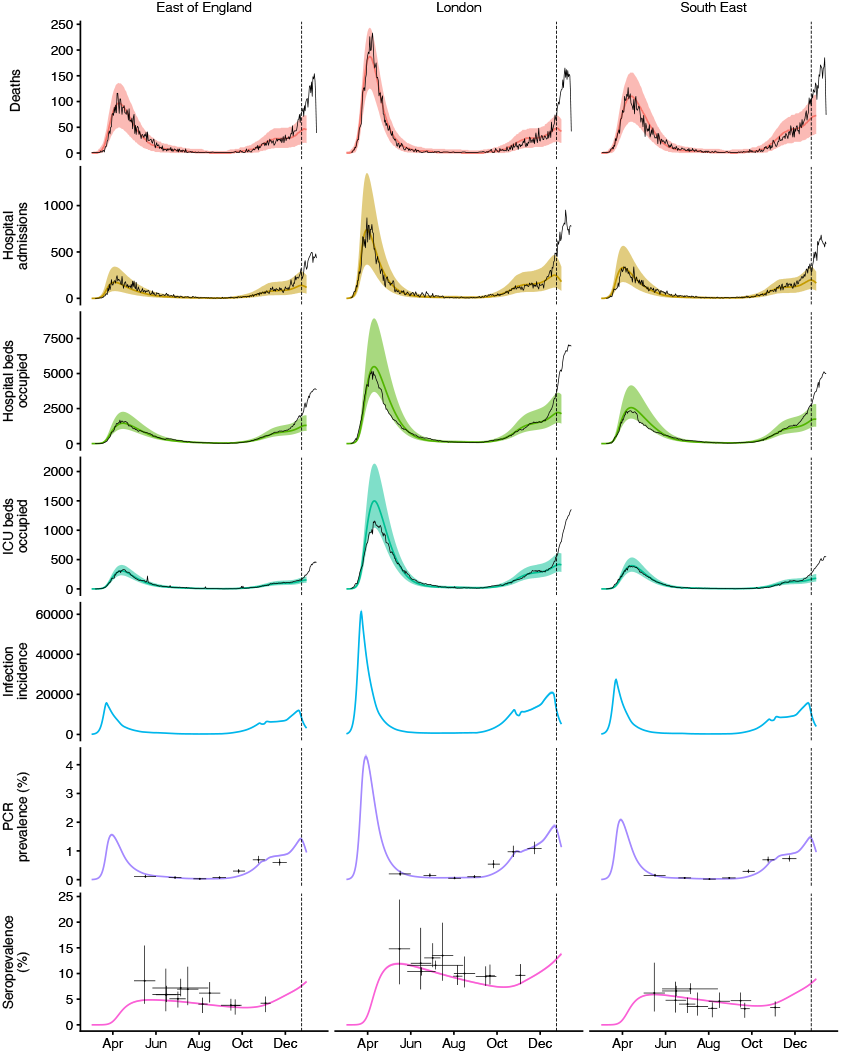
Fit of a model with no second strain up to 24 December 2020. The model increases transmission of the original strain to compensate, but cannot capture surges in the East of England, London, and the South East. Black lines show observed data, while coloured lines and shaded regions show median and 95% credible intervals from the fitted model.

**Fig. S20.**
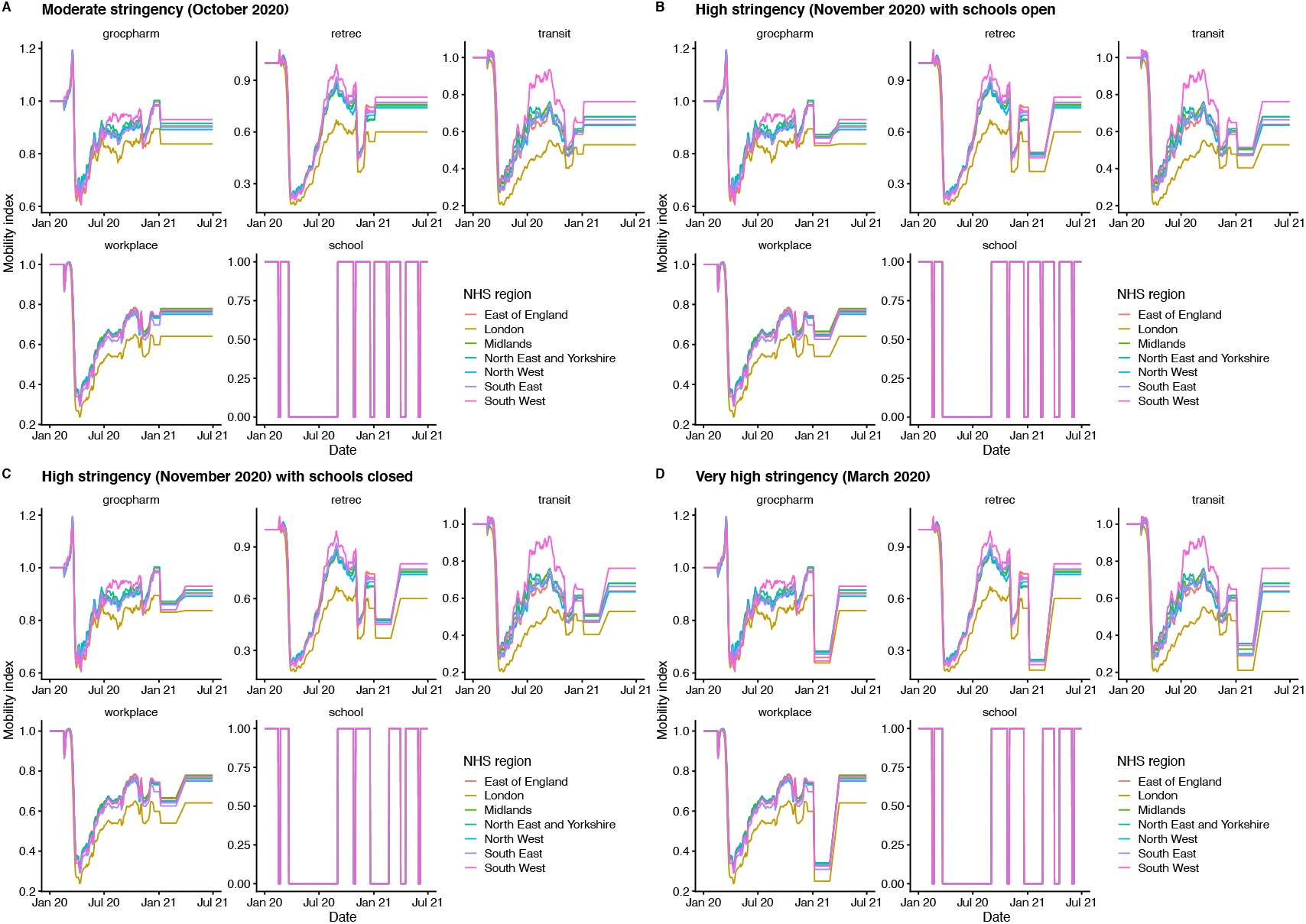
Google Mobility indices used for projections in the main text (Fig. 4).

**Fig. S21.**
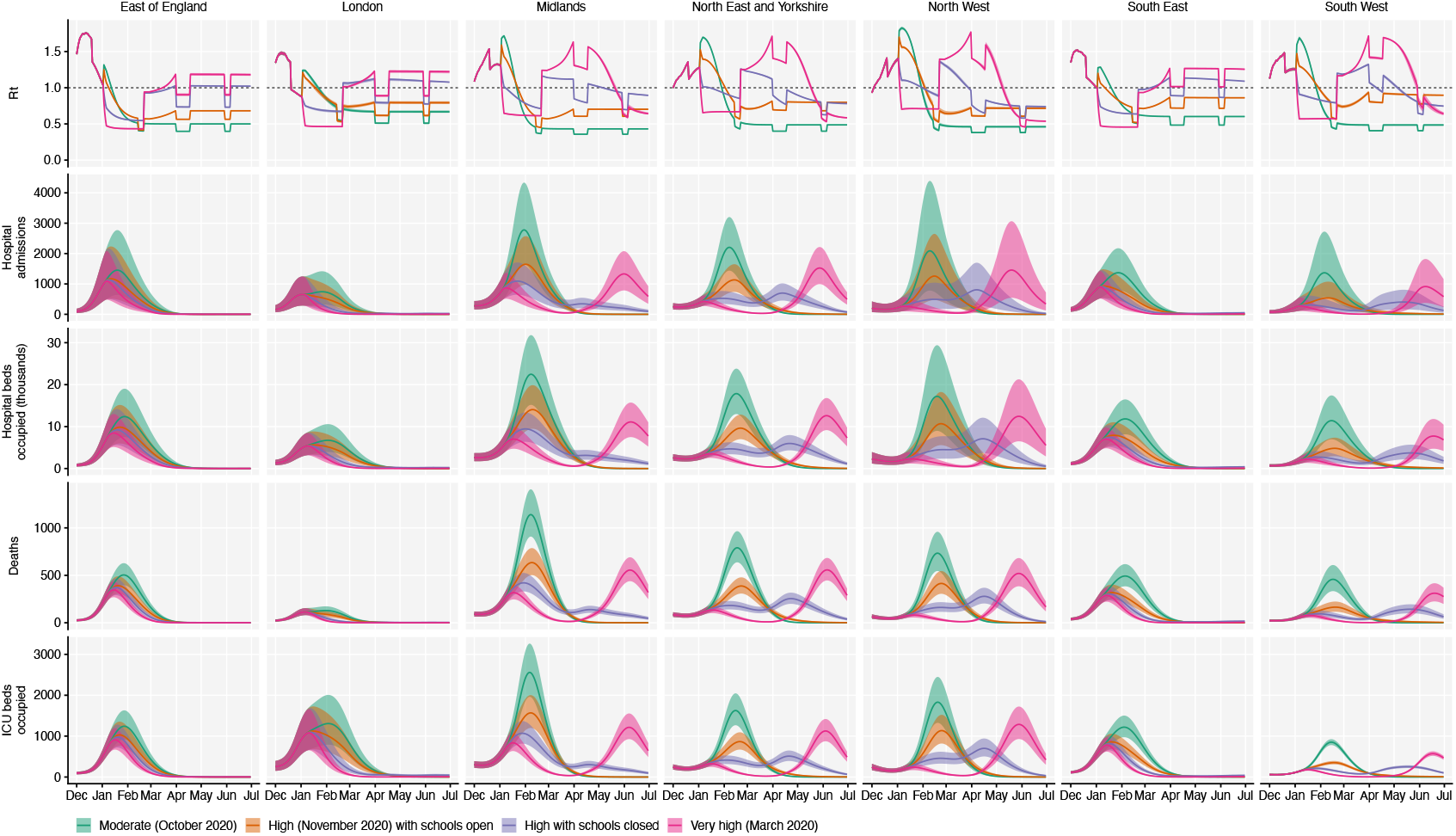
Model projections by NHS region, without vaccination. Median and 95% credible intervals are shown. See Fig. 4A, main text.

**Fig. S22.**
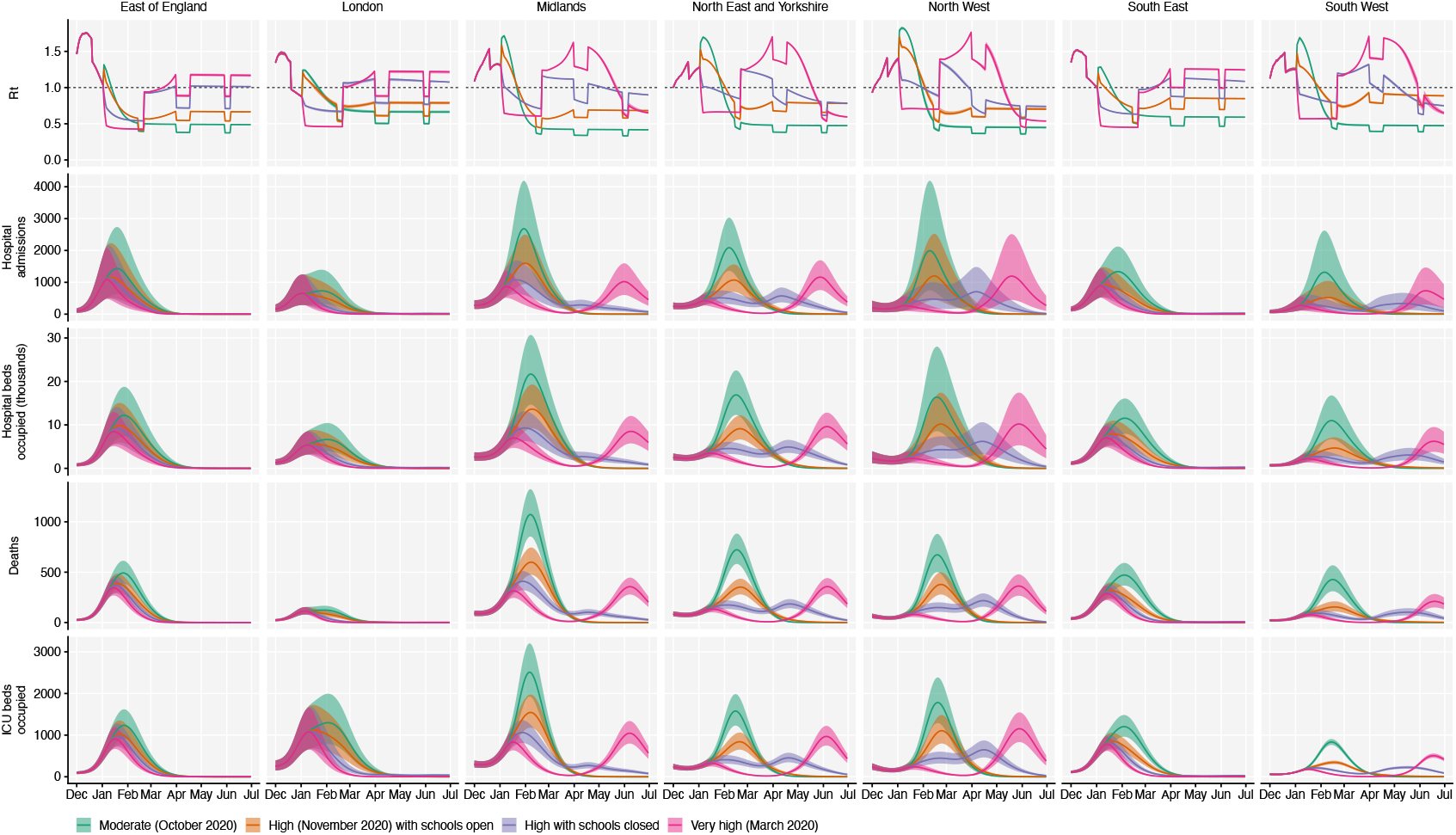
Model projections by NHS region, with 200,000 vaccinations per day. Median and 95% credible intervals are shown. See Fig. 4B, main text.

**Fig. S23.**
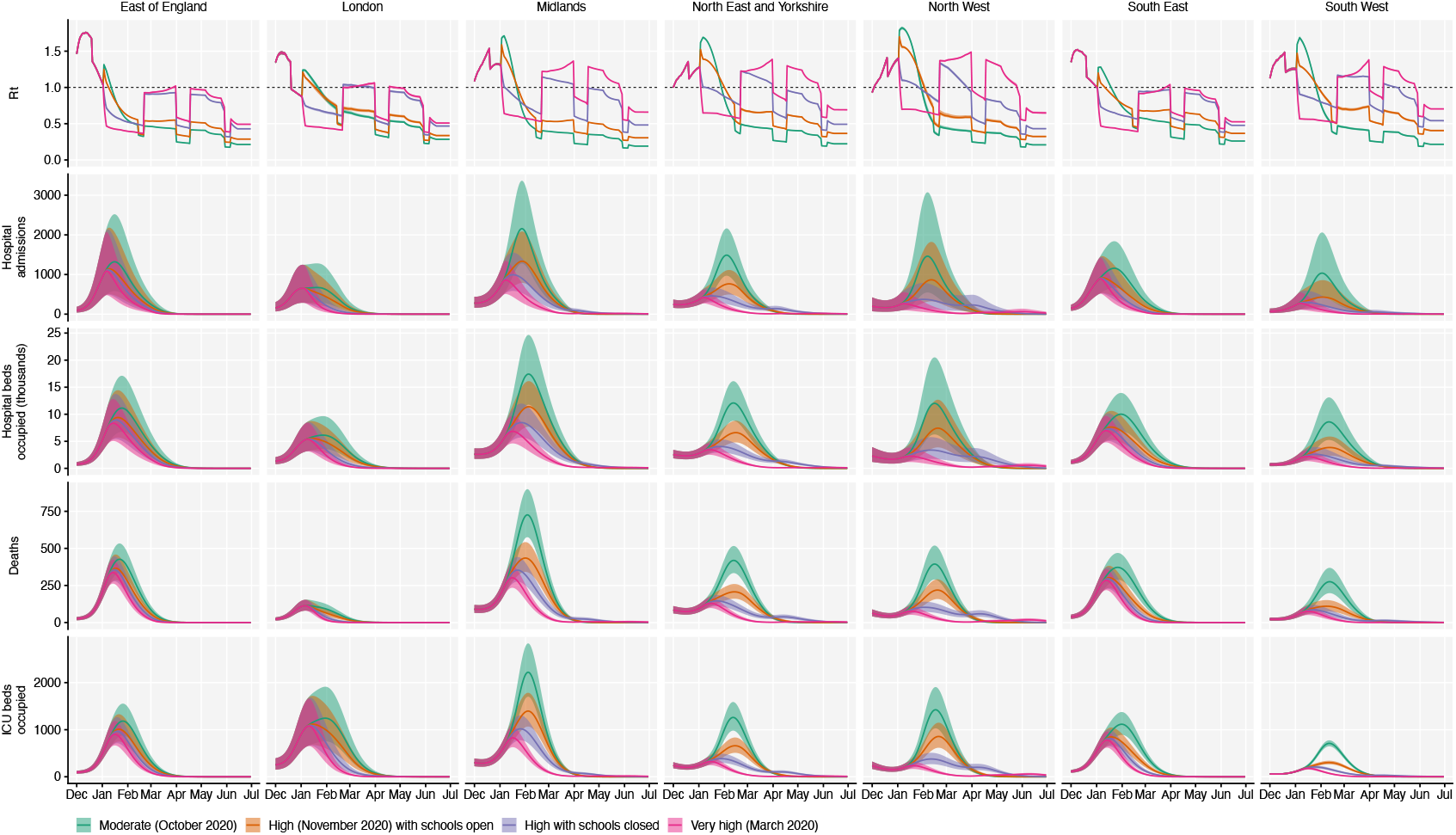
Model projections by NHS region, with 2 million vaccinations per day. Median and 95% credible intervals are shown. See Fig. 4C, main text.

**Fig. S24.**
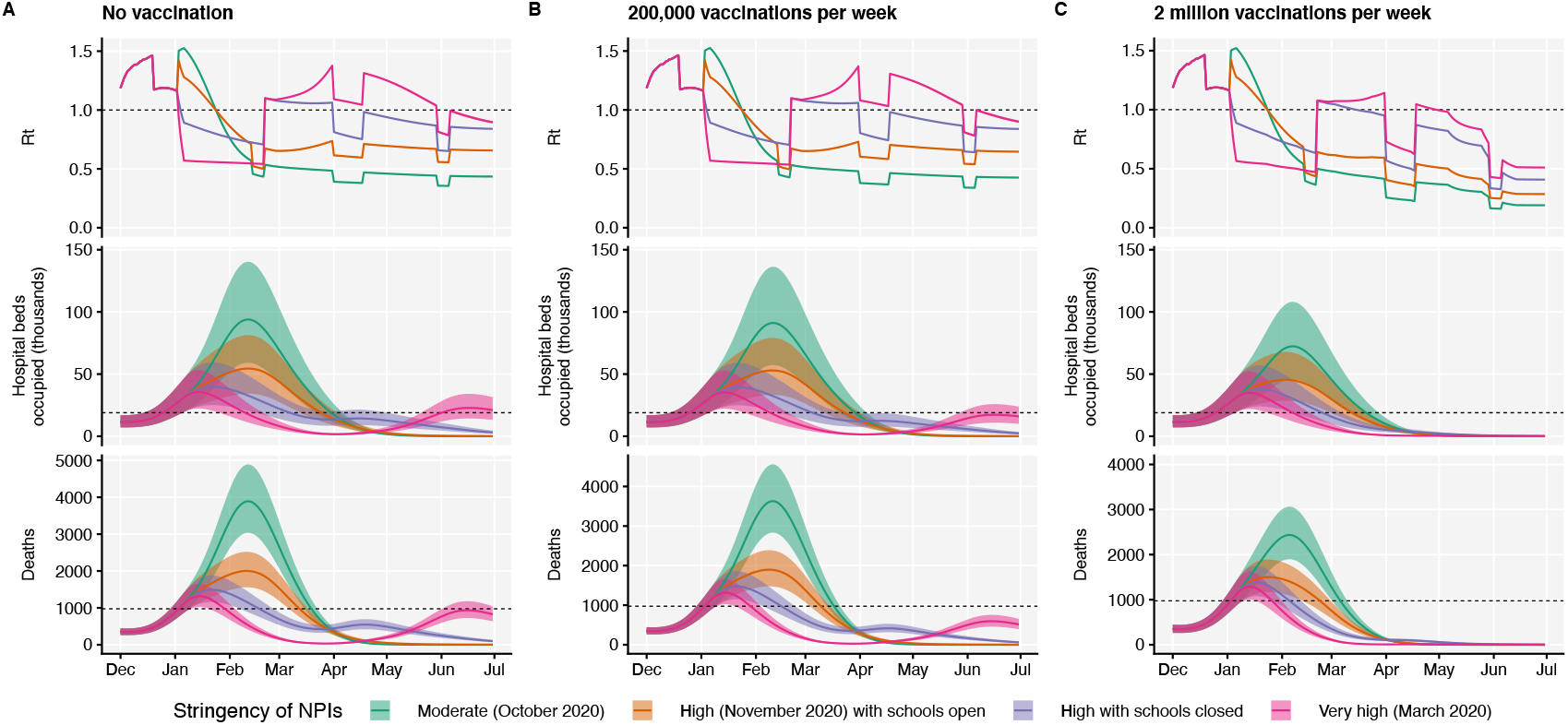
Model projections for England, with a seasonal component of transmission equivalent to 20% greater transmission at the peak of winter (1 January) relative to summer (1 July) (*22*). Median and 95% credible intervals are shown. See Fig. 4, main text.

## Supplementary Tables

**Table S1.**
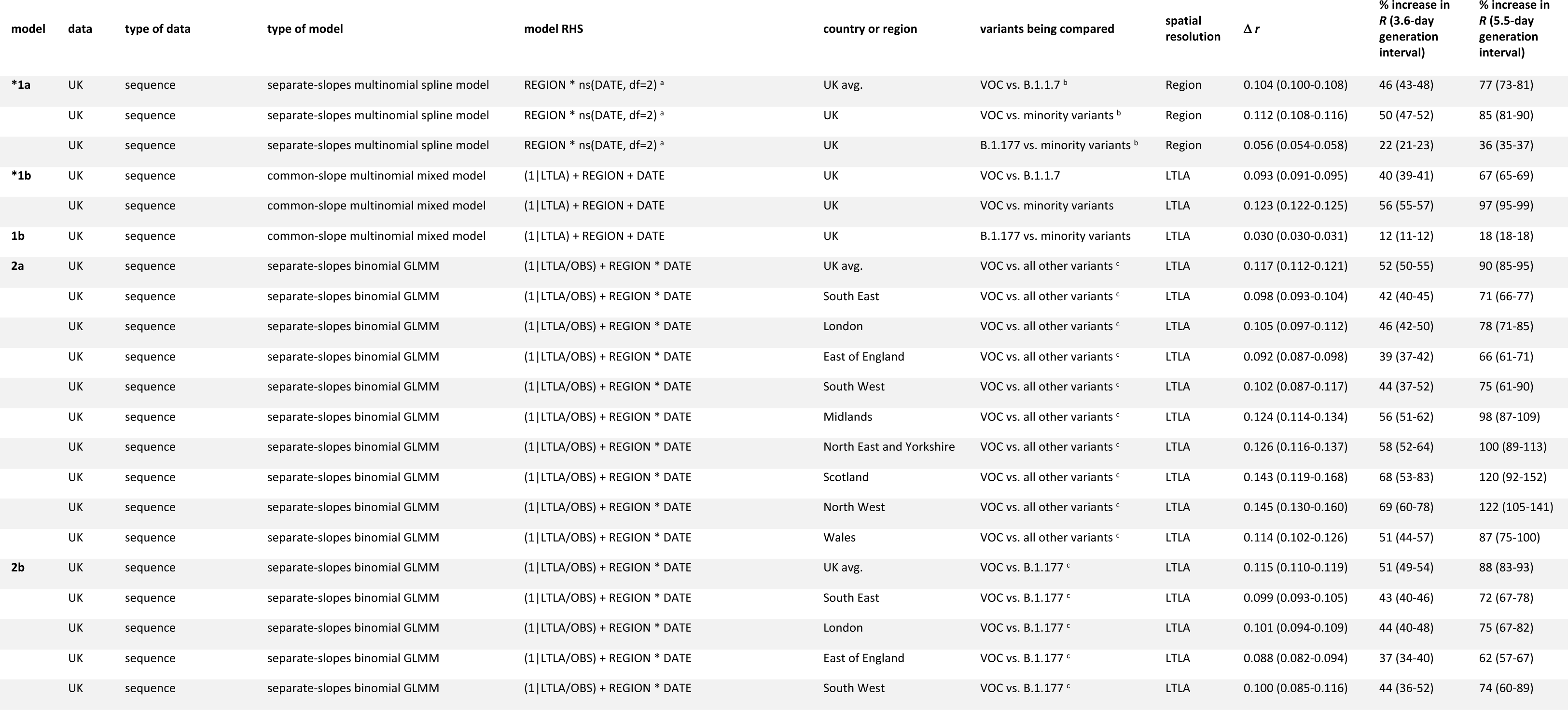
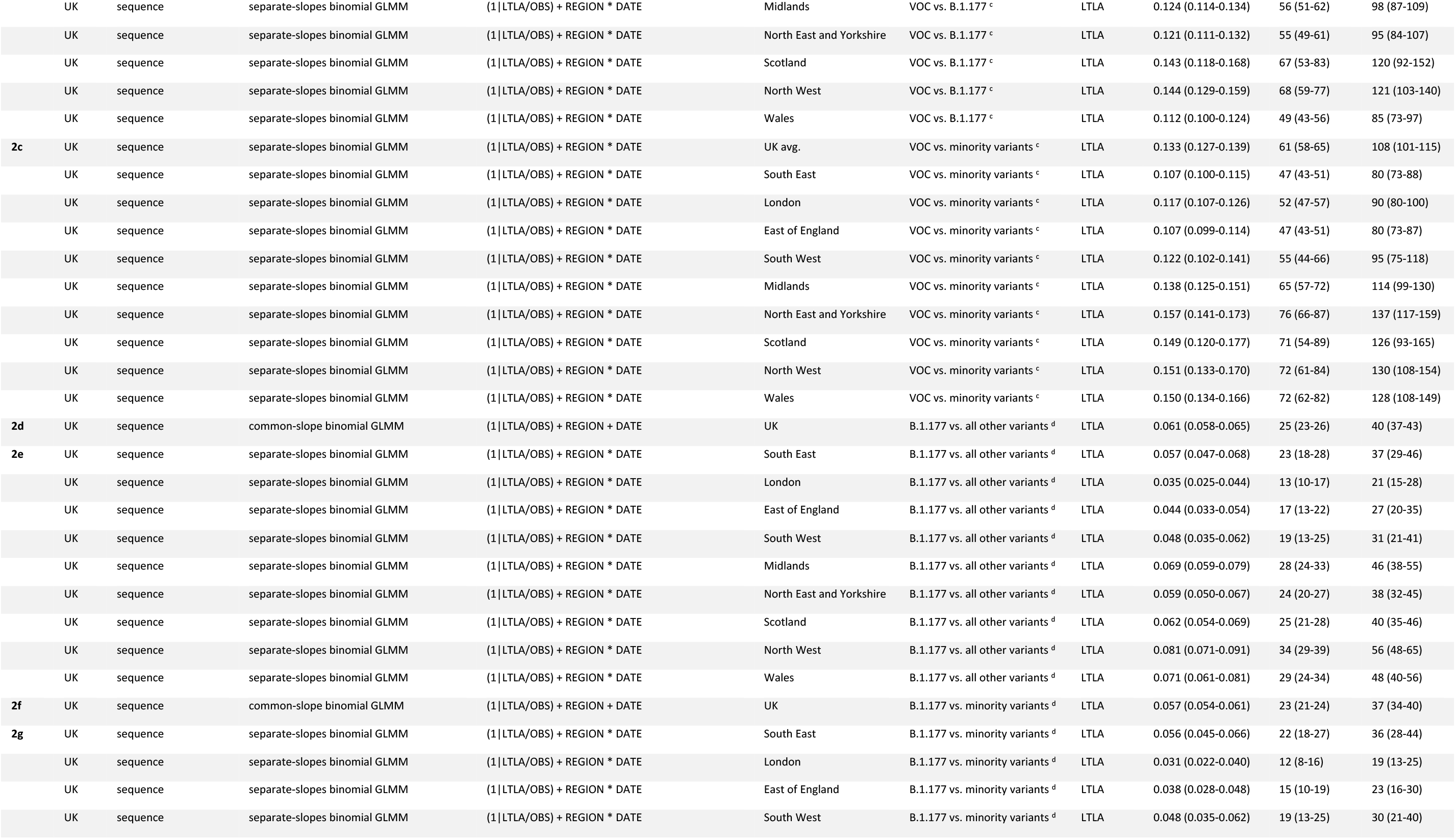
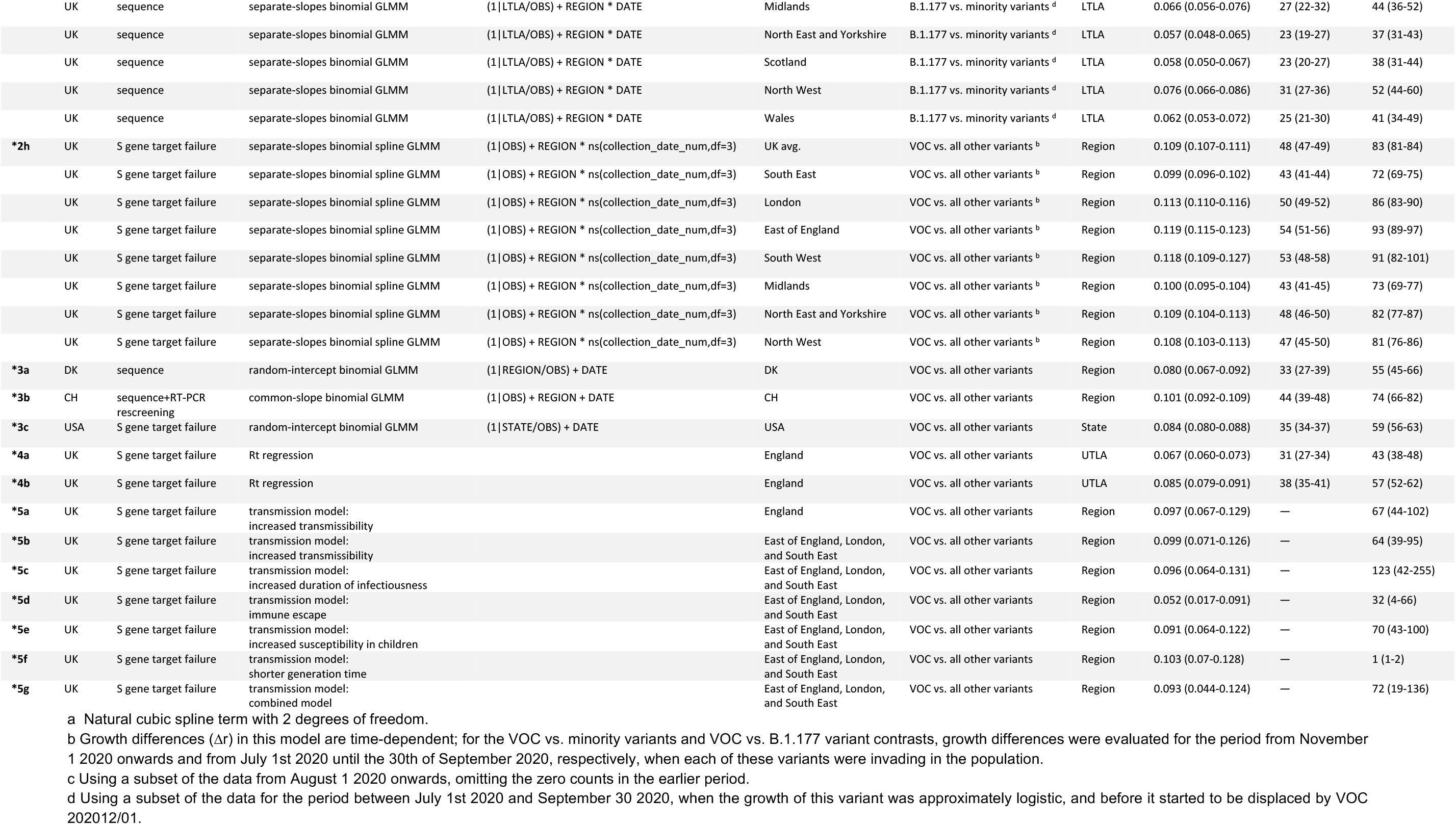
Estimates of increased growth rate. Models marked with an asterisk are those given in Table 1.

**Table S2.** Details of fitted parameters.

| Parameter | Description | Prior distribution | Notes |
| --- | --- | --- | --- |
| `tS` | Start date of epidemic in days after 1 January 2020 | $\sim \text{uniform}(0, 60)$ | Determines date at which seeding begins in region; starting on this date, one random individual per day contracts SARS-CoV-2 for 28 days |
| `u` | Basic susceptibility to infection | $\sim \text{normal}(0.09, 0.02)$ | Determines basic reproduction number $R_0$ |
| `death_mean` | Mean delay in days from start of infectious period to death | $\sim \text{normal}(15, 2)$ | Delay is assumed to follow a gamma distribution with shape parameter 2.2. Prior and shape of distribution informed by analysis of CO-CIN data (66). |
| `admission` | Mean delay in days from start of infectious period to hospital admission | $\sim \text{normal}(7.5, 1)$ | Delay is assumed to follow a gamma distribution with shape parameter 0.71. Prior and shape of distribution informed by analysis of CO-CIN data (66). |
| `icu_admission` | Mean delay in days from start of infectious period to ICU admission | $\sim \text{normal}(11.1, 1)$ | Delay is assumed to follow a gamma distribution with shape parameter 1.91. Prior and shape of distribution informed by analysis of CO-CIN data (66). |
| `hosp_rlo` | Log-odds of hospital admission, relative to age-specific probabilities of hospital admission given infection derived from Salje et al. (67). | $\sim \text{normal}(0, 0.1)$ | Based on Salje et al. (67), we assumed that the basic shape of the age-specific probability of hospitalisation given infection was $\text{logistic}(7.37 + 0.068a)$, where $a$ is the individual's age in years. This overall relationship is then adjusted according to the `hosp_rlo` parameter. |
| `icu_rlo,`<br>`icu_rlo2` | Log-odds of ICU admission, relative to age-specific probabilities of ICU admission given hospital admission derived from CO-CIN data. | $\sim \text{normal}(0, 0.1)$ | We fit a spline to CO-CIN data on hospital admission and ICU admission by age to derive the basic age-specific probability of ICU admission, which was then adjusted based on the `icu_rlo` and `icu_rlo2` parameters. `icu_rlo` applies for the first half of 2020 while `icu_rlo2` applies for the second half of 2020 into 2021. |
| `cfr_rlo,`<br>`cfr_rlo2,`<br>`cfr_rlo3` | Relative log-odds of fatality due to COVID-19 | $\sim \text{normal}(0, 0.1)$ | Based on Levin et al. (68), we assumed the basic shape of the age-specific infection fatality ratio of SARS-CoV-2 was $\text{logistic}(-7.56 + 0.121a)$ (see entry for `hosp_rlo`). This is adjusted by `cfr_rlo`, `cfr_rlo2`, and `cfr_rlo3` to adjust the fatality rate for each region. |
| `contact_final` | Relative rate of effective contact at end of 2020 | $\sim \text{normal}(1, 0.1) \leq 1$ | To capture continued low incidence of SARS-CoV-2 infection in spite of rising contact rates |
| contact_s0 | Parameter for curve specified by contact_final | ~exponential(0.1) | as shown by mobility data and social contact surveys, we assume that the effective contact rate over time is multiplied by a factor $asc(t/366, 1, contact_{final}, -contact_{s0}, contact_{s1})$, where $t$ is time in days since 1 January 2020. |
| contact_s1 | Parameter for curve specified by contact_final | ~exponential(0.1) |  |
| concentration1 | Increased contact among young people in July | ~normal(2, 0.3) $\geq 2$ | Because initial increases in SARS-CoV-2 prevalence from July in England were especially apparent in young people, we allow increases in mobility to be more emphasized in young people starting from July. We model a relative contact-rate multiplier for individuals of age $a$ as $beta(a/100 \vert a = 0.2(k - 2) + 1, b = 0.8(k - 2) + 1)$, where $k$ is the concentration parameter and $beta$ is the beta distribution probability density function. This gives flat contact rates across age groups when $k = 2$, and relatively higher contact rates in individuals around age 20 when $k > 2$. |
| concentration2 | Increased contact among young people in August | ~normal(2, 0.2) $\geq 2$ | |
| concentration3 | Increased contact among young people from September | ~normal(2, 0.1) $\geq 2$ | |
| disp_deaths,<br>disp_hosp_inc,<br>disp_hosp_prev,<br>disp_icu_prev | Negative binomial dispersion for deaths, hospital incidence (admissions), hospital prevalence (beds occupied), and ICU prevalence | ~exponential(10) | We estimate the size parameter for negative binomial likelihood functions of deaths, hospital incidence, hospital prevalence and ICU prevalence, where size = $1/(disp^2)$ (69) |

Parameters for VOC 202012/01 strain
| Parameter | Description | Prior distribution | Notes |
| --- | --- | --- | --- |
| v2_when | Introduction date of VOC 202012/01 in days after 1 January 2020 | ~uniform(144, 365) | On this date, ten random individuals contract VOC 202012/01 |
| v2_hosp_rlo | Relative log-odds of hospitalisation for VOC 202012/01, compared to preexisting variants | ~normal(0, 0.1) | Vague prior |
| v2_icu_rlo | Relative log-odds of ICU admission for VOC 202012/01, compared to preexisting variants | ~normal(0, 0.1) | Vague prior |
| v2_cfr_rlo | Relative log-odds of death for VOC 202012/01, compared to preexisting variants | ~normal(0, 0.1) | Vague prior |
| v2_relu | Relative transmission rate of VOC 202012/01, compared to preexisting variants | ~lognormal(0, 0.4) | Vague prior |
| `v2_immesc` | Cross protection against VOC 202012/01 due to previous infection by a preexisting variant (0 = no cross protection, 1 = total cross protection) | $\sim\text{beta}(3, 1)$ | Vague prior |
| `v2_ch_u` | Relative susceptibility to VOC 202012/01, compared to preexisting variants, for 0-19yo individuals | $\sim\text{lognormal}(0, 0.4)$ | Vague prior; $v2\_ch\_u = 1$ corresponds to children having reduced susceptibility relative to adults as in ref. (19). Susceptibility of 0-19 year olds to VOC 202012/01 is multiplied by $v2\_ch\_u$. |
| `v2_infdur` | Relative length of infectious period of VOC 202012/01, compared to preexisting variants | $\sim\text{lognormal}(0, 0.4)$ | Vague prior |
| `v2_serial` | Relative length of generation interval of VOC 202012/01, compared to preexisting variants | $\sim\text{normal}(0, 0.4)$ | Vague prior. The latent and infectious periods are multiplied by $v2\_serial$, while the infectiousness is multiplied by $1 / v2\_serial$, in order to maintain overall infectiousness when integrated over the infectious period. |

**Table S3.**
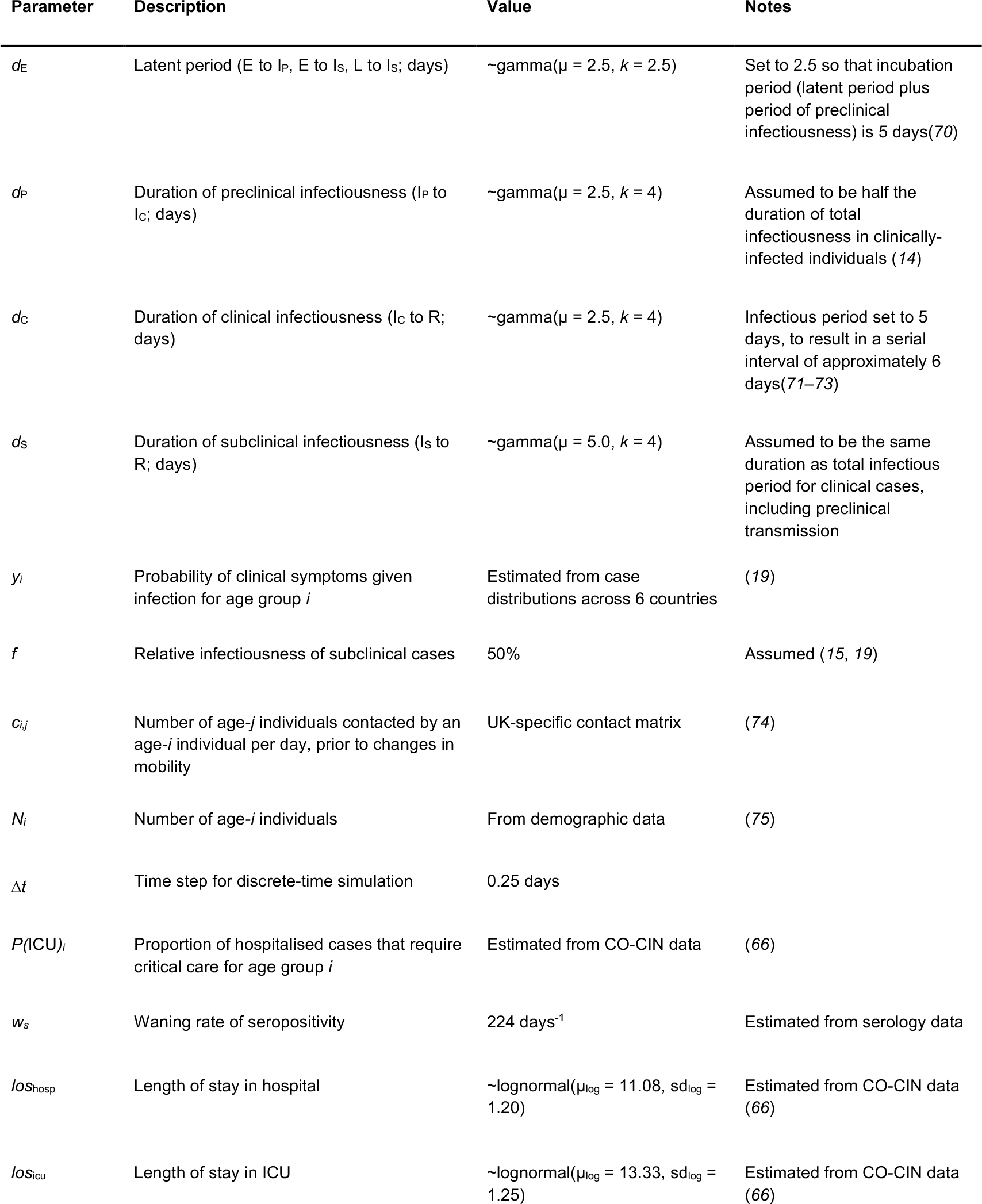
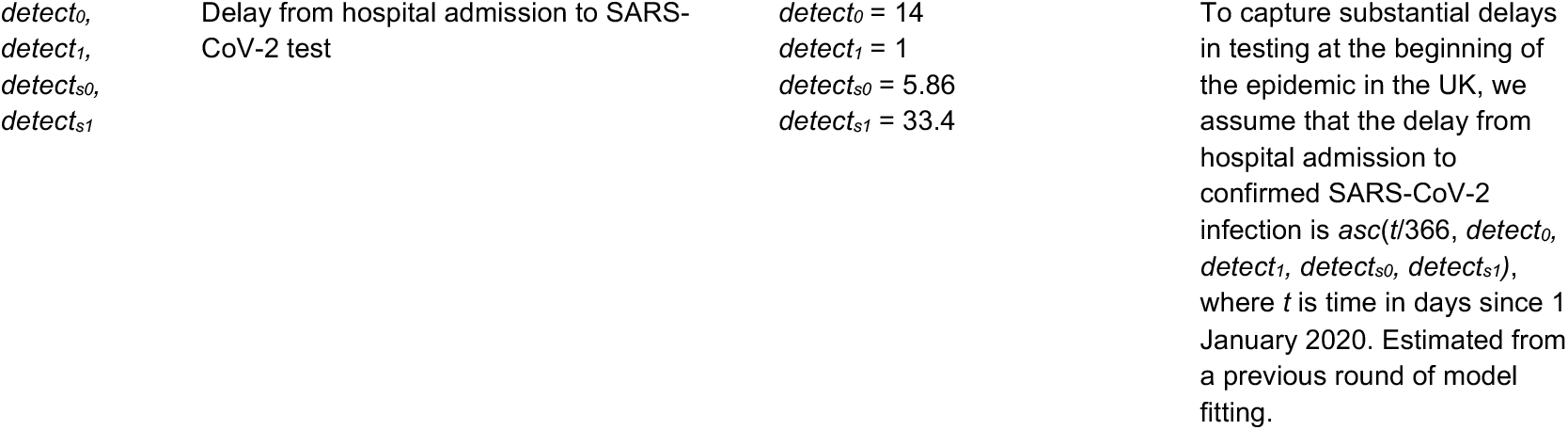
Model parameters not subject to fitting.

**Table S4.** Model comparison for dynamic transmission models.

| Hypothesis | DIC | Predictive deviance | ΔDIC | ΔPD | Rank |
| --- | --- | --- | --- | --- | --- |
| Increased transmissibility | 16246 | 6872 | 4 | 0 | 1 |
| Increased duration of infectiousness | 16242 | 8188 | 0 | 1316 | 2 |
| Immune escape | 19988 | 9314 | 3747 | 2442 | 4 |
| Increased susceptibility in children | 16385 | 8056 | 144 | 1184 | 3 |
| Shorter generation time | 17205 | 58373 | 963 | 51501 | 5 |
| Combined | 16295 | 18141 | 53 | 11269 | — |

**Table S5.** Summary of projections for England, 15 Dec 2020 – 30 June 2021, including a seasonal decline in transmission. Compare to Table 2, main text.

|  | Moderate (October 2020) | High (November 2020) with schools open | High with schools closed | Very high (March 2020) |
| --- | --- | --- | --- | --- |
| **Peak ICU (rel. to 1st wave)** | 271% (254 - 290%) | 160% (150 - 171%) | 130% (122 - 136%) | 119% (113 - 125%) |
| **Peak ICU requirement** | 9,880 (9,240 - 10,500) | 5,830 (5,450 - 6,220) | 4,720 (4,450 - 4,960) | 4,340 (4,110 - 4,550) |
| **Peak deaths** | 3,910 (3,690 - 4,140) | 2,020 (1,900 - 2,130) | 1,500 (1,440 - 1,560) | 1,320 (1,260 - 1,380) |
| **Total admissions** | 627,000 (596,000 - 650,000) | 442,000 (421,000 - 460,000) | 395,000 (376,000 - 411,000) | 334,000 (318,000 - 352,000) |
| **Total deaths** | 213,000 (202,000 - 224,000) | 142,000 (134,000 - 148,000) | 128,000 (122,000 - 134,000) | 104,000 (99,400 - 110,000) |

200,000 vaccinations per week
|  | Moderate (October 2020) | High (November 2020) with schools open | High with schools closed | Very high (March 2020) |
| --- | --- | --- | --- | --- |
| **Peak ICU (rel. to 1st wave)** | 268% (252 - 284%) | 158% (148 - 169%) | 129% (122 - 136%) | 118% (112 - 124%) |
| **Peak ICU requirement** | 9,770 (9,170 - 10,300) | 5,760 (5,390 - 6,140) | 4,710 (4,440 - 4,950) | 4,310 (4,070 - 4,520) |
| **Peak deaths** | 3,630 (3,470 - 3,850) | 1,910 (1,800 - 2,020) | 1,480 (1,430 - 1,550) | 1,320 (1,280 - 1,380) |
| **Total admissions** | 605,000 (579,000 - 636,000) | 427,000 (407,000 - 444,000) | 371,000 (353,000 - 386,000) | 299,000 (285,000 - 314,000) |
| **Total deaths** | 198,000 (190,000 - 210,000) | 133,000 (126,000 - 139,000) | 116,000 (110,000 - 120,000) | 89,200 (84,500 - 93,100) |

2 million vaccinations per week
|  | Moderate (October 2020) | High (November 2020) with schools open | High with schools closed | Very high (March 2020) |
| --- | --- | --- | --- | --- |
| **Peak ICU (rel. to 1st wave)** | 234% (220 - 250%) | 148% (139 - 158%) | 128% (121 - 134%) | 118% (111 - 124%) |
| **Peak ICU requirement** | 8,520 (7,990 - 9,100) | 5,380 (5,050 - 5,730) | 4,650 (4,390 - 4,880) | 4,290 (4,060 - 4,500) |
| **Peak deaths** | 2,450 (2,310 - 2,590) | 1,510 (1,450 - 1,570) | 1,390 (1,340 - 1,450) | 1,290 (1,250 - 1,340) |
| **Total admissions** | 479,000 (455,000 - 497,000) | 348,000 (333,000 - 362,000) | 268,000 (256,000 - 278,000) | 184,000 (177,000 - 191,000) |
| **Total deaths** | 138,000 (132,000 - 145,000) | 97,700 (93,400 - 102,000) | 78,300 (75,000 - 81,300) | 56,700 (54,700 - 58,700) |

**Table S6.**
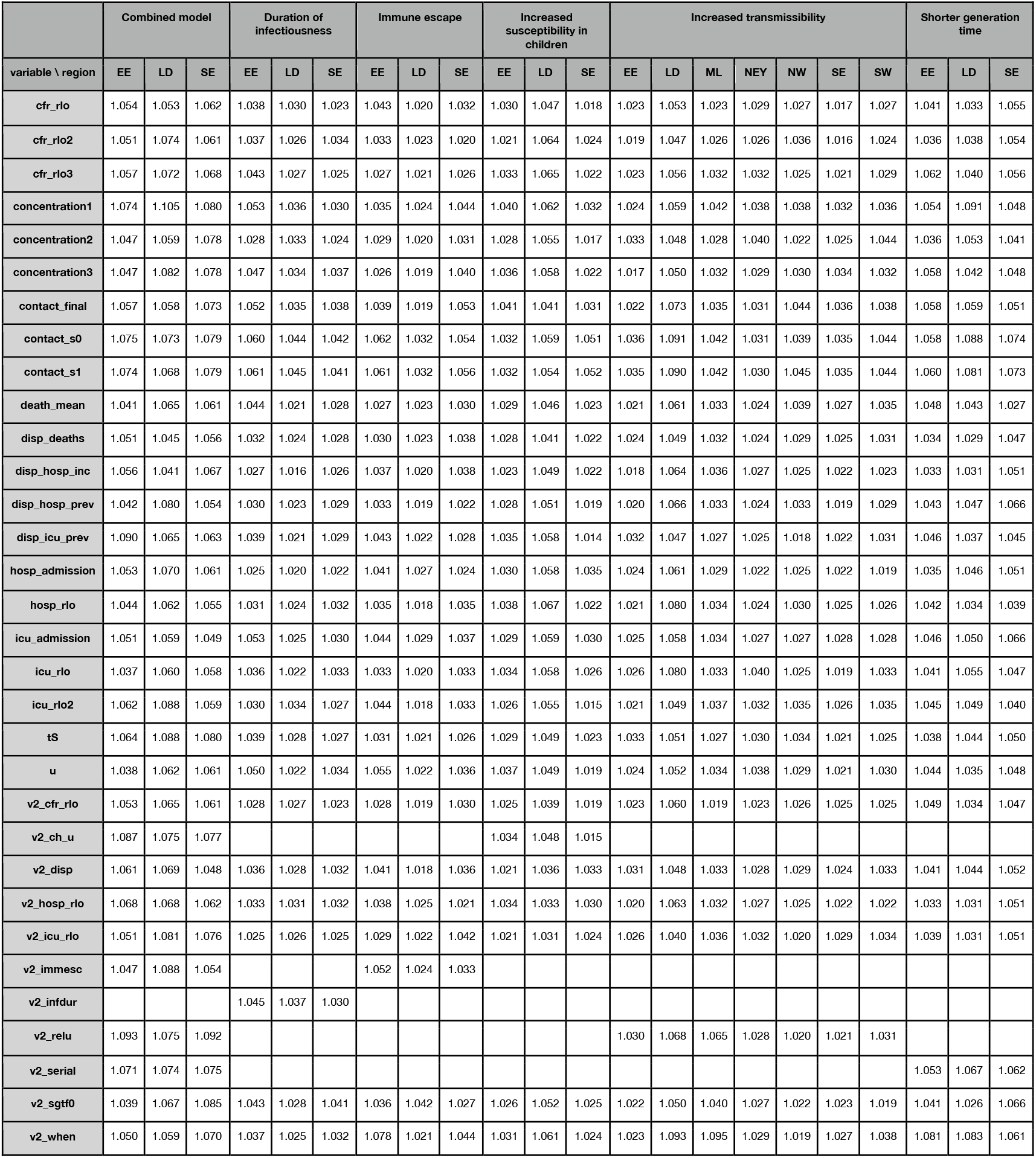
Gelman-Rubin convergence diagnostics 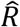 for all transmission models. Regions are East of England (EE), London (LD), Midlands (ML), North East and Yorkshire (NEY), North West (NW), South East (SE), and South West (SW).

